# A Dynamic Genetic Atlas of Gestational Phenotypes

**DOI:** 10.1101/2024.10.15.24315491

**Authors:** Siyang Liu, Hao Zheng, Yuqin Gu, Zijing Yang, Jianxin Zhen, Yuandan Wei, Yanhong Liu, Yanchao Chen, Zijun Wan, Xinxin Guo, Liang Hu, Xiaohang Chen, Xiaotian Li, Xiu Qiu, Shujia Huang, Guo-Bo Chen, Zhibin Hu, Shinichi Namba, Masahiro Kanai, Koichi Matsuda, Yukinori Okada, Quanfu Zhang, Fengxiang Wei

## Abstract

The ∼40-week gestational period is central to human reproduction, yet the genetic architecture of diverse gestational phenotypes and their links to maternal late-life health remain unclear. In 111 phenotypes from up to 121,579 Chinese pregnancies (median n = 78,535 per phenotype), we identified 4,688 independent genome-wide significant loci, including 1,703 novel associations. Gestation-specific effects were observed for 7.8% of variants across 30 phenotypes, and 18.7% of signals for 24 longitudinal hematological traits exhibited gene-by-gestational-timing interactions across five antenatal and postpartum periods. Dynamic genetic effects were enriched in growth- and hormone-regulatory pathways, reflecting maternal–fetal interactions. Genetic correlation and Mendelian randomization analyses with 80 diseases and medication traits in Biobank Japan females revealed shared genetic architecture and potential causal links between gestational phenotypes and maternal mid- and late-life health. These results establish a dynamic genetic atlas of human gestational phenotypes with implications for precision maternal-health strategies. Results are visualized at https://monn.pheweb.com/.

## Introduction

Women’s reproductive, maternal, newborn, child, and adolescent health underpins human well-being and drives long-term population and societal progress^1^. Amid declining fertility, rising early-life disorders, and global population ageing, improving maternal and child health has become a global major public health priority. The approximate 40-week gestational period represents a critical developmental window, during which the fetus undergoes rapid growth and differentiation, accompanied by profound maternal physiological adaptations^2–4^. Accordingly, health systems worldwide implement structured prenatal and postpartum assessments- including anthropometric measures, blood and urine tests and prenatal screening, which collectively define gestational phenotypes. These clinical indicators are central to diagnosing pregnancy-related disorders and predicting birth outcomes (**Fig. 1; Supplementary Table 1**).

**Figure 1.**
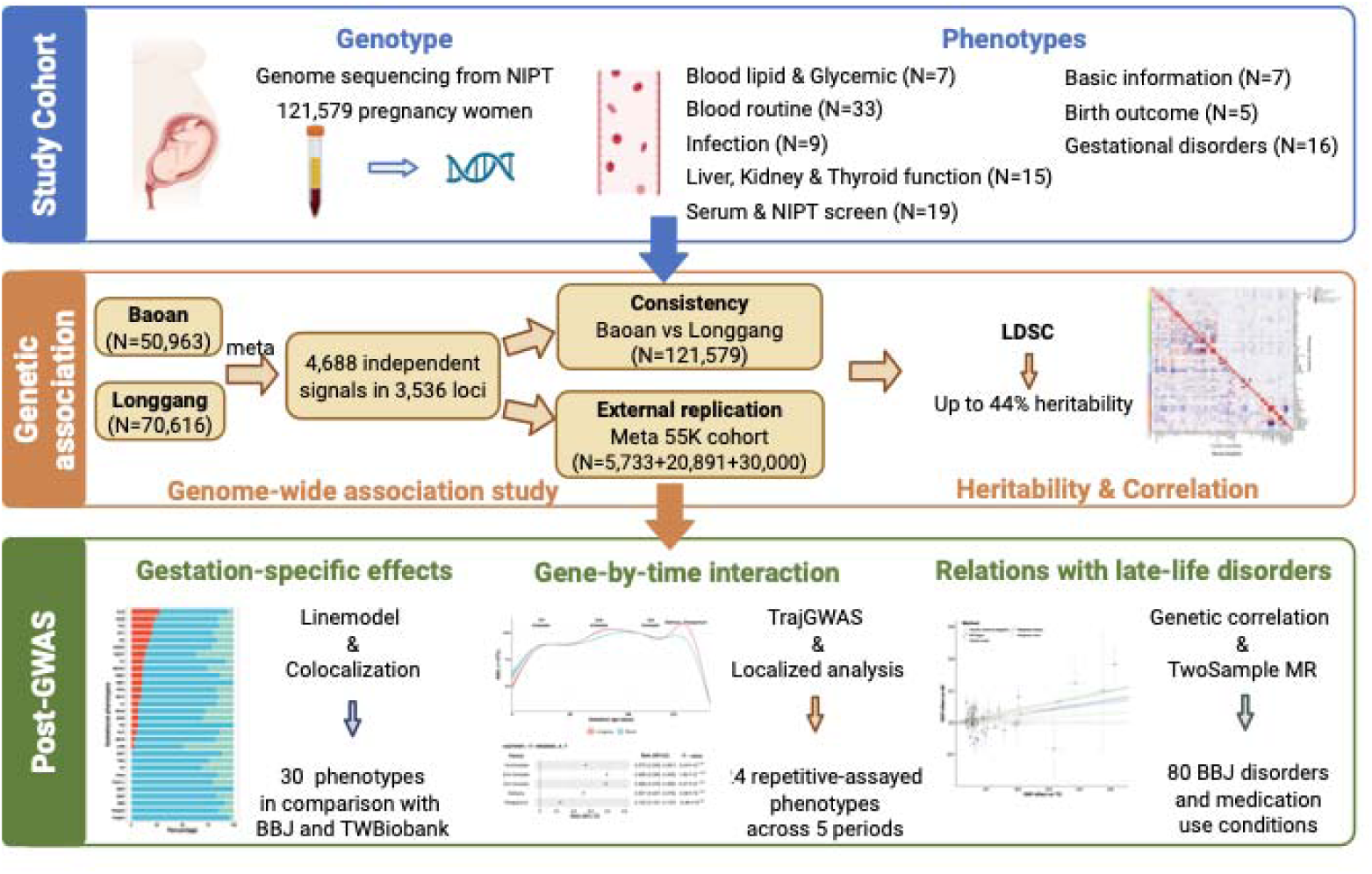
Schematic overview of the study. We assembled a comprehensive dataset of 111 gestational phenotypes from 120,579 Chinese pregnancies. The study characterized the genetic architecture of these phenotypes, identified enriched biological pathways, and examined genetic variants influencing their longitudinal trajectories. We further assessed potential causal relationships between gestational phenotypes and late-life disorders.

Despite their clinical relevance, the genetic underpinnings of gestational phenotypes and their links to long-term maternal health remain poorly characterized. Of the 111 gestational phenotypes examined in this study, only 31 have been evaluated in GWAS with >20,000 participants, 21 of which were reported in our prior studies^5–8^ (**Supplementary Table 1**). Two additional studies^9,10^ - not indexed in the GWAS Catalog were constrained by limited sample size (**Supplementary Notes** and **Supplementary Fig. 1**). Consequently, little is known about the genetic basis of gestational phenotypes or how genetic effects may vary across implantation and fetal development. Moreover, although observational studies have linked gestational conditions to later-life maternal health^11,12^, systematic genetic correlation and causal analyses of gestational phenotypes- typically measured in early adulthood- on mid-and late-life health outcomes remain scare, limiting opportunities for early disease risk stratification in women aged 20–40 years.

To address these gaps, we analyzed 111 prenatal and postnatal clinical gestational phenotypes alongside non-invasive prenatal test (NIPT) sequencing data from up to 121,579 unrelated pregnant women. Using large-scale GWAS and advanced analytical protocol^13,14^, we estimated SNP heritability and genetic correlations, constructed an atlas of gestational trait loci, and performed external replication. We further identified gestation-specific genetic effects using Bayesian classification, and characterized gene-by-time (*G* × *T*) interactions across repeated measurements of 24 traits throughout pregnancy and postpartum. Finally, we integrated this atlas with 80 common mid- and late-life disorders and medication-use conditions from BioBank Japan (mean age 63.0 years)^15^ to assess shared genetic correlations and potential causal relationships, thereby exploring the long-term health implications of gestational phenotypes (**Fig. 1**).

## Results

### Study design

We collected NIPT sequencing data with 111 prenatal and postnatal clinical phenotypes from up to 121,579 unrelated pregnancies receiving routine obstetric care at two hospitals in Shenzhen between 2017 to 2022. Of these, 83 gestational phenotypes comprised molecular and screening-based measures and were classified into nine categories: blood lipid (n = 2), glycemic markers (n = 5), blood routine indices (n = 33), infection-related markers (n = 9), kidney function (n = 3), liver function (n = 8), thyroid function (n = 4), Serum screening (n = 16), and NIPT screening (n = 3) (**Fig.1; Supplementary Table 2**). An additional 28 phenotypes were derived from electronic medical records, including maternal baseline characteristics (n = 7), gestational disorders (n = 16) and birth outcomes (n = 5) (**Fig. 1; Supplementary Table 3**).

Sample sizes ranged from 18,454 (Cytomegalovirus IgM antibody quantification) to 110,343 (trisomy 18 risk score), with a median of 78,535 participants per phenotype. The mean maternal age was 29.7 ± 4.28 years (**Supplementary Fig. 2**; **Supplementary Table 4**). Phenotype distributions, including the prevalence of common gestational comorbidities, were highly consistent across the two hospitals **(Extended Data Fig. 1; Supplementary Tables 4–5).**

Using our established analytical workflow^13,14^, we inferred and imputed genotypes from NIPT sequencing data, estimated family relatedness, characterized population structure and conducted GWAS analyses. Identified associations were evaluated in three independent cohorts: NIPT-PLUS (n= 5,733), Baoan-20K (n=20,891) and Longgang-30K (n=30,000). Detailed methods are provided in the **Methods** and **Supplementary Notes**.

### 4,688 genome-wide association signals

We performed GWAS for the 111 gestational phenotypes across the two hospitals using linear or logistic regression (PLINK), and mixed-model association testing (REGENIE), followed by fixed-effect meta-analysis of 11.6 million variants (**Methods**). Both methods generated genomic inflation factors (λ) ranged from 1.00 to 1.42, and LD score regression (LDSC) intercepts from 1.00 to 1.10, indicating minimal statistical inflation (**Supplementary Table 6A-B**). Observed-scale SNP heritability (and liability-scale heritability for binary phenotypes) estimated by LDSC^16^ ranged from 0% for in vitro fertilization (IVF) to 43.97% for maternal serum vitamin B12 (**Extended Data Fig. 2; Supplementary Fig. 3**), closely approximating reported twin-based estimate of 56% VB12 heritability in White British women^17^.

Phenotypic and genetic correlation matrices were largely symmetric along the diagonal, indicating substantial alignment between phenotypic covariance and shared genetic architecture **(Extended Data Fig. 3; Supplementary Table 7**). Below, we report REGENIE-based results using genome-wide significance (*P* < 5×10^-8^) and a study-wide Bonferroni threshold for 111 traits (*P* < 4.5×10^-10^). Consistent with theorectical expectations^14^, female-only GWAS yielded larger effect-size estimates than mixed-sex analyses owing to model specification differences (**Supplementary Notes; Supplementary Figs. 4–5**).

Using GCTA conditional and joint analysis^18^, we identified 4,688 indepedent association signals across 3,536 loci (signals ±1 Mb merged into a single locus). Of these, 4,009 signals at 2,978 loci were associated with at least one molecular phenotype (*P* < 5×10^-8^), including 2,776 signals at 1,874 loci surpassing the study-wide threshold (*P*<;4.5×10^-10^) (**Fig. 2, Supplementary Tables 8-9**). For maternal basic traits, gestational disorders, and birth outcomes, we detected 679 independent signals at 558 loci (*P* < 5×10^-8^), including 383 signals at 282 loci meeting study-wide significance (**Supplementary Tables 10-11**). Blood routine traits yielded the largest number of associations, with platelet large-cell ratio (P_LCR) exhibiting the highest signal density (**Fig. 2**). Across all loci, we identified 5,045 functional variants (including missense and nonsense variants) at genome-wide significance, of which 4,438 exceeded the study-wide threshold (**Supplementary Tables 12-13**). After consolidating LD (pairwise SNP r^2^ < 0.2) and cross-phenotype similarity, we derived 3,613 independent signals across all 111 phenotypes.

**Figure 2.**
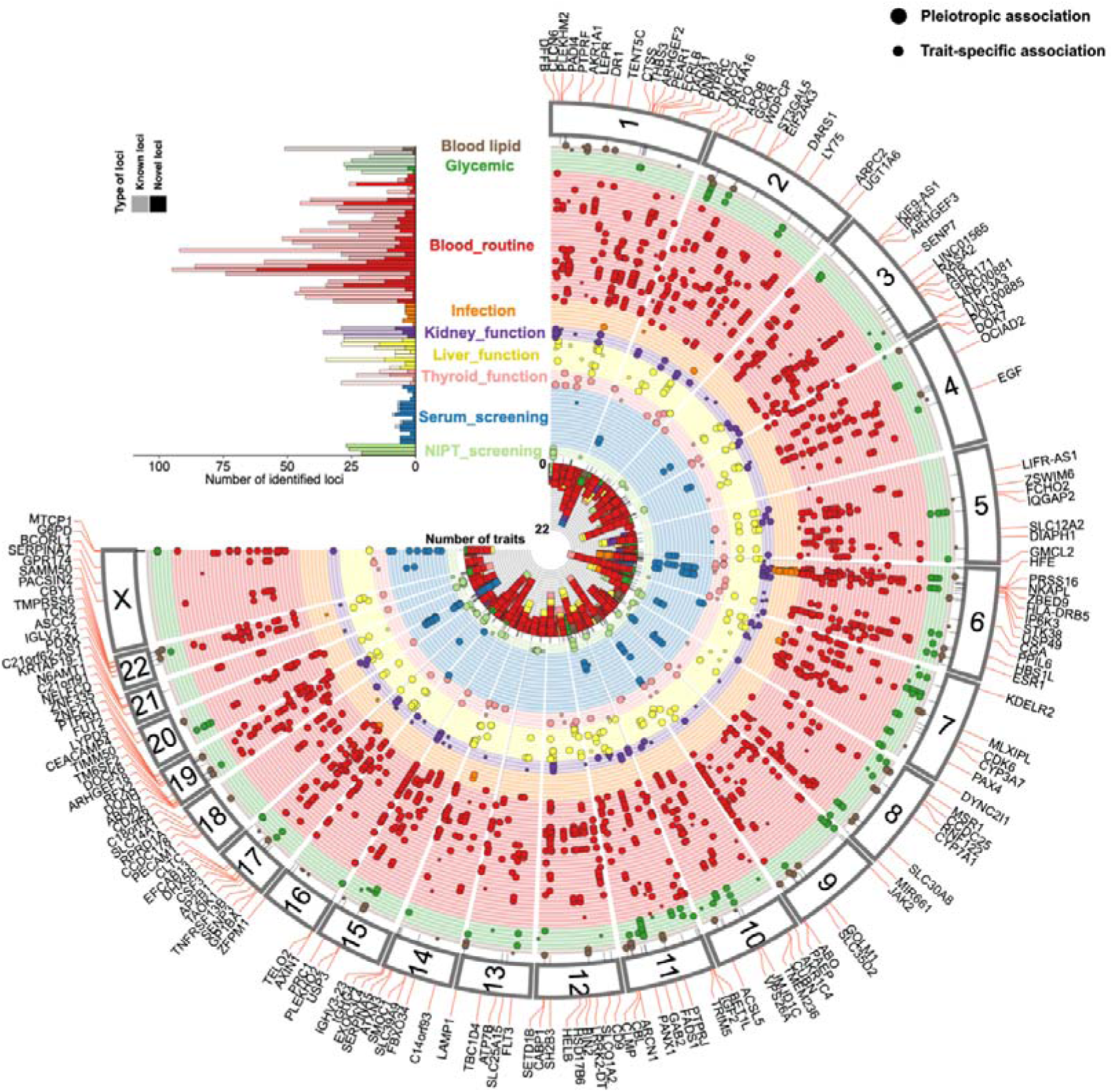
Genetic associations across nine molecular test categories of gestational phenotypes. The top-left inset shows the distribution of loci identified per gestational phenotype, separated into novel and previously reported associations. The main panel presents a Fuji plot summarizing GWAS results across nine molecular test categories, displaying independent variants identified by meta-analysis (*P* < 4.5 × 10^−10^). For each pleiotropic locus, the number of associated phenotypes is indicated by filled coloured boxes, with functional and newly discovered loci labeled. Functional annotations are detailed in Supplementary Table 13. Pleiotropic and trait-specific loci are represented by circles of different sizes. The ordering of the 83 phenotypes in the histogram and Fuji plot follows Supplementary Table 2.

Effect estimates were highly concordant across cohorts. Between the two hospitals, effect sizes were strongly correlated (*Pearson’s* R = 0.75-0.99 for molecular phenotypes and 0.90-0.98 for EMR-derived phenotypes; **Supplementary Fig. 6**). Replication in the three independent cohorts (NIPT-PLUS, Baoan-20K, and Longgang-30K; meta-analysis N =56,624, per-phenotype sample size ranges from 2,336 to 47,991) further confirmed robustness of the effect estimates (**Supplementary Table 14**). Among 4,678 lead variants testable in 106 phenotypes, effect-size correlations ranged from 0.87 to 0.97, with 99.59% showing concordant effect directions, 88.05% further reaching *P* < 0.05 and 62.8% surpassing Bonferroni correction (**Supplementary Fig. 7; Supplementary Table 15**). Similar concordance was observed for study-wide significant variants.

Of all 4,688 genome-wide significant signals, 404 exhibited pleiotropic associations with multiple gestational phenotypes (**Supplementary Table 16**). PPIN analysis highlighted several hub genes, including *ESR1*, *ALB*, *BCL2,* and *PTPRC* (**Supplementary Fig. 8; Supplementary Table 7**). For example, *ESR1,* which regulates embryo implantation through uterine natural killer cell motility and early placental vascular remodeling^19^, showed pleiotropic effects on leukocyte-related and glycemic traits (**Supplementary Fig. 9**). Comprehensive pleiotropy profiles for all significant variants are available through the MONN PheWeb.

### 1,703 novel genetic association signals

By comparing our results with previously reported associations in the GWAS Catalog^20^ (e114_r2025-07-10; **Supplementary Table 18**) and the two pregnancy-focused GWAS not indexed in the GWAS Catalog^9,10^, we identified a substantial number of novel association signals. Among 4,009 genome-wide significant signals across the nine molecular phenotype categories, 1,536 (38.3%) showed no prior associations for the same or related trait (LD R^2^ < 0.2) (**Supplementary Table 8**).

Similarly, 593 of 1,874 study-wide significant signals (31.6%) were novel (**Supplementary Table 9**). For EMR-derived phenotypes, 167 of 679 genome-wide significant signals (24.5%), and 65 of 383 study-wide significant signals (16.9%) were previously unreported (**Supplementary Table 10-11**). After consolidating cross-phenotype similarity and LD (pairwise SNP r^2^ < 0.2), these 1,703 novel associations (across 1,245 loci) resolved into 1,335 independent novel signals. Among these, 293 study-wide significant novel signals containing at least one functional variant are illustrated in **Fig. 2**.

Novel loci were particularly enriched in maternal serum screening and NIPT-based fetal aneuploidy screening metrics, reflecting their historical underrepresentation in large-scale GWAS (**Fig. 2**). Maternal serum and NIPT screening are central to fetal trisomy risk assessments^21,22^ (**Supplementary Notes**), and our results indicate that a measurable portions of variance in these clinical risk scores is attributable to common genetic variation. For maternal serum screening, nine independent loci were associated with first- and second-trimester risk scores for trisomies 21 and 18 (**Extended Data Fig. 4)** . Phenome-wide association analysis in the MONN PheWeb linked these variants to proteins incorporated into clinical risk-score calculation.

For NIPT-based screening, we identified 28, 34 and 34 loci associated with risk scores for trisomy 21 (T21), trisomy 18 (T18) and trisomy 13 (T13), respectively (*P* < 5×10^-8;^ **Supplementary Fig. 10)** . Many loci mapped to the corresponding trisomic chromosomes (20/28 for T21; 13/34 for T18; 16/34 for T13), likely reflecting interactions between SNP-derived genotype likelihood and local copy-number amplification. Notably, several loci—*PADI4* (chr1), *CCDC71L* (chr7), and *PANX1* (chr11)—were shared across T21, T18, and T13. The associated variants act as cis-eQTLs for these genes. *PADI4* has established roles in progenitor cell proliferation and translation in developing hair collicles^23^, *CCDC71L* has roles in immune regulation^24^,and *PANX1* encodes a membrane channel highly expressed in human oocytes and early embryos, with loss-of-function variants causing familial “oocyte death” infertility syndrome^25,26^. Variants associated with reduced expression of *PANX1, PADI4 and CCDC71L* were linked to higher NIPT trisomy risk scores in our analysis. These findings suggest previously unrecognized biological contributions to aneuploidy-related risk scores and indicate that integrating maternal genomic variation may improve the accuracy of clinical risk prediction models.

### Gestation-specific genetic effects

Gestational phenotypes may exhibit genetic effects distinct from those observed outside pregnancy. To assess gestation-specific effects, we compared our GWAS results for 30 phenotypes with data from non-pregnant female participants in the Biobank of Japan^15^ (BBJ; n = 28,315 to 76,026), and for 21 phenotypes from the Taiwan Biobank^27^ (**Supplementary Table 19**). Among 1,136 genome-wide significant signals replicated under Bonferroni correction in independent cohorts, effect size estimates were available for 905 signals in BBJ females. Effect-size concordance between gestational and non-pregnant females varied substantially (*Pearson’s R* = 0.46–0.91), and LDSC-based genetic correlations for the same phenotypes ranged from 0.34 to 1.00 (**Supplementary Table 20**).

To distinguish gestation-specific from general genetic effects, we applied a Bayesian clustering algorithm^28^ combined with colocalization analysis^29^ **(Supplementary Fig. 11; Supplementary Table 21; and Methods).** Loci were classified as shared (general), gestation-specific, or unclassified. Of the 905 signals, 742 were classifiable, including 71 (7.8%) identified as gestation-specific and 708 (78.2%) as shared (**Supplementary Fig. 12; Supplementary Table 22**). Albumin (ALB) exhibited the highest proportion of gestation-specific variants, followed by hemoglobin (HGB). In contrast, maternal height, early-pregnancy anthropometric traits (weight, BMI, DBP, SBP), and several molecular test phenotypes (TP, TBIL, AST, HbA1c) showed no evidence of gestation-specific genetic effects, consistent with their relative physiological stability during pregnancy (**Fig. 3A, Supplementary Table 23**).

**Figure 3.**
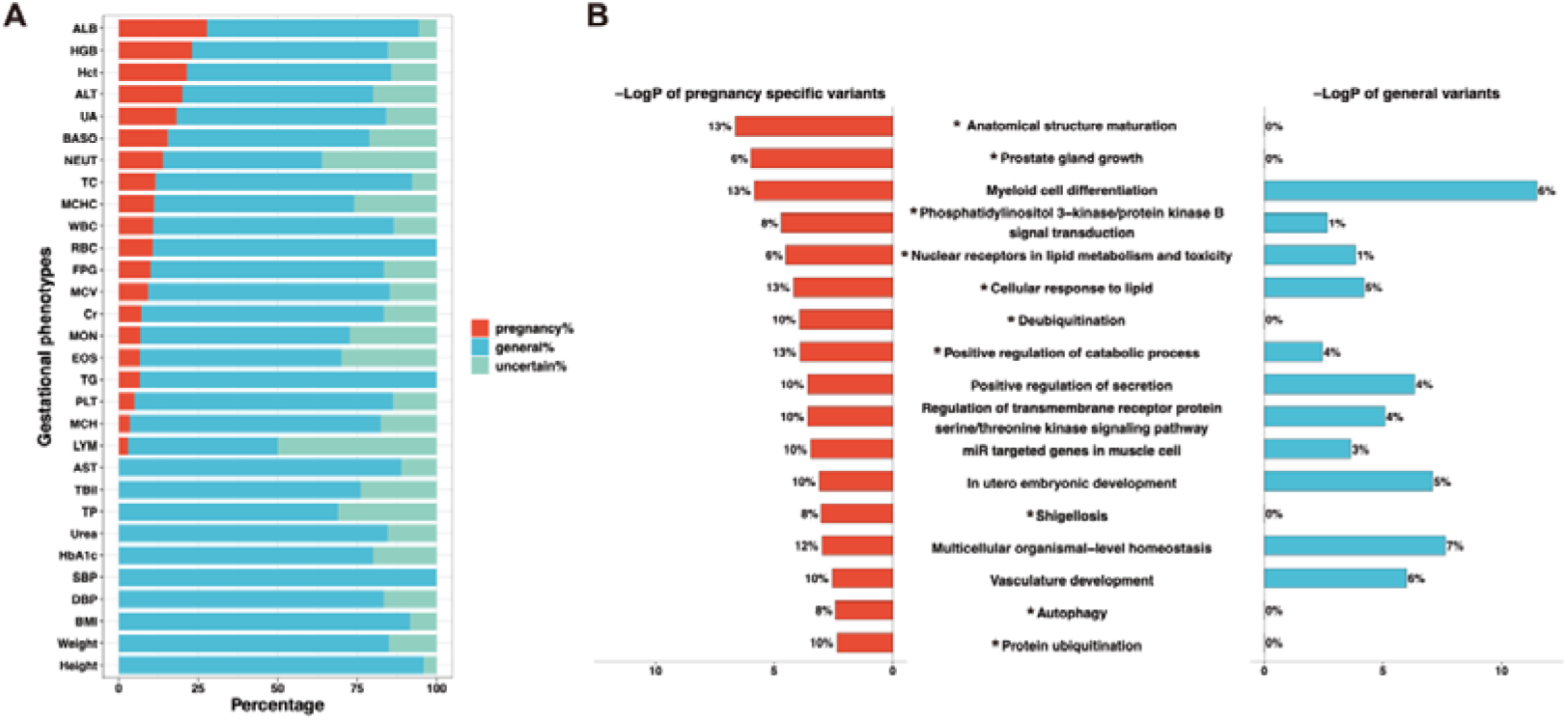
Characteristics of loci exhibiting gestation-specific genetic effects. **A**, Classification of SNP effects passing Bonferroni-corrected replication using a Bayesian framework combined with colocalization analysis. **B**, Butterfly bar plot showing the top 20 pathways enriched for genes harboring gestation-specific variants, identified using Metascape. Left: enrichment P values and the proportion of pregnancy-specific genes within each pathway. Right: corresponding results for general variants. Pathways with significantly different enrichment between gestation-specific and general loci (chi-square/fisher exact test, *P* < 0.05) are marked with an asterisk (*). Trait abbreviations: ALB, albumin; HGB, hemoglobin; Hct, hematocrit; ALT, alanine transaminase; UA, uric acid; BASO, basophils; NEUT, neutrophils; TC, total cholesterol; MCHC, mean corpuscular haemoglobin concentration; WBC, white blood cell count; RBC, red blood cell count; FPG, fasting plasma glucose; MCV, mean corpuscular volume; Cr, creatinine; MON, monocytes; EOS, eosinophils; TG, triglycerides; PLT, platelets; MCH, mean corpuscular haemoglobin; LYM, lymphocytes; AST, aspartate transaminase; TBil, total bilirubin; TP, total protein; Urea, urea; HbA1c, glycated haemoglobin; SBP, systolic blood pressure; DBP, diastolic blood pressure; BMI, body mass index;

Albumin illustrates a representative gestation-specific pattern. Of 18 high-quality ALB-associated signals identified in this study, five were classified as gestation-specific, showing robust associations during pregnancy but not in BBJ or the Taiwan Biobank (**Supplementary Table 22-23**). A representative locus, rs4764725-C (β 95% CI: 0.08 - 1.10, P= 4.43 x 10^-40^), is shown in **Extended Data Fig. 5**. This variant is a cis-eQTL for *ASCL1*, a transcription factor involved in neuronal commitment and differentiation, including the development of olfactory and autonomic neurons^24^. Its pregnancy-restricted association suggests gestation-dependent regulatory activity potentially linked to fetal development.

Pathway enrichment analyses revealed distinct functional profiles for gestation-specific and shared loci (**Supplementary Fig. 13)**. Gestation-specific loci were preferentially enriched in pathways related to fetal development and pregnancy maintenance, including anatomical structure maturation, prostate gland growth, autophagy, protein ubiquitination and deubiquitination, and shigellosis (two-tailed χ² or Fisher’s exact test; **Fig. 3b**; **Supplementary Tables 24–25**). PPI network analysis highlighted key hub regulators among gestation-specific genes, including the IGF1 (Insulin Like Growth Factor 1)^24^, ESR1 (Estrogen Receptor 1)^24^, and PPARG which plays a role in adipocyte differentiation and glucose regulation^24^, implicating growth-and hormonal- regulatory pathways as central mechanisms underlying gestation adaptation (**Supplementary Fig. 14; Supplementary Table 26**).

Because genetic effects present in non-pregnant populations but absent during pregnancy may represent an alternative form of gestation specificity, we further examined 283 genome-wide significant signals for 21 shared phenotypes that were replicated in BBJ female participants and the Taiwan Biobank. Only nine variants showed female-specific effects outside pregnancy, being significant in non-pregnant females but not during gestation (**Supplementary Table 27**). These variants were enriched exclusively in lipid homeostasis pathways (**Supplementary Table 28**). Methodological considerations regarding the identification of gestation-specific effects using female-only versus mixed-sex GWAS are detailed in the **Supplementary Notes** and **Supplementary Fig. 15**.

### G×T interactions across gestation and the postpartum period

Among the 111 gestational phenotypes examined, 24 complete blood count (CBC) traits were repeatedly measured at a median of 5 to 6 time points across five periods in each of the two hospitals (**Supplementary Fig. 16-17; Supplementary Table 29**), enabling systematic investigation of time-varying genetic effects during gestation.

All 24 traits exhibited pronounced and highly concordant physiological trajectories across hospitals (**Extended Data Fig. 6**). White blood cell (WBC)–related traits, including WBC, neutrophil count and percentage (NEUT, NEU_P), and monocyte count (MON), increased throughout pregnancy and declined postpartum, whereas lymphocyte, eosinophil and basophil measures decreased during gestation and rebounded after delivery. Red blood cell indices showed progressive reductions in RBC count, haemoglobin (HGB) and haematocrit (HCT), accompanied by gestational increases in mean corpuscular indices and sustained elevations in red cell distribution width, followed by postpartum recovery. Platelet count and plateletcrit declined until delivery, whereas platelet size indices increased sharply in late gestation.

To identify genetic determinants of these temporal dynamics, we conducted gene-by-time (G×T) interaction analyses using an extended trajectory GWAS framework^30^ (**Methods**). In a two-stage design, genome-wide G×T signals were first screened using score tests (**Supplementary Figs. 18-25)**, and subsequently evaluated using Wald tests and inverse-variance-weighted meta-analysis across hospitals. We identified 663 variants associated with trait means (*P* < 5×10^-8^), of which 124 (18.7%) demonstrated significant **G×T** interactions (*P* < 5×10^-8^) (**Supplementary Table 30**); the remaining 539 variants influenced mean trait levels without detectable termporal modulation (**Supplementary Table 31**). The proportion of interacting variants varied across phenotypes, ranging from 5.4% to 47.8% (**Supplementary Table 32**).

To resolve temporal effect patterns of these 124 interacting variants, we performed localized association analyses across five periods (first trimester, second trimester, third trimester, delivery, and postpartum; **Methods**). 82 interacting variants (66.1%) exhibited non-overlapping 95% confidence intervals for genetic effects between at least two periods (**Supplementary Table 33**). The top 25 variants interacting variants are shown in **Fig. 4A**, of which 22 were strong eQTLs.

**Figure 4.**
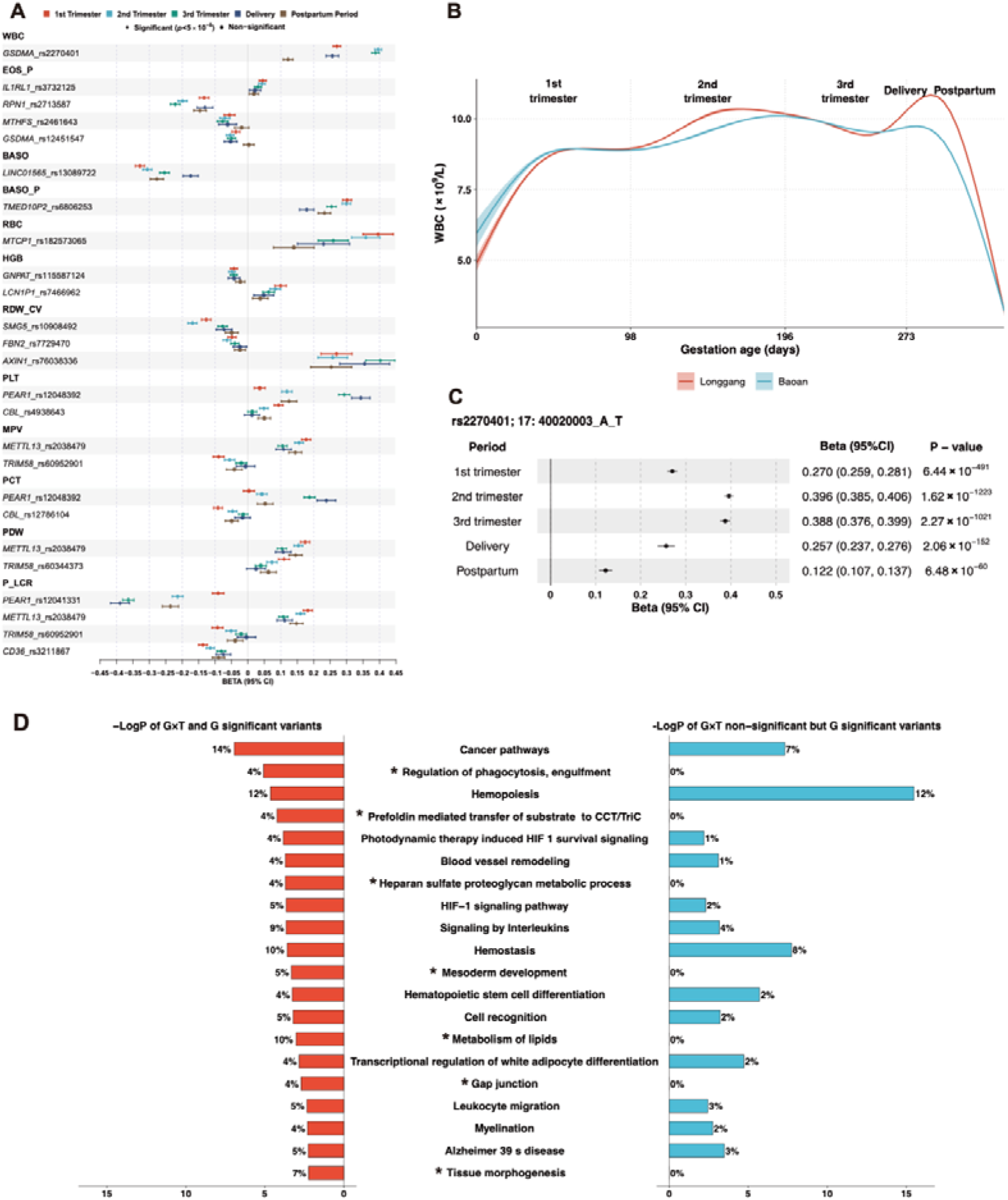
Gene-by-time interactions for 24 blood cell phenotypes during gestation. **A**, Forest plots of genetic effects across five gestational and postpartum periods for the 25 variants with the strongest gene-by-time (G×T) interactions, each showing non-overlapping 95% confidence intervals in at least two periods. Colours denote gestational periods, with effect sizes and confidence intervals shown; variants less than 5 × 10^−8^ are annotated. eQTL status is indicated next to each variant (see Supplementary Table 31). **B**, Trajectories of white blood cell (WBC) counts across trimesters, delivery, and postpartum in two hospitals, modelled using generalized additive models; shaded ribbons indicate 95% confidence intervals. **C**, Forest plot of the genetic effect of rs2270401-A at 17q21.1 (an eQTL for GSDMA) on WBC counts, based on meta-analysis across all five periods. Points represent effect estimates and error bars show 95% confidence intervals. **D**, Enrichment analyses comparing variants with significant versus non-significant G×T interactions. Pathways with significantly different enrichment between gestation-specific and general loci (chi-square/fisher exact test, *P* < 0.05) are marked with an asterisk (*). Trait abbreviations: WBC, white blood cell count; EOS_P, Eosinophil percentage; BASO: basophil absolute value; BASO_P: basophil percentage; RBC, red blood cell count; HGB: hemoglobin level; RDW_CV: red blood cell volume distribution width variation; PLT: platelet count; MPV: mean platelet volume; PCT: platelet crit; PDW: platelet distribution width; P_LCR: platelet-large cell ratio.

WBC count illustrates a representative example. WBC levels increased steadily from early pregnancy, peaked in late gestation and at delivery, and declined postpartum (**Fig. 4B**). Of 27 WBC-associated variants, seven (25.9%) displayed significant **G×T** interactions. The strongest signal, rs2270401 at 17q21.1, a cis-eQTL for *GSDMA*, showed marked temporal variation in effect size, with a 3.33-fold higher effect in the second trimester compared with postpartum (**Fig. 4C**). The remaining interacting loci exhibited similar U-shaped trajectories (**Supplementary Fig. 26**), highlighting dynamic genetic regulation of leukocyte expansion during pregnancy.

Finally, pathway enrichment analyses revealed that G×T-interacting variants were preferentially enriched in pathways related to phagocytosis, heparan sulfate metabolism, mesoderm development, lipid metabolism, gap junctions and tissue morphogenesis, compared with non-interacting variants (**Fig. 4D; Supplementary Table 34; Supplementary Fig. 27**). PPIN analyses highlighted *ESR1*, the estrogen receptor 1^24^; *EGF*, a key regulator of cell proliferation^24^; and *GATA2*, a master transcription factor in hematopoietic development^24^, as major hubs (**Extended Data Fig. 7; Supplementary Table 35**). Together, these findings indicate that a distinct subset of variants exerts dynamic, time-dependent genetic effects on maternal hematological traits during gestation, implicating hormonal and developmental regulatory pathways in gestational adaptation.

### Relations with common disorders and medication use

Gestation occurs predominantly in early adulthood (95% CI, 21–37 years in this study), offering a potential window for identifying genetic predisposition to later-life disease. To examine the long-term health relevance of gestational phenotypes, we evaluated their relationships with 58 common disorders (predominantly late-onset diseases) and 22 medication-use traits (prevalence >1%) from Biobank Japan (BBJ; mean age at recruitment, 63.0 years)^15^ (**Supplementary Table 36**) . We first estimated genome-wide genetic correlations between 111 gestational phenotypes and BBJ traits using LD score regression, followed by two-sample Mendelian randomization (MR). Analyses were conducted primarily in female BBJ participants and complemented by full-cohort analyses to balance sex-specific modeling with statistical power (Methods; Supplementary Notes).

In the female-only genetic correlation analysis, 82 significant correlations were identified (FDR *Q* < 0.05), primarily linking 36 gestational phenotypes to type 2 diabetes and related medication use (**Extended Data Fig. 8a; Supplementary Table 37**). However, owing to limited heritability for some BBJ traits in females, genetic correlations could be computed for only 5.9% of all phenotype pairs. In contrast, full-cohort analyses enabled estimation for 85.4% of pairs, yielding 266 significant correlations (FDR Q < 0.05; **Extended Data Fig. 8b; Supplementary Table 38**).

Beyond diabetes-related traits, these included correlations between gestational adiposity and cardiocerebrovascular diseases, eosinophil indices and allergic traits, alanine transaminase and cholelithiasis, and thyroid peroxidase antibody levels and thyroid hormone use.

To assess the direction of the relationship, we further performed MR analyses between 108 gestational phenotypes with genome-wide significant instruments and the 80 BBJ outcomes, excluding weak instruments, outliers, pleiotropic associations, heterogeneous effects, and incorrect causal directions (Methods; Supplementary Tables 39–41). In female participants, 20 associations passed FDR correction (*Q* < 0.05 across 8640 tests) (**Fig. 5A-B; Supplementary Fig. 28; Supplementary Table 42; scatter plots in Supplementary Figs. 29-30**). These included genetically higher gestational triglycerides increasing myocardial infarction risk (OR = 1.13, 95% CI = 1.07–1.21), elevated OGTT measures and gestational diabetes increasing risks of diabetes and related medication use, higher gestational diastolic blood pressure increasing angina risk, hepatitis B serological markers increasing chronic hepatitis B risk, and higher total bilirubin increasing cholelithiasis risk (**Fig. 5C; Supplementary Figs. 29–30**). All associations were supported by at least one sensitivity analysis, and three had prior causal evidence (**Supplementary Tables 42–43**).

**Figure 5.**
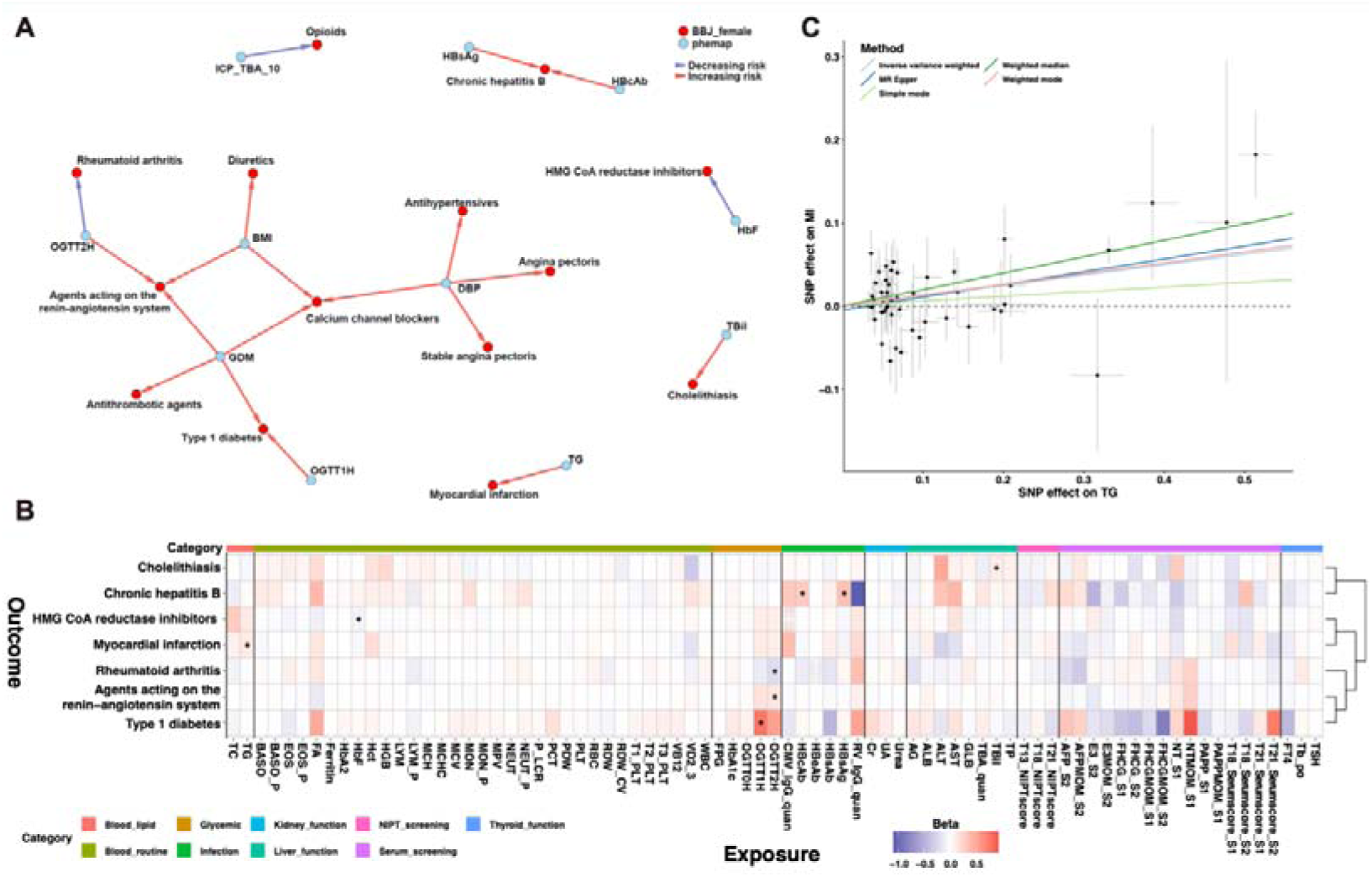
Mendelian randomization linking gestational phenotypes to common disorders and medication use in BBJ females. a, Mendelian randomization (MR) estimates accounting for pleiotropy, heterogeneity, and reverse causation, illustrating putative causal relationships between gestational phenotypes (blue nodes) and late-life disorders in BioBank Japan (BBJ; red nodes). Arrows indicate direction of effect, with red denoting risk-increasing and blue risk-decreasing associations. b, Heatmap of MR results for nine categories of gestational phenotypes (right) as exposures and BBJ disorders (bottom) as outcomes. Associations surpassing the FDR threshold are marked with an asterisk (*, q < 0.05). Beta values shown in the legend and heatmap correspond to inverse-variance weighted (IVW) estimates. c, MR scatterplot illustrating the effect of maternal triglyceride (TG) levels on the risk of myocardial infarction in BBJ females.TG, triglyceride; HbF, hemoglobin F; OGTT1H, oral glucose tolerance test 1 hour; OGTT2H, oral glucose tolerance test 2 hour; HBcAb, hepatitis B core antibodies; HBsAg, hepatitis B surface antigen; TBil, total bilirubin; BMI, body mass index; GDM, gestational diabetes mellitus; ICP_TBA_10, intrahepatic cholestasis of pregnancy diagnosed by total bile acids >10 μmol/L; MI: myocardial infarction.

In the full BBJ cohort, which afforded greater power, 78 associations reached FDR significance, of which 40 satisfied all pleiotropy, heterogeneity and directionality criteria and were also significant in female-only analyses (FDR Q<0.05 for 78 tests; **Extended Data Fig. 9; Supplementary Table 44)**. Two dominant clusters emerged: (i) gestational adiposity and cardiometabolic traits influencing cardiovascular and cerebrovascular disease risk; and (ii) gestational glycaemic traits increasing diabetes risk while inversely associating with asthma, rheumatoid arthritis and cerebral aneurysm. Additional associations linked immune, hepatic and hematological gestational traits to cancer, gallstone disease and atrial fibrillation. Corresponding medication-use associations were also observed. Twenty-two of these 40 associations were supported by prior causal inference, and all but one were corroborated by sensitivity analyses (**Supplementary Tables 44–45**).

To determine whether the observed MR associations reflect pregnancy-specific biological effects, we performed multivariable Mendelian randomization incorporating GWAS data from the Taiwan Biobank (mean age at recruitment, 50.0Lyears) for 22 phenotypes shared with our study. After adjustment for genetic effects estimated in the Taiwan Biobank, all associations were attenuated to null, indicating that the observed relationships are unlikely to result from irreversible biological effects incurred during pregnancy (**Supplementary Table 46**).

Complete forward MR results are summarized in **Supplementary Tables 43 and 45**. The distribution of the MR Egger 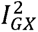 is shown in **Supplementary Fig. S31.** Scatter, forest, leave-one-out and funnel plots of each MR analyses are available at https://monn.pheweb.com/ (Mendelian randomization module).

## Discussion

Human genetic diversity is shaped not only by spatial but also by temporal variation. Although the genetic architecture of mid to late-life traits has been extensively characterized, early-life phenotypes- particularly during gestation- remains poorly understood. Here, we present a large-scale genetic investigation of 111 gestational phenotypes. We identified 4,688 independent genome-wide significant signals, of which 1,703 had not been previously reported for the same or related traits. After accounting for linkage disequilibrium and cross-phenotype similarity, 3,613 independent signals remained, including 1,335 novel loci. Replication analyses showed highly consistent effects, with >99.6% of associations displaying concordant directions, 88.1% reaching nominal significance, and 62.8% exceeding Bonferroni-corrected thresholds. SNP heritability estimates ranged from 0% to 43.97%.

The resulting genetic atlas is dynamic, reflecting interactions between maternal genetics and the evolving intrauterine environment. Integrating data from the Biobank of Japan and the Taiwan Biobank with Bayesian classification and colocalization analyses of 905 high-confidence signals across 30 phenotypes, we identified 71 gestation-specific loci (7.8%) absent in non-pregnant populations, with IGF1, ESR1 and PPARG emerging as key hubs. The proportion of gestation-specific effects varied widely, from near zero for early-pregnancy anthropometric traits to 27.8% for liver function markers. Trajectory GWAS of repeated measures for 24 complete blood count traits across five gestational periods revealed significant gene-by-time interactions for 18.7% of associated variants (124 of 663). Protein–protein interaction analyses highlighted ESR1, EGF and GATA2 as central nodes. Pathway enrichment analysis of genes exhibiting dynamic genetic effects highlighted roles in developmental and hormonal regulatory processes.

Gestational traits may offer an early window for identifying women at risk of common diseases later in life^11^. However, the mechanisms underlying these observational associations remain unclear. In this study, genetic correlation and Mendelian randomization analyses of 111 gestational phenotypes with 80 common disorders and medication-use traits revealed shared genetic architecture and potential causal relationships. Notable findings included elevated gestational glycemic traits increasing diabetes risk; higher gestational adiposity, lipid levels, and blood pressure elevating cardiovascular risk; increased white blood cell counts reducing susceptibility to certain cancers, whereas infection-related markers increased risk for specific malignancies; higher total bilirubin associating with cholelithiasis; and greater standing height with atrial fibrillation. Multivariable MR incorporating 22 shared phenotypes from the Taiwan Biobank attenuated all associations, suggesting these relationships do not arise from irreversible effects of pregnancy. Expanding gestational and population cohorts with broader exposures and outcomes will be critical to disentangle direct versus indirect effects and to guide precision early-warning strategies for women’s long-term health using the framework established here.

Despite substantial advances, this study has limitations. First, although it represents the largest GWAS of gestational phenotypes to date, statistical power remains limited for certain traits. We excluded binary phenotypes with fewer than 2,000 cases, precluding analysis of clinically important but underpowered phenotypes such as gestational hypertension, pre-eclampsia, macrosomia, hyperemesis gravidarum, and intruterine hypoxia. As non-invasive prenatal testing and pregnancy screening expand globally, future large-scale, cross-population studies will be well positioned to investigate these phenotypes. Our open-access workflow and summary statistics provide a foundation for such efforts.

Second, we hypothesize that the observed gestational-specific and interaction effects may be fetal-driven, but the underlying molecular mechanisms remain unclear. Recent work on *GDF15 in* hyperemesis gravidarum- where fetal-derived GDF15 in maternal plasma mediates disease- illustrates how targeted assays can distinguish maternal and fetal contributions^31^. Applying similar strategies to gestation-specific loci identified here could clarify these interactions.

Third, several caveats apply to our assessment of links between gestational phenotypes and long-term maternal health. Restricting analyses to female participants mitigates effect-estimate bias but reduces power; we therefore complemented female-only analyses with whole-cohort analyses. As more sex-specific GWAS become available in East Asian populations, this limitation is expected to lessen. Despite using a stringent MR framework—accounting for instrument strength, pleiotropy, heterogeneity, and causal directionality—and observing support from sensitivity analyses and prior evidence, residual false positives cannot be excluded. We plan to iteratively update these results in the MONN PheWeb as exposure and outcome GWAS expand. Additionally, while we focused on maternal long-term health, intergenerational effects on offspring were not assessed. Quantifying these effects requires family-based genomic data combined with intergenerational MR^32–34^ or structural equation modelling^35^, resources that are currently limited in East Asia.

Nevertheless, as birth cohort genomic studies expand in China^32,36^, the instruments identified here will enable powerful investigation of gestational influences on offspring health across the life course.

This study provides foundational insights into the dynamic genetic atlas of human gestational phenotypes. To support future research, we provide interactive visualization of GWAS results via the MONN PheWeb server (https://monn.pheweb.com/). As MONN grows, future efforts will prioritize increasing power for rare gestational outcomes, elucidating the molecular basis of gestation-specific and temporal genetic effects, and leveraging the findings here to advance risk prediction, prevention and precision medicine for maternal and neonatal disorders. Coordinated global efforts will be essential for fully realize the full translational potential of these discoveries.

## Methods

### Study population

Between 2017 and 2022, we recruited 129,403 pregnant women receiving routine maternity care at the Longgang District Maternity and Child Healthcare Hospital and Shenzhen Baoan Women’s and Children’s Hospital in Shenzhen, China. Participants underwent prenatal and postnatal serum biomarkers testing and non-invasive prenatal testing (NIPT) covered by the local medical insurance program. In addition, we collected their basic demographic information, gestational complications, and birth outcomes from electronic medical records. Detailed participant counts per test are provided in **Supplementary Tables 2-3**. Maternal age distribution, phenotypic concordance between the two hospitals are summarized in **Extended Data Fig. 1; Supplementary Fig. 1; Supplementary Table 4-5**.

Maternal genotypes were inferred from the low-depth whole-genome sequencing data generated from NIPT, utilizing our previously established methods^13,14^. Related individuals up to third-degree relatedness were excluded based on the procedures described in the Methods section “ Family relatedness and principal component analyses,” resulting in 121,579 unrelated pregnancies included in all downstream genetic analyses.

By integrating obstetrical, genotypic, and phenotypic data, we investigated the genetic basis of gestational phenotypes. This study was approved by the Medical Ethics Committee of the School of Public Health (Shenzhen), Sun Yat-sen University, Longgang District Maternity and Child Healthcare Hospital of Shenzhen City, and Shenzhen Baoan Women’s and Children’s Hospital. Data collection was authorized by the Human Genetic Resources Administration of China (HGRAC).

### Phenotype definition

To ensure adequate statistical power and accuracy for common variant discovery and effect size estimates, we excluded binary phenotypes with fewer than 2,000 case or controls, less than 2,000, as recommended for GWAS of complex traits^37^. Particularly, we removed phenotypes with insufficient case counts, including but not limited to hepatitis B E-antigen positivity, herpes simplex virus type I/II IgM or type II IgG antibody seropositivity, toxoplasma IgG antibody positivity, stillbirth, twin pregnancy, overt hyperthyroidism, preeclampsia, vitamin B12, folate deficiency anemia, macrosomia, and intruterine hypoxia.

Quantitative traits were retained because all had sample sizes exceeding 18,000 participants. After filtering, 111 gestational phenotypes remained. These phenotypes were grouped into nine molecular test categories—blood lipids (N=2), glycemic markers (N=5), blood routine indices (N=33), infection-related markers (N=9), kidney function (N=3), liver function (N=8), thyroid function (N=4), serum screening (N=16), and NIPT screening (N=3)—and three medical-record-based categories, including maternal baseline characteristics (N=7), gestational disorders (N=16), and birth outcomes (N=5) (**Supplementary Table 2-3**). 24 complete blood count traits were measured at an average of five to six time points; all other phenotypes were typically measured once. For repeated phenotypes, we used the earliest measurement in the primary GWAS and performed trajectory GWAS to investigate potential gene-by-time interactions.

### Genotype imputation from NIPT data and quality control

Non-invasive prenatal testing (NIPT) sequences cell-free DNA (cfDNA) from maternal plasma for fetal aneuploidy screening^38^, generating low- to mediate-coverage genomic sequencing data. In our previous study, we demonstrated that NIPT data can be effectively utilized for GWAS, generating highly accurate variant discoveries and GWAS effect estimates^13,14^. We used our established analytical framework in this study.

In brief, genotype dosages were imputed from NIPT data using genotype-likelihood-based imputation and phasing implemented in GLIMPSE (version 1.1.1)^39^, with a reference panel of high-depth whole-genome sequencing genotypes from 10,000 Chinese individuals. Genotype imputation accuracy was assessed in 100 NIPT participants with matched high-depth sequencing data, yielding an average R^2^ > 0.84 for variants with minor allele frequency (MAF) > 0.01, as defined by the reference panel, and INFO score > 0.4, as computed by GLIMPSE. Accordingly, variants with MAF < 0.01 or INFO score < 0.4 were excluded from downstream analyses in this study.

### Family relatedness and principal component analyses

We used PLINK^40^ (version 2.0) to extract SNPs with MAF ≥ 5%. Kinship coefficients were estimated using the KING-robust algorithm, applying a cutoff of 0.044 to remove duplicate samples and individuals related up to the third degree; one individual from each related pair was retained in subsequent analyses. In total, 7,824 participants were excluded. Principal component analysis (PCA) was then conducted using PLINK on the remaining 50,963 and 70,616 participants for Baoan and Longgang hospitals.

### Variant annotation

Variant annotation was performed using the Ensembl Variant Effect Predictor (VEP) (version 101)^41^, with indexed GRCh38 cache files (version 109). HGVS nomenclature was generated from the primary *Homo sapiens* reference FASTA. Since a variant may map to multiple transcripts, we applied the --pick option to assign a single representative annotation per variant based on VEP’s default prioritization. For intergenic variants, the --nearest option was used to identify the nearest gene according to their protein-coding transcription start site (TSS).

### Genome-wide association and conditional analysis

After quality control of samples and genotype imputation as described above, genome-wide association analysis (GWAS) were performed for all 111 phenotypes using linear mixed models implemented in REGENIE^42^ (version 4.1). Quantitative phenotypes were first pre-processed by applying a rank-based inverse normal transformation (quantile transformation) to standardize distributions across participants. This approach ensures approximate normality of the traits, minimizes the influence of outliers, and allows effect estimates to be interpreted per standard deviation change in the transformed phenotype.We included maternal age, the top ten principal components, and gestational week at measurement (when available) as covariates in Step 1.

GWAS summary statistics from the two participating hospitals were combined by inverse-variance weighted (IVW) fixed-effect meta-analysis using METAL^43^ (version 2011-03-25). Fuji plot for visualization were generated using custom R scripts and Circos^44^. Meta-analysis summary statistics were used in all downstream analyses.

Independent genome-wide significant signals were identified using multi-SNP conditional and joint analysis (COJO^18^) implemented in GCTA^45^ with stepwise model selection (--cojo-slct). A collinearity threshold of 0.2 was applied. Loci were defined as a merged ±500 kb window around each genome-wide significant signal, with the variant showing the smallest *P* value designated as the lead SNP. Associations passing the threshold of P< 5.8 x 10^-l0^and the threshold of P < 4.5 x 10^-l0^ were considered genome-wide and study-wide significant.

### Replication of independent genome-wide significant signals

We applied two methods to assess the robustness of the independent association signals. First, we assessed the consistency of genetic effect estimates between the two discovery hospitals using Pearson’s correlation coefficient (R). Second, we compared effect estimates from the discovery meta-analysis with those from three independent replication cohorts- the NIPT-PLUS cohort (N = 5,733), the Baoan 20K cohort (N = 20,891), and the Longgang 30K cohort (N = 30,000)- as well as their meta-analysis. These participants underwent maternal check-ups at Shenzhen Baoan Women’s and Children’s Hospital (Shenzhen, China) or Longgang District Maternity and Child Healthcare Hospital after the year of 2022. Duplicate samples with the discovery sets were already removed using their unique identifiers. Of the 111 gestational phenotypes analyzed, 106 were available in the replication datasets (**Supplementary Table 14**). A SNP was considered replicated if it exhibited a consistent effect direction, and exceeded the Bonferroni-corrected significance threshold (*P* < 0.05/number of independent SNPs per phenotype). Given the modest sample size of the replication cohorts, we additionally report the number of SNPs with consistent effect directions, and those with concordant direction and nominal significance (*P* < 0.05). In addition, Pearson’s correlation coefficients between discovery and replication effect estimates are provided.

### Genetic heritability and correlation estimates

Because summary statistics from linear mixed models (LMM) cannot be used to estimate SNP-heritability owing to the use of an effective rather than the true sample size^16^, we estimated SNP-based observed-scale heritability as well as liability-scale heritability for binary traits using meta-GWAS summary statistics generated with PLINK2.0, and computed phenotype-wise genetic correlations (*r_g_*) using LD Score Regression^46^ based on REGENIE results. LD scores were calculated with GCTA using high-depth whole-genome sequencing data from the same 10,000-sample Chinese reference panel used for genotype imputation. The genome was divided into 1000-kb segments (--ld-wind 1000), and an LD R^2^ cutoff of 0.01 was applied (--ld-rsq-cutoff 0.01). Phenotypes were standardized, and Spearman correlation coefficients were computed to generate the phenotypic correlation matrix. Phenotypic and genetic correlation matrices from both hospitals were extracted for visualization using heat maps.

### Identification of novel signal and loci

We compiled genome-wide association data from the GWAS Catalog (gwas_catalog_v1.0.2-associations_e114_r2025-07-10.tsv)^20^, and incorporated significant variants from two pregnancy-related studies whose data were not submitted to GWAS catalog^9,10^. A locus was considered novel if the ±500kb region surrounding the lead SNP did not overlap with previously reported associations for the same or similar traits (**Supplementary Table 18**). An association signal was classified as novel if the LD *R^2^* between the identified SNPs and any previously reported SNP for the same or similar traits was less than 0.2. LD *R^2^* was calculated using the LDpair tool within LDlink^47^ (version 5.5.1) based on the GRCh38 1000 Genomes reference for East Asian (EAS) and European (EUR) populations.

### Detection of gestation-specific genetic effects

To distinguish gestation-specific from general genetic effects, we applied a Bayesian clustering algorithm^28^ combined with colocalization analysis. To mitigate potential biases from sex-specific GWAS models and population structure (**Supplementary Note 4**), we integrated GWAS summary statistics for 30 shared phenotypes (**Supplementary Table 21**) from female participants in the BBJ, and for 21 of these phenotypes, from a male-female mixed population in the Taiwan Biobank^27^. Independent associated SNPs were included if present in either BBJ or Taiwan GWAS, with matched effect alleles and minor allele frequencies. Only association signals surpassing Bonferroni-corrected significance in replication cohorts were analyzed in the following to minimize the false positive judgement of gestation-specific genetic effects.

We applied the linemodels package^28^ (https://github.com/mjpirinen/linemodels) to cluster 905 independent signals (well-replicated in the replication cohorts) across the 30 phenotypes (**Supplementary Table 22**) into gestation-specific or general based on effect sizes. Variants with significantly different effects from the general population (posterior probability > 0.8) were classified as gestation-specific; variants with consistent effects (posterior probability > 0.8) were classified as general; all others were labeled uncertain. Clusters were modeled as lines with a fixed slope of 0 for the gestational cluster, correlation constrained to 0.999, and remaining parameters (slope, scale, and correlation) estimated via an EM algorithm (**Supplementary Table 21**).

To account for LD structure, we performed colocalization analyses between gestational variants and variants for the same phenotypes in BBJ and Taiwan Biobank. Prior probabilities for a genetic locus being associated with gestational phenotypes (*p*_1_) and phenotypes in Japan or Taiwan Biobank (*p*_2_), or both (*p*_12_) were set at 1×10^-4^, 1×10^-4^, and 5×10^-6^, respectively^29^. L*inemodels* classification were adjusted based on colocalization results following these criteria (**Supplementary Fig. 7**):

1. **General**: linemodel posterior probability of general ≥ 0.8 and coloc ≥ 0.2, or posterior < 0.8 but coloc ≥ 0.5
2. **Gestation-specific**: posterior probability of gestation ≥ 0.8 and coloc ≤ 0.2
3. **Uncertain**: all remaining combinations

Assuming most loci are general rather than pregnancy-specific, a locus was ultimately classified as gestation-specific only if it was gestation-specific in both BBJ and Taiwan GWAS. If either indicated general, the locus was classified as general; all other loci were considered uncertain. Remaining gestation-specific effects reflect loci replicated in pregnancy cohorts but lacking associations in both BBJ-female and Taiwan GWAS.

We performed an additional analysis of 283 genome-wide significant signals in BBJ female participants that were replicated in the Taiwan Biobank, representing high-confidence associations outside pregnancy using the same protocol described above.

These significant associations present in non-pregnant populations but absent during pregnancy may represent another type of gestation-specific effect.

### Detection of G×T interaction effects

We applied TrajGWAS^30^ to identify genetic variants associated with longitudinal changes in 24 repeated-measure CBC phenotypes across the antenatal and postnatal periods. First, following Ko et al.^30^, a score test was performed for each SNP to jointly assess the main genetic effect and the gene-by-time interaction (G + G x T) for both mean (β) and within-subject variance (τ).

Subsequently, SNPs with *P* < 5×10^-8^ for either β= 0 or τ= 0 proceeded to a Wald test to estimate effect sizes for genotype dosage (β_g_ or τ_g_) and gene-time interaction terms (β_GXT_ or τ_GXT_) using a mixed-effects model:

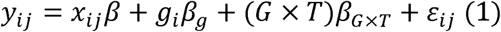

where y_ij_ is the phentoype of the jth measurement for the ith participant; x_ij_ includes fixed-effects covariates (first ten principal components, maternal age, and gestational week) with effect β: g_i_ denotes genotype dosage with effect β_g_ ; and *Gx T* represents the genotype-gestational time interaction with effect β_GXT_. The random-effect term ε_ij_ models within-subject (WS) variance, with variance 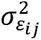 defined as:

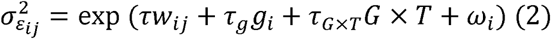

where w_ij_ are covariates, r their effects, r_g_ the genetic effect on WS variance, r_GXT_ the interaction effect, and w_i_ a random intercept.

Results from each of the two hospitals were meta-analyzed using METAL. Independent loci were defined by selecting the lead SNP (smallest *P*) within 1 Mb, grouping all variants within ±500 kb into a single locus.

To assess temporal variation of genetic effects across five periods—first, second, third trimester, delivery day, and within 42 days postpartum—we conducted localized association analyses using PLINK 2.0 A variant was classified as temporally changing if its 95% confidence intervals for genetic effects did not overlap in at least two periods.

### Pathway enrichment analysis

Pathway enrichment was performed using Metascape (v3.5.20240101)^48^. For each pathway, Metascape calculates the observed hit rate in the input gene set relative to the background rate, with the enrichment factor defined as the ratio of these two rates. Statistical significance was assessed using a cumulative hypergeometric test, reflecting the probability that the observed enrichment would occur by chance.

Analyses were conducted using default settings: minimum gene overlap of 3, *P*-value threshold of 0.01, and minimum enrichment factor of 1.5. The search space included Gene Ontology Biological Processes, Reactome gene sets, KEGG pathways, WikiPathways, Canonical Pathways, and PANTHER pathways.

### Protein-protein interaction network analysis

Genes harboring loci identified in this study were subjected to protein-protein interaction (PPI) analysis. Genes with PPI score > 0.4 were included in the network construction, which was visualized using Cytoscape (Version:3.10.1)^49^. Hub genes were ranked using the degree algorithm implemented in CytoHubba, and the top 10 nodes were designated as hub genes. All analyses were performed using default parameters.

### Genetic correlation and Mendelian randomization analysis across 80 BBJ disorders and medication conditions

To assess whether gestational phenotypes are linked to common mid-late-age disease risk, we conducted genetic correlation and two-sample Mendelian randomization (MR) analyses using GWAS data of our cohort and from the Biobank Japan (BBJ), including both female-only and mixed-sex GWAS results.

From the BBJ release of 220 phenotypes (159 disease endpoints, 23 medication categories, and 38 quantitative traits)^15^, we selected 80 traits, including 58 common disease endpoints and 22 medication-use traits. Participants in BBJ had a mean recruitment age of 63.0 years, and 46.3% were female. Assessment methods and diagnostic criteria of these 80 common diseases/medication use are described in the BBJ data release^15^. Diseases irrelevant to maternal populations (for example, prostate cancer), as well as rare (prevalence <1%) or acute conditions, were excluded. Sample sizes for all BBJ outcomes are summarized in **Supplementary Table 34**. BBJ female-only GWAS adjusted for age, age^2^, and the top 20 principal components. Mixed-sex BBJ GWAS further adjusted for sex, ageL×Lsex, and age^2^ ×Lsex. Missing data were not imputed in any exposure or outcome GWAS.

Genetic correlation analyses were performed using the same methodology described above. MR analyses adhered to MR-STROBE reporting guidelines (**Supplementary Information**). A detailed comparison of MR estimates derived from female-only versus mixed-sex GWAS models is provided in **Supplementary Note 5**.

We employed the TwoSampleMR package (version 0.5.7)^50^ and selected independent SNPs identified by GCTA-COJO (*P* < 5×10^-8^) as genetic instruments. The effect sizes of each variant on the exposure were taken from the GWAS summary statistics. The phenotypic variance (PVE) by each instruments was calculated using the following formula(3) and (4)^51^, and instrument strength was assessed using the *F*-statistic; values > 10 were considered indicative of sufficiently strong instruments.

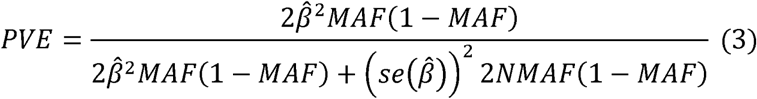

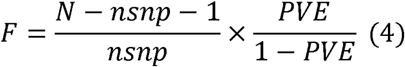

Sensitivity analyses included 1) MR Egger; 2) Simple mode; 3) Weighted median; and 4) Weighted mode estimators, with the inverse variance weight (IVW) method used as the primary reference owing to its superior statistical power when horizontal pleiotropy is absent or balanced.

Horizontal pleiotropy was evaluated using MR-PRESSO47^52^; when significant distortion was detected, identified outliers were removed and analyses were repeated. Steiger’s directionality test was applied to assess potential reverse causation.

Causal effects were considered robust if they met all of the following criteria after removal of MR-PRESSO outliers: (i) significant IVW association after FDR correction (q < 0.05)^53^; (ii) no evidence of heterogeneity (*P* > 0.05); and (iii) no evidence of horizontal pleiotropy; (*P* > 0.05).(iv) Steiger’s test supporting the inferred causal direction (P < 0.05) and no significant associations in the reverse MR .

To account for the potential influence of mid-life health status on causal relationships, we conducted multivariable MR analyses incorporating 22 phenotypes from the Taiwan Biobank as secondary exposures. In these analyses, gestational phenotypes and Taiwan Biobank phenotypes were used as exposures, while the 80 phenotypes from the BBJ were used as outcomes. The TwoSampleMR package was used for these analyses, with a significance threshold of *q* < 0.05.

## Data availability

The full GWAS summary statistics generated in this study have been deposited in the GWAS catalog database (https://www.ebi.ac.uk/gwas/) under accession numbers (GCSTXXXXXX), with approval from the China’s National Health Commission (permission number: 2024SQXB002074). Raw sequencing data have been deposited in the Genome Sequence Archive (GSA) for Human at the National Genomics Data Center, under the BioProject accession number GSA-Human: HRA006833 with approval from China’s National Health Commission (permission number: 2024BAT01079). Data can be accessed via applications, following the GSA guide (https://ngdc.cncb.ac.cn/gsa-human/document). The access authority can be obtained for academic research use only. GWAS summary statistics for phenotypes from the Taiwan Biobank are available in the GWAS Catalog (https://www.ebi.ac.uk/gwas/, accession numbers GCST90278615–GCST90278674). GWAS summary statistics for the BioBank Japan (BBJ) phenotypes were obtained from https://pheweb.jp/ and the female only GWAS summary statistics were provided by Prof. Yukinori Okada.

## Code availability

All code used in this study is publicly available. Code for variant calling, population structure analysis, and genome-wide association studies is available at: https://github.com/liusylab/NIPT-human-genetics.

Code for downstream analyses—including Plink GWAS, Regenie GWAS, SNP heritability and genetic correlation estimation, detection gestation-specific genetic effect, gene-by-time interaction analysis, and two-sample Mendelian randomization—is available at: https://github.com/liusylab/MONN-Genetics.

## Contributions

Design, management, or subject recruitment of the individual studies: S.L., F.W., Q.Z., and data curation: J.Z., Y.W., Z.Y., H.Z., L.H., S.C., and genotyping of the individual studies: Y.W., Y.G., Y.L., Z.Y., and statistical methods, analysis, bioinformatics, or interpretation of the results in the individual studies: S.L., H.Z., Y.G., Z.Y., Y.L., Y.W., X.G, Y.C.,X.L., Z.H.,S.N., M.K., K.M., Y.O., and providing professional guidance and interpretation of data: S.L., F.W., J.Z., S.H., G.C., X.Q., Q.Z., and drafting of manuscript: S.L., H.Z., and critical revision of manuscript: all authors.

## Acknowledgement

The study was supported by National Natural Science Foundation of China (82522078, 32470642, 32470679, 31900487, 82203291, 82173525), Shenzhen Science and Technology Program (20220818100717002, ZDSYS20230626091203007, JCYJ20250604184708012), Guangdong Basic and Applied Basic Research Foundation (2022B1515120080, 2020A1515110859), the Shenzhen Health Elite Talent Training Project and the High-performance Computing Public Platform (Shenzhen Campus) and Tianhe Super Computating platform of SUN YAT-SEN UNIVERSITY. Y.O. was supported by JSPS KAKENHI (25H01057), and AMED (JP24km0405217, JP24ek0109594, JP24ek0410113, JP24kk0305022, JP223fa627001, JP223fa627002, JP223fa627010, JP223fa627011, JP22zf0127008, JP24tm0524002, JP24wm0625504, JP24gm1810011), JST Moonshot R&D (JPMJMS2021, JPMJMS2024), Takeda Science Foundation, Ono Pharmaceutical Foundation for Oncology, Immunology, and Neurology, Bioinformatics Initiative of Osaka University Graduate School of Medicine, Institute for Open and Transdisciplinary Research Initiatives, Center for Infectious Disease Education and Research (CiDER), and Center for Advanced Modality and DDS (CAMaD), Osaka University, RIKEN TRIP initiative (AGIS). We would like to thank Prof. Xia Shen from Fudan University for helpful discussions over the G by T analysis. We would like to acknowledge the valuable assistance provided by the Smart Health Center of Long and Baoan hospitals during the data processing stage- In particular, we thank Song Xu, Qijun Cui, Zhuyan Huang, Shushi Luo, and Guodong Guo for their substantial support. We thank Dr. Saori Sakaue for contribution to the GWAS analysis of BBJ.

## Competing interests

The authors declare no competing interests.

---

**Extended Data Fig. 1.**
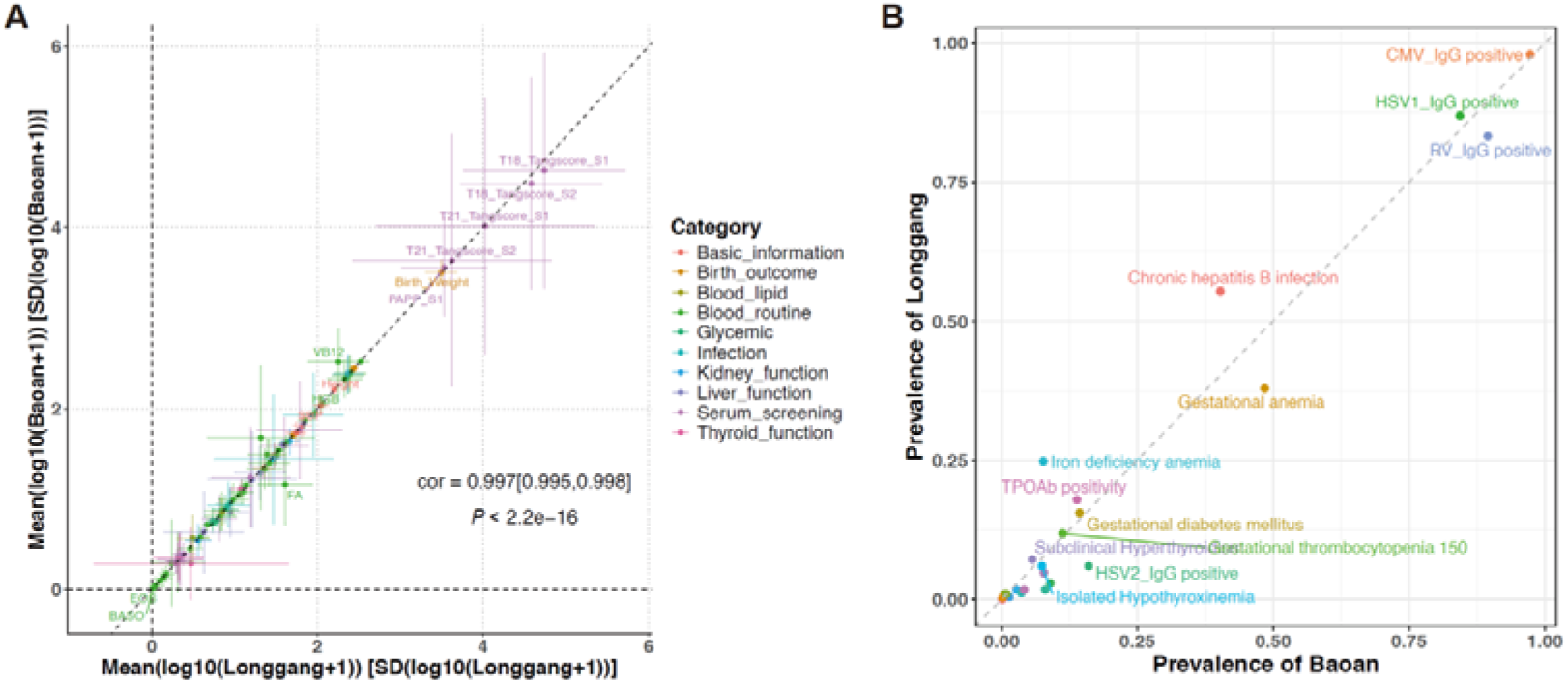
Phenotypic concordance between the two hospital cohorts. A, Correlation of 83 quantitative phenotypes between the Baoan and Longgang hospital cohorts. Each point represents a phenotype, plotted on a log10 scale. The Pearson correlation coefficient (95% CI) is indicated. The dashed grey line (slope = 1, intercept = 0) denotes identity. B, Concordance in the prevalence of gestational disorders or qualitative traits between the two cohorts. Prevalence in Baoan Hospital is plotted against prevalence in Longgang Hospital. The dashed grey line (slope = 1, intercept = 0) represents perfect agreement.

**Extended Data Fig. 2.**
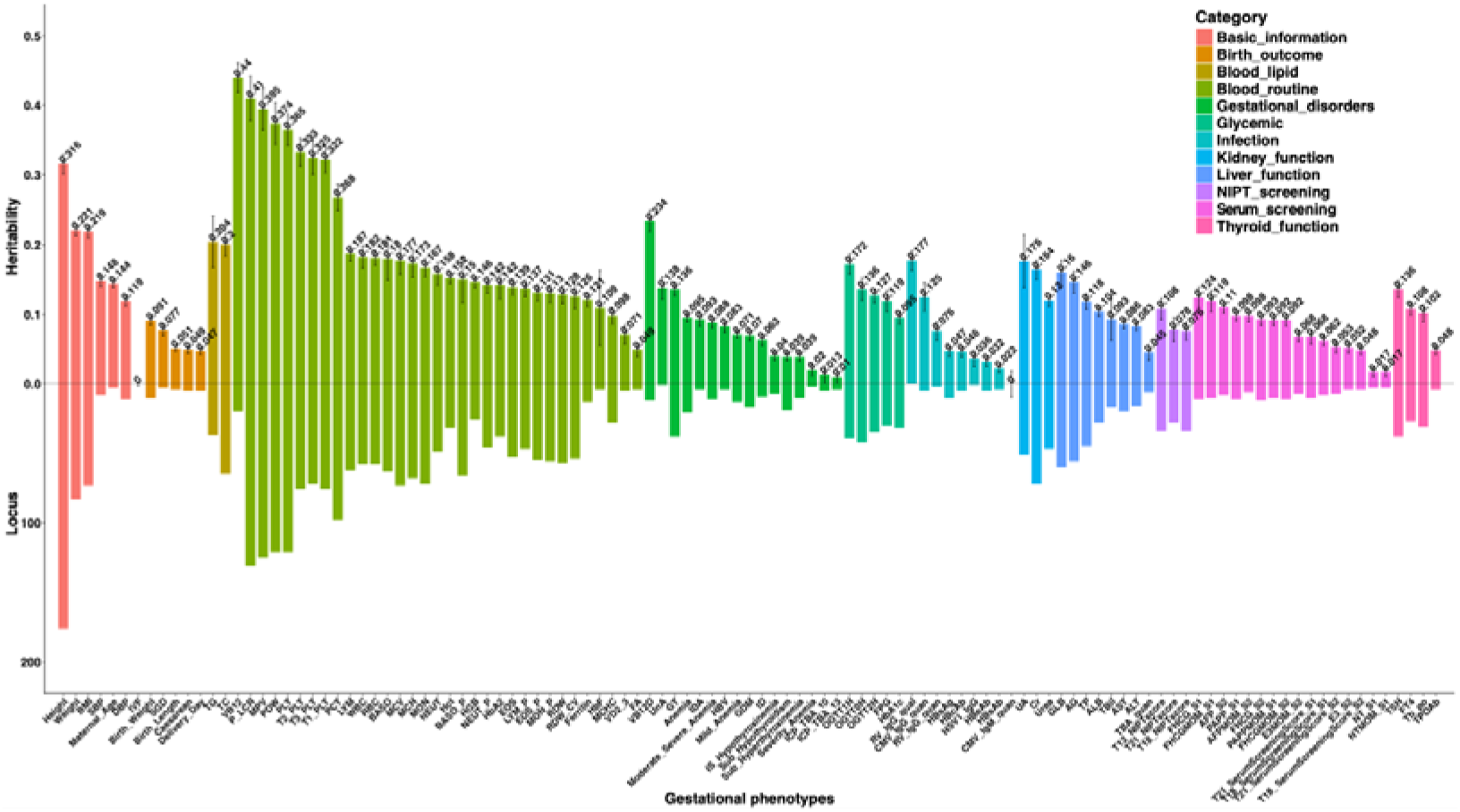
SNP Heritability and associated loci for all gestational phenotypes. Mirror plot displaying the estimated SNP heritability (above the zero line) and the number of genome-wide significant loci (below the zero line) for each gestational phenotype following meta-analysis. Negative heritability estimates have been set to zero for visualization.

**Extended Data Fig. 3.**
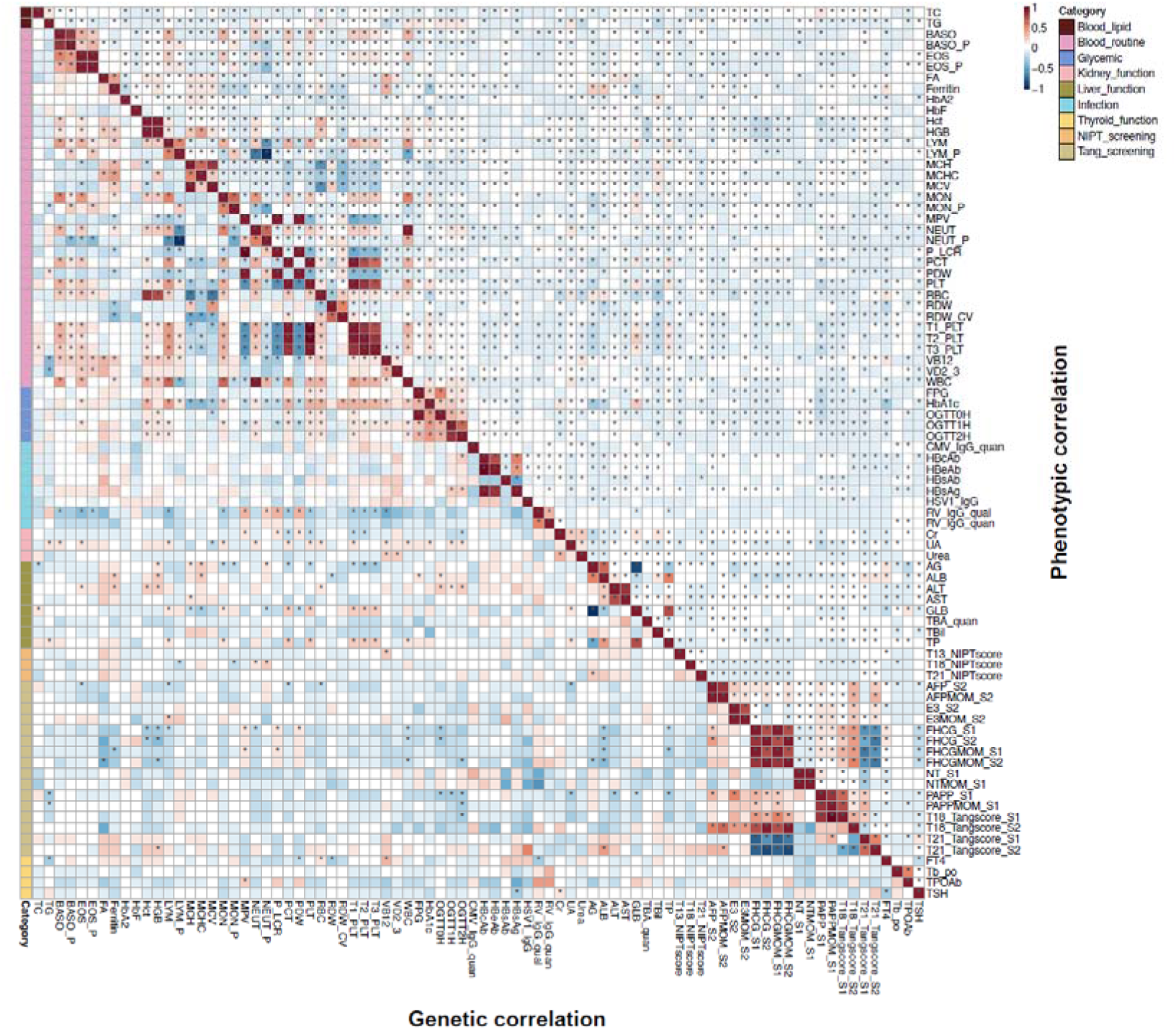
Phenotypic and genetic correlations among 82 molecular test phenotypes. Pairwise phenotypic (Spearman; upper triangle) and genetic (LDSC; lower triangle) correlation matrices for 82 molecular phenotypes, categorized into nine groups (one phenotype, CMV_IgM_quan, was excluded due to missing values). Asterisks (*) indicate significant correlations after Bonferroni correction (P < 0.05/(82×81)). Strong correlations were typically clustered within categories, though significant cross-category associations were also observed (e.g., blood cell counts showed widespread correlations with traits from other groups).

**Extended Data Fig. 4.**
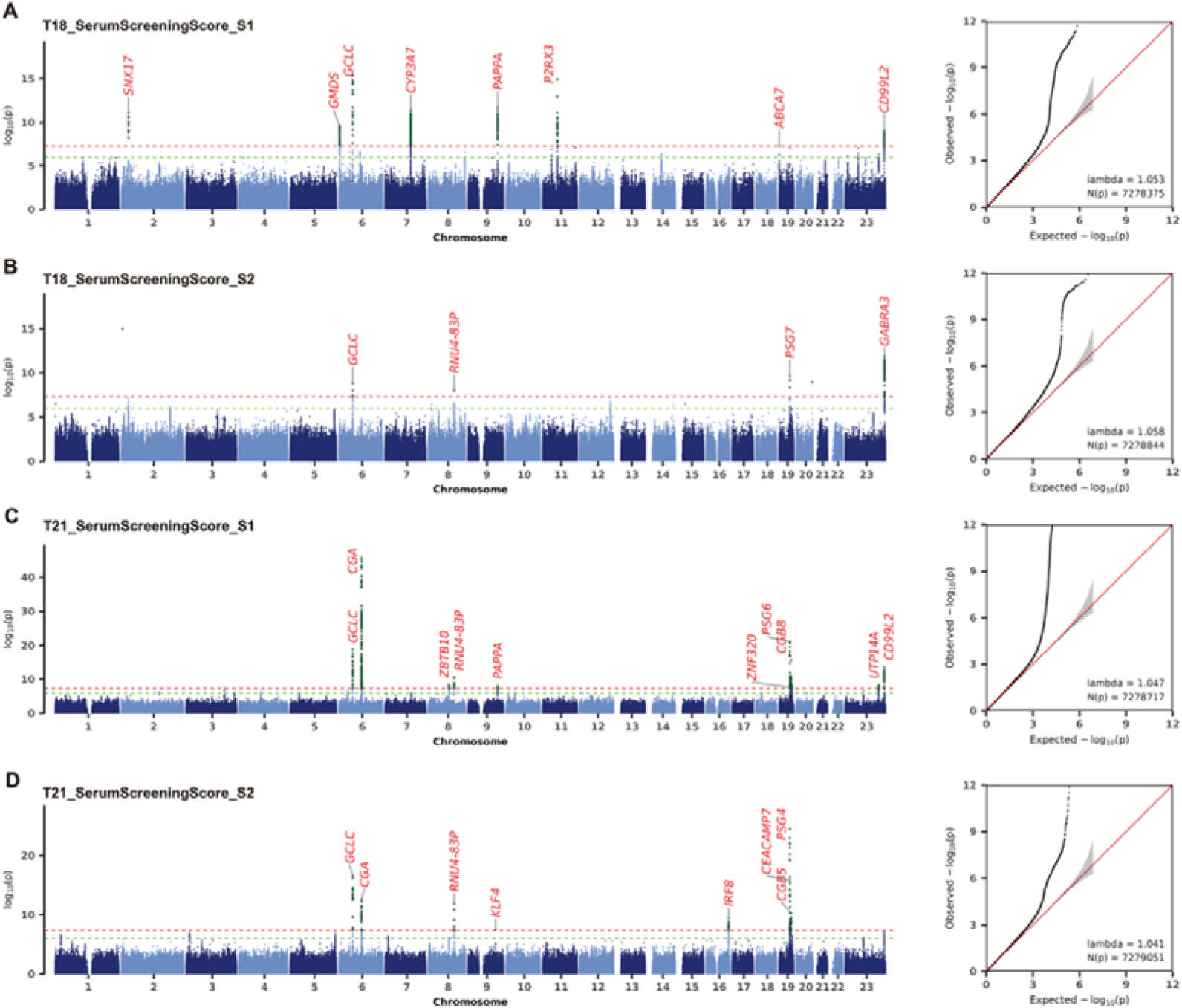
Manhattan and QQ plots from GWAS meta-analysis of prenatal serum screening scores. Manhattan plots show genome-wide association results for (a) first-trimester trisomy 18 risk score (T18_SerumScreeningScore_S1), (b) second-trimester trisomy 18 score (T18_SerumScreeningScore_S2), (c) first-trimester trisomy 21 risk score (T21_SerumScreeningScore_S1), and (d) second-trimester trisomy 21 score (T21_SerumScreeningScore_S2). The x axis indicates chromosomal position; the y axis shows –log10(*P*). Green and black dashed horizontal lines mark genome-wide significance (5 × 10^−8^) and suggestive (1 × 10^−6^) thresholds, respectively. Variants in loci previously reported in the GWAS Catalog are coloured black; novel loci are red. Corresponding QQ plots compare observed –log10(*P*) (points) against the null expectation (red line); the grey band indicates standard error.

**Extended Data Fig. 5.**
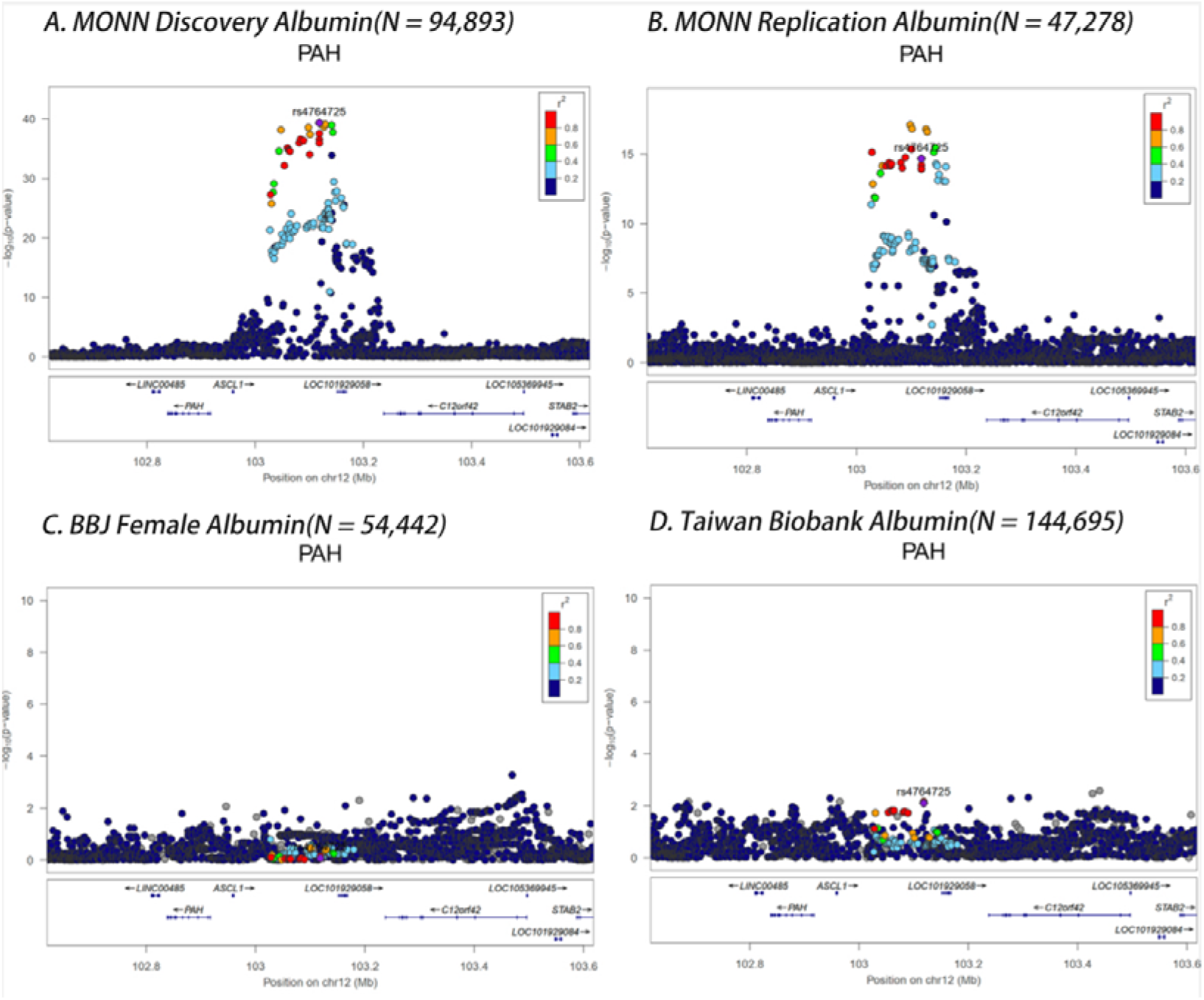
Regional association plot for the gestation-specific albumin locus (rs4764725-C). LocusZoom plots display the association signals across genomic regions surrounding the lead variant rs4764725□C in (A) the discovery GWAS from this study, (B) the independent replication cohort, (C) female participants from the BioBank Japan (BBJ), and (D) the Taiwan Biobank. Each plot shows −log10(*P*) values of SNPs (y-axis) against chromosomal position (x□axis), with color indicating linkage disequilibrium (r²) relative to the lead variant (purple diamond).

**Extended Data Fig. 6.**
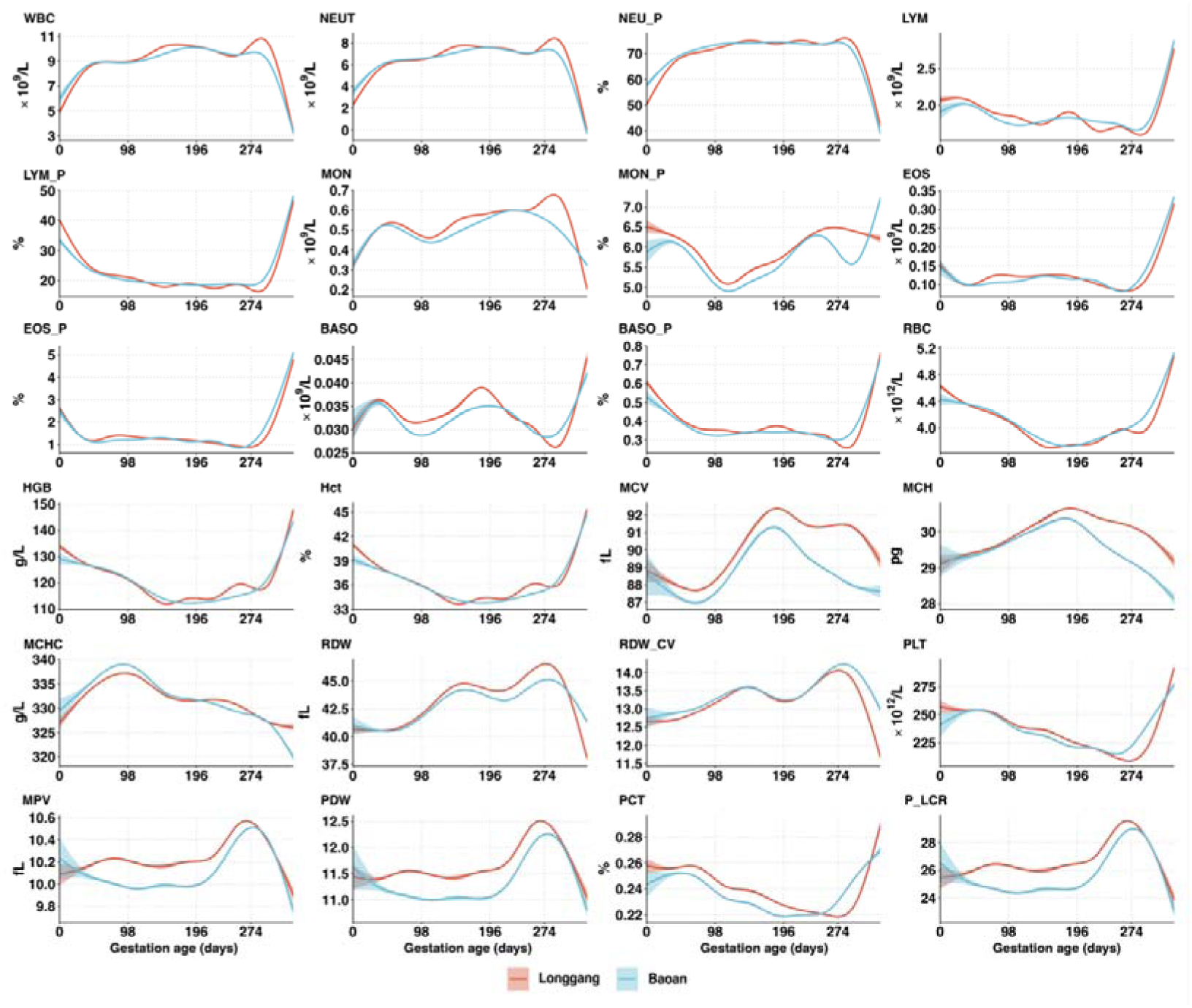
Longitudinal trajectories of 24 complete blood count phenotypes. Values are plotted against days from conception to 42 days postpartum for the Baoan (blue) and Longgang (orange) hospital cohorts. Vertical dashed lines delineate clinical trimesters: first (0–90 days), second (91–180 days), third (181–280 days), and delivery (>280 days).

**Extended Data Fig. 7.**
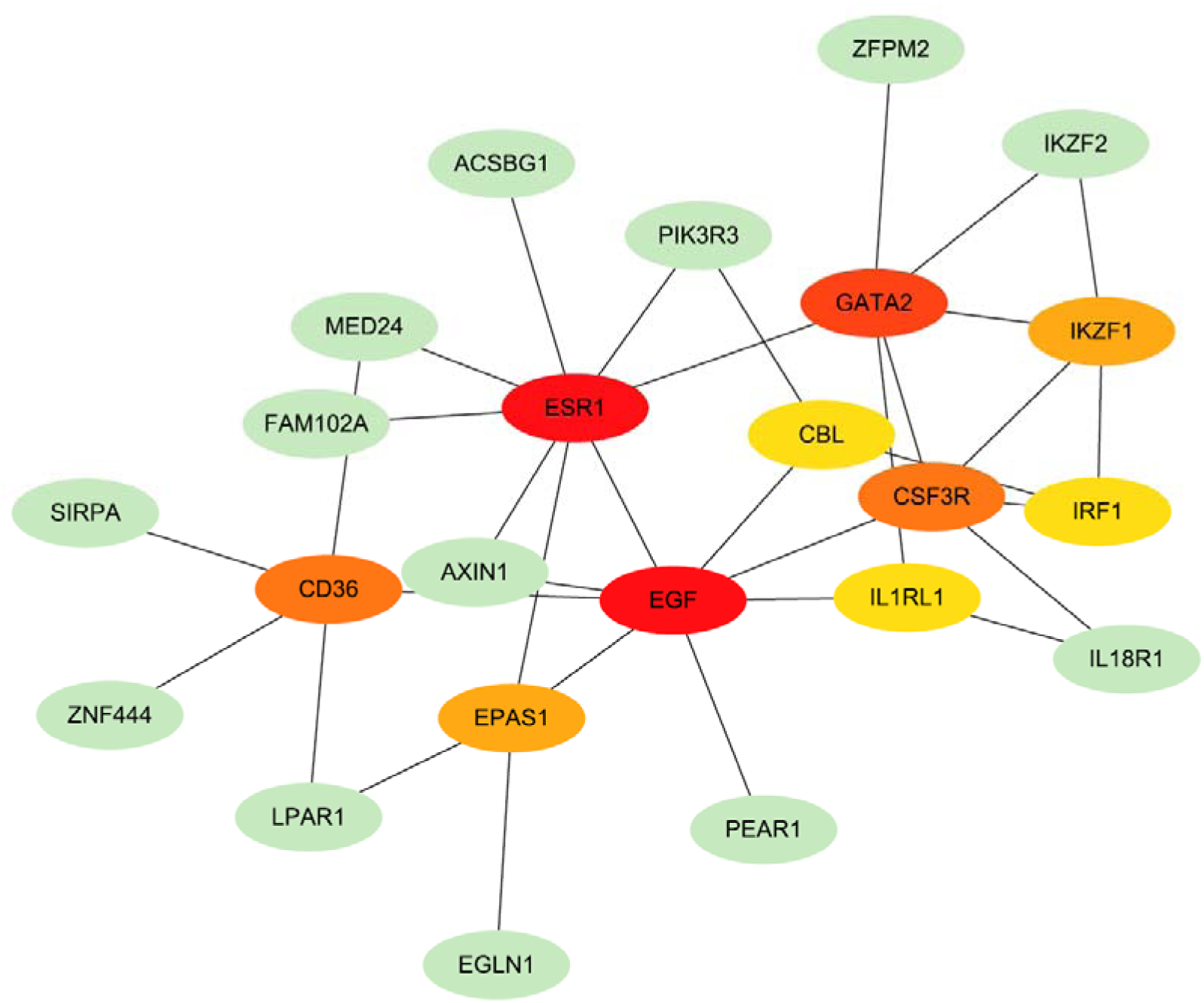
Protein-protein interaction network for loci with significant genotype-by-time (G×T) effects. Each node represents a gene mapped from a locus with significant G×T interaction. Hub genes are highlighted in red (highest degree), orange, and yellow.

**Extended Data Fig. 8.**
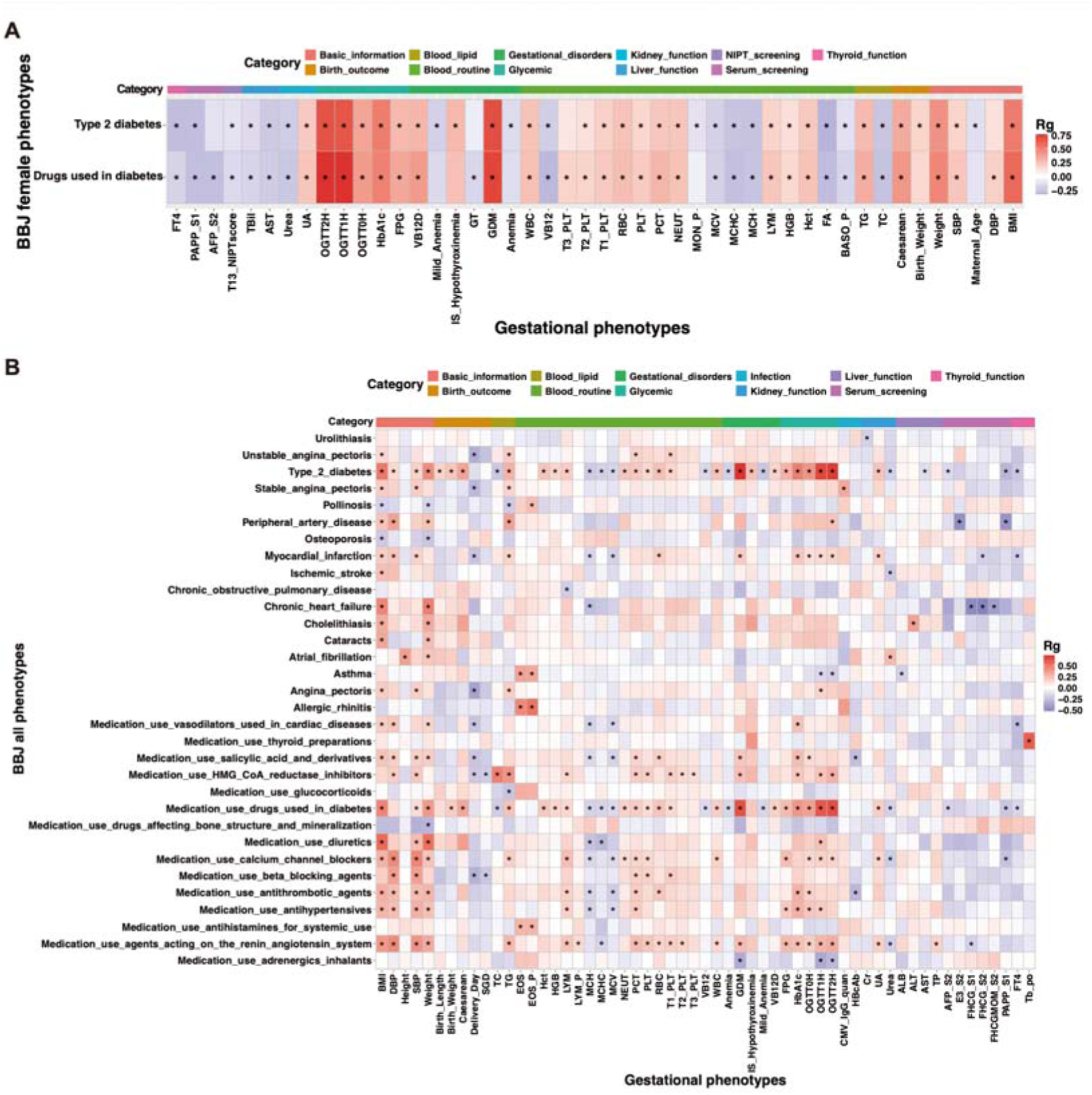
Genetic correlations between gestational phenotypes and common disorders and medication usage. Genetic correlation (rg) matrices of 111 gestational phenotypes with 80 BioBank Japan (BBJ) common disorders and medication use conditions for (A) the female-only population and (B) the complete (mixed-sex) population. Only pairs with at least one significant genetic correlation (false discovery rate, FDR q < 0.05) are displayed. Asterisks indicate correlations that meet the significance threshold (FDR q < 0.05). Full definitions of gestational phenotype abbreviations are provided in Supplementary Table 2.

**Extended Data Fig. 9.**
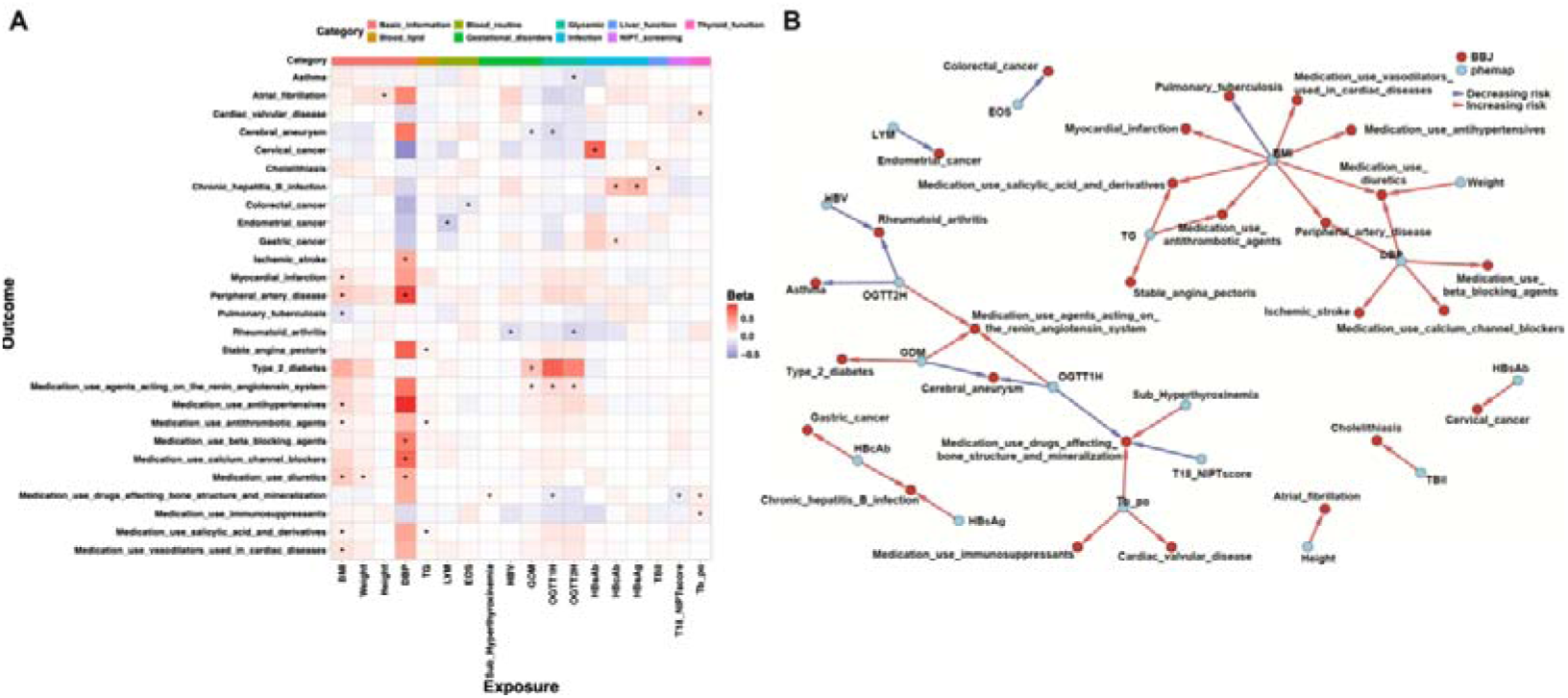
Mendelian randomization of gestational traits with common disorders and medication use in the complete BBJ participants. a, Matrix of genetic correlations (rg) between 111 gestational phenotypes and 80 later-life disorders and medication-use traits from the BioBank Japan (BBJ) cohort. b, Corresponding Mendelian randomization (MR) estimates (beta) for the same trait pairs. Only pairs with at least one significant genetic correlation (FDR q < 0.05) are shown. Asterisks denote significance after false discovery rate correction (FDR q < 0.05).

## Notes

### Competing Interest Statement

The authors have declared no competing interest.

### Funding Statement

The study was supported by Shenzhen Basic Research Foundation (20220818100717002), Guangdong Basic and Applied Basic Research Foundation (2022B1515120080, 2020A1515110859), National Natural Science Foundation of China (31900487), and the Shenzhen Health Elite Talent Training Project.

### Author Declarations

This study was approved by the Medical Ethics Committee of the School of Public Health (Shenzhen), Sun Yat-sen University, Longgang District Maternity and Child Healthcare Hospital of Shenzhen City, and Shenzhen Baoan Women's and Children's Hospital. Data collection was approved by the Human Genetic Resources Administration of China (HGRAC).

### Summary of Updates

1. Title was revised. 2. Author list was updated.

