## Supplementary Information for "A Dynamic Genetic Atlas of Gestational Phenotypes"

Supplementary Notes

### 1. Comparison with prior studies on gestational phenotypes

#### (1). Comparison with Guo et al. (PMID: 39389020):

Guo et al. investigated 26 phenotypes related to maternal comorbidities and child health in 25,639 pregnant Chinese women. Their GWAS phenotypes were defined using ICD-10 class I codes (e.g. O99, E02, N81), which do not capture specific clinically meaningful gestational disorders or birth outcomes, such as gestational diabetes, chronic hepatitis B infection, or birth weight, as examined in our study.

To enable comparison, we approximately mapped our maternal and child phenotypes to theirs (e.g., chronic hepatitis B infection mapped to ICD-10 code B18, "Certain infectious and parasitic diseases"). Only two overlapping phenotypes were identified, and of the 4,009 genome-wide significant associations discovered in our study, just two were reported by Guo et al (**Supplementary Table 8**). This underscores the novelty and significance of our findings. The reliance on broad ICD-10 Class I codes in Guo et al. likely reflects power limitations associated with their smaller sample size; prior work indicates that at least 30,000 participants are needed to obtain robust and reliable genetic effect estimates (PMID: 39389018).

#### (2). Comparison with Studies by Han et al. (PMID: 39389017) :

Han et al. analyzed 50 of the 111 gestational phenotypes included in our study, based on 20,900 Chinese women. Their smaller sample size (<1/5 of ours) substantially limited statistical power. Han et al. reported 410 trait–locus associations, 71.7% of which were previously known. By contrast, our study identified 4,009 associations, with 1,536 (38.3%) representing novel findings. None of these novel loci had SNPs in linkage disequilibrium ($R^2 > 0.2$) reported in the GWAS catalog or in the studies by Han et al. and Guo et al.

For example, in a GWAS of maternal height (**Supplementary Fig. 1**), we identified 210 independent genome-wide significant loci, whereas Han et al. identified only four. Our LDSC ratio was lower, reflecting more precise genetic effect estimates (**Supplementary Fig. 1**). These differences further highlight the impact of sample size on discovery power, consistent with prior evidence that ≥30,000 participants are needed for robust GWAS findings (PMID: 39389018).

### 2. Properties of GWAS Effect Estimates and Hypothesis Testing in Female-Only Versus Mixed-Sex Populations

As GWAS conducted for gestational phenotypes are restricted to female-only participants, we would like to document the properties of genetic effect estimates for these GWAS compared to a mixed-sex population GWAS across the general population, such as Taiwan and Japan Biobanks.

#### A. Impact of Model Specification on Effect Size Estimates

Model specification differences arise when distinct sets of covariates are included in GWAS models, resulting in phenotypes being analyzed on different residual variance scales. In our NIPT-based MONN GWAS, all participants were women; thus, no adjustment for sex was required:

$$y = a + \beta_{MONN} \cdot snp + e \quad (1)$$

In contrast, the Taiwan Biobank GWAS (PMID: 38116116) included sex as a covariate:

$$y = a + \beta_1 \cdot snp + \beta_2 \cdot sex + e \quad (2)$$

For both models, both SNP and sex are standardized to have zero mean and unit variance, respectively.
After standardization for phenotype y, these two models yield systematically different SNP effect estimates. In a female-only sample,

$$var(y_{MONN}) = 1 = h^2 + \sigma_E^2, \text{ with } \hat{\sigma}_b \approx \sqrt{\frac{\sigma^2_{y_{MONN}}}{N * 2 * p * (1-p)}}.$$

whereas in a mixed-sex sample,

$$var(y_{TW}) = 1 = \tilde{h}^2 + \tilde{\sigma}^2_{sex} + \tilde{\sigma}^2_E, \text{ and } \hat{\sigma}_b \approx \sqrt{\frac{1 - c^2 * var(sex)}{N * 2 * p * (1-p)}}.$$

Thus,

$$\beta_{TW} \sim N\left(0, \frac{\tilde{h}^2}{\tilde{h}^2 + \tilde{\sigma}^2_{sex} + \tilde{\sigma}^2_E}\right), \quad \beta_{MONN} \sim N\left(0, \frac{h^2}{h^2 + \sigma_E^2}\right),$$

Implying

$$|\beta_{MONN}| > |\beta_{TW}|$$

for the same SNP variant.

#### B. Effect on GWAS Test Statistics

Although model specification modifies effect size magnitudes, the GWAS test statistic and p-value remain asymptotically unchanged, because the change in the numerator (β) is offset by a proportional change in its standard error.

Using standard GWAS derivations, we outline the proof below.

Consider a GWAS of $N$ individuals in which the phenotype was determined by a total of $m$ causal SNPs.

***Female-only model (Model 1; MONN)***

For SNP $i$,

$$\beta_i = \frac{cov(x_i, y)}{var(x_i)}$$

Under the polygenic assumption, $\beta_i \sim N(0, \frac{h^2}{m})$, so $E(\beta_i) = \pm\sqrt{\frac{h^2}{m}}$

The standard error is

$$se(\beta_i) = \sqrt{\frac{\sigma_y^2 - \hat{\beta}_i^2}{(N-1)}}$$

Because each single SNP explains only a negligible proportion of phenotypic variance and $\sigma_y^2$ is scaled to unit,

$$se(b_i) = \sqrt{\frac{\sigma_y^2 - \hat{\beta}_i^2}{(N-1)}} \approx \sqrt{\frac{1}{(N-1)}}$$

Under the null hypothesis $\beta_i = 0$, the test statistic of a GWAS becomes

$t_1 = \frac{\hat{\beta}_i}{se(b_i)}$, which has expectation $E(t_1) = \pm(N-1)\sqrt{\frac{h^2}{m}}$.

***Mixed-sex model with sex covariate (Model 2)***

When sex is included as an additional covariate (Model 2),

$$\boldsymbol{\beta} = (X^T X)^{-1} X^T Y = \frac{1}{N}\begin{bmatrix}1 & 0\\ 0 & 1\end{bmatrix}^{-1}\binom{snp \cdot y_n}{sex \cdot y_n} = \frac{1}{N}\begin{bmatrix}1 & 0\\ 0 & 1\end{bmatrix}^{-1}\binom{\rho_{snp \cdot y}}{\rho_{sex \cdot y}}$$

In which $\boldsymbol{\beta} = [\beta_1, \beta_2]^T$ for SNP effect ($\beta_1$) and sex effect ($\beta_2$). The SNP effect becomes

$$\beta_1 = \rho_{snp \cdot y}$$

And under polygenic assumption, $\rho_{snp \cdot y} \sim N(0, \frac{1}{m} \cdot \frac{\tilde{h}^2}{\tilde{h}^2 + \tilde{\sigma}_{sex}^2 + \tilde{\sigma}_E^2})$

So, $E(\beta_1) = \pm\sqrt{\frac{1}{m} \cdot \frac{\tilde{h}^2}{\tilde{h}^2 + \tilde{\sigma}_{sex}^2 + \tilde{\sigma}_E^2}} = \pm\sqrt{\frac{1}{m} \cdot \tilde{h}^2 (1 - {\beta_2}^2)}$.

The corresponding standard error is

$$se(\beta_1)=\sqrt{\frac{\sigma_y^2-\beta_1{}^2-\beta_2{}^2}{(N-2)}}\approx\sqrt{\frac{1-\beta_2{}^2}{N-2}}$$

Thus, under the null hypothesis,

$$t_2=\frac{b_i}{se(b_i)}$$

which has expectation $E(t_2)=\frac{\sqrt{\frac{1}{m}\cdot\tilde{\mathrm{h}}^2(1-\beta_2{}^2)}}{\sqrt{\frac{1-\beta_2{}^2}{N-2}}}=\pm(N-2)\sqrt{\frac{\mathrm{h}^2}{m}}$.

#### *Asymptotic equivalence of test statistics*

It is easy to see that for large sample sizes,

$$t_1\approx t_2.$$

Therefore, although including sex as a covariate reduces SNP effect sizes by absorbing between-sex phenotypic variance, the GWAS test statistic, t-statistics here, and their p-values remain effectively unchanged.

#### *Empirical validation*

To empirically validate these model-based inferences, we analyzed two independent datasets. First, we compared genetic effect estimates from the MONN NIPT maternal height GWAS with those from the BioBank Japan (BBJ) height GWAS conducted in the full cohort (N = 165,056) and in female participants only (N = 75,797). Consistent with expectation, effect sizes were systematically larger in the BBJ female-only and MONN NIPT analyses than in the mixed-sex BBJ analysis (**Supplementary Fig. 4**).

Second, we analyzed publicly available GWAS summary statistics for type 2 diabetes (T2D) (PMID: 32499647), which report results for both mixed-sex and female-only cohorts. Using genome-wide significant loci identified in the female-only analysis (Supplementary Table 6 of PMID: 32499647), we compared SNP effect estimates from the female-only GWAS (x-axis) with those from the mixed-sex GWAS (y-axis) under both BMI-adjusted (**Supplementary Fig. 5a**) and BMI-unadjusted (**Supplementary Fig. 5b**) models. In both settings, effect sizes were consistently larger in the female-only analyses, in agreement with the predicted shrinkage of genetic effects in mixed-sex models.

### 3. Properties of Identifying Gestation-Specific Genetic Effects using Female-Only Versus Mixed-Sex GWAS

With respect to the identification of gestation-specific genetic effects, we emphasize that our classification framework is based on a Bayesian clustering approach combined with colocalization analysis (Methods: "Detection of gestation-specific genetic effects"). Both methods rely on relative patterns of association and signal sharing across conditions rather than on absolute effect size magnitudes. Therefore, in principle, systematic scaling differences in β estimates should not materially influence the classification results.

To empirically assess this assumption, we conducted a comparative analysis between gestational diabetes mellitus (GDM) GWAS results from our MONN cohort and type 2 diabetes GWAS conducted in East Asian populations including (i) all participants and (ii) female-only participants (**Supplementary Fig. 15**). Among the 19 loci associated with GDM, classification concordance was observed for 17 loci across models. Only two loci showed minor discrepancies: FOXA2, which was classified as pregnancy-specific in the female-only analysis but unclassified in the mixed-sex analysis, and GCKR, which was classified as general in the mixed-sex analysis but unclassified in the female-only analysis. These limited differences suggest that GWAS model specification has minimal impact on the overall classification of gestation-specific versus general loci.

Although, in theory, the clustering results are not expected to differ substantially when using a female-only GWAS versus a mixed-sex GWAS for comparison with the MONN GWAS, restricting analyses to female-only participants minimizes potential confounding from sex-specific genetic effects when inferring gestation-specific associations.

### 4. Properties of MR Effect Estimates and Hypothesis Testing in Female-Only Versus Mixed-Sex Populations

We next demonstrate that although differences in GWAS model specification induce systematic changes in SNP effect size estimates, the corresponding $t$ statistics and $p$ values for Mendelian randomization (MR) hypothesis testing remain unchanged.

MR estimates were obtained using the Wald ratio for each SNP, defined as the ratio of the SNP-outcome association ($\widehat{\beta_{Y_j}}$ obtained from BBJ) to the SNP-exposure association ($\widehat{\beta_{X_j}}$ from our MONN GWAS). The overall causal effect ($\theta$) was estimated using inverse-variance weighted (IVW) regression:

$$\widehat{\beta_{Y_j}} = \theta\widehat{\beta_{X_j}} + \varepsilon_j \; (E1)$$

In the exposure GWAS, $\widehat{\beta_{X_j}}$ may arises from two alternative model specifications:

$$\begin{cases} y = \widehat{\beta_{X_j}} X_j + e \; (female-only\ model) \\ y = \widetilde{\beta_{X_j}} X_j + \beta_s Sex + e \; (mixed-sex\ model) \end{cases}$$

As shown in "B.Effect on GWAS Test Statistics", inclusion of sex as a covariate leads to a shrinkage of SNP effect estimates by a factor

$$\eta = \sqrt{1-\beta_s^2}$$

such that

$$\widetilde{\beta_{X_j}} = \sqrt{1-\beta_s^2}\,\widehat{\beta_{X_j}} = \eta\widehat{\beta_{X_j}}$$

Substituting these expressions into equation E(1), the IVW estimator and its standard error are given by the following.

- **Female-only model:**

$$\begin{cases} \theta_1 = \dfrac{\sum_{j=1}^{J} \widehat{\beta_{X_j}}\widehat{\beta_{Y_j}} se(\widehat{\beta_{Y_j}})^{-2}}{\sum_{j=1}^{J} \widehat{\beta_{X_j}}^2 se(\widehat{\beta_{Y_j}})^{-2}} \\ se(\theta_1) = \sqrt{\dfrac{1}{\sum_{j=1}^{J} \widehat{\beta_{X_j}}^2 se(\widehat{\beta_{Y_j}})^{-2}}} \end{cases}$$

Under the null hypothesis $\theta_1 = 0$, the test statistic is

$$t_1 = \frac{\theta_1}{se(\theta_1)} = \frac{\sum_{j=1}^{J} \widehat{\beta_{X_j}}\widehat{\beta_{Y_j}} se(\widehat{\beta_{Y_j}})^{-2} / \sum_{j=1}^{J} \widehat{\beta_{X_j}}^2 se(\widehat{\beta_{Y_j}})^{-2}}{\sqrt{\dfrac{1}{\sum_{j=1}^{J} \widehat{\beta_{X_j}}^2 se(\widehat{\beta_{Y_j}})^{-2}}}}$$

- **Mixed-sex model where $\widetilde{\beta_{X_j}} = \eta\widehat{\beta_{X_j}}$**

$$\begin{cases} \theta_2 = \dfrac{\sum_{j=1}^{J} \eta\widehat{\beta_{X_j}}\widehat{\beta_{Y_j}} se(\widehat{\beta_{Y_j}})^{-2}}{\sum_{j=1}^{J} \eta^2\widehat{\beta_{X_j}}^2 se(\widehat{\beta_{Y_j}})^{-2}} = \dfrac{\sum_{j=1}^{J} \eta\widehat{\beta_{X_j}}\widehat{\beta_{Y_j}} se(\widehat{\beta_{Y_j}})^{-2}}{\eta\sum_{j=1}^{J} \widehat{\beta_{X_j}}^2 se(\widehat{\beta_{Y_j}})^{-2}} = \dfrac{\theta_1}{\eta} \\ se(\theta_2) = \sqrt{\dfrac{1}{\sum_{j=1}^{J} \eta^2\widehat{\beta_{X_j}}^2 se(\widehat{\beta_{Y_j}})^{-2}}} = \dfrac{1}{\eta}\sqrt{\dfrac{1}{\sum_{j=1}^{J} \widehat{\beta_{X_j}}^2 se(\widehat{\beta_{Y_j}})^{-2}}} = \dfrac{se(\theta_1)}{\eta} \end{cases}$$

Accordingly, under the null hypothesis $\theta_2=0$, the test statistic is:

$$t_2=\frac{\theta_2}{se(\theta_2)}=t_1$$

Therefore, although differences in GWAS model specification systematically alter the absolute magnitude of MR causal effect estimates ($\theta$), the statistical evidence for causality— quantified by the $t$ statistic and correspoinding $p$ value—remains invariant.

### 5. Definition of Maternal serum screening or trisomy 18/21 risk at first and second trimesters

Maternal serum screening does not have a single universal "formula" for trisomy 18/21 risk. Instead, **all first-trimester and second-trimester screening programs worldwide use the same general statistical framework**:

General Formula Used in All Screening Programs

The estimated risk is calculated as:

$$Posterior\ Risk = A\ Priori\ Risk \times Likelihood\ Ratio\ (LR)$$

Where:

- **A Priori Risk** = maternal age–based risk of T21/T18 at term
- **Likelihood Ratio** = the combined likelihood ratio generated from each serum marker, NT measurement (1st trimester), and ultrasound findings.

FIRST-TRIMESTER SCREENING (FTS)

Markers used:

- PAPP-A (Pregnancy-Associated Plasma Protein A)
- Free β-hCG
- Nuchal translucency (NT)

Each marker is converted to MoM (Multiples of the Median) adjusted for:

- gestational age
- maternal weight
- ethnicity
- smoking
- IVF/pregnancy factors

General Likelihood Ratio Calculation

For each marker:

$$LR_i = \frac{f(MoM|T21 \text{ or } T18)}{f(MoM|unaffected)}$$

where f(.) is is the log-Gaussian density function
(mean and SD differ for affected vs. unaffected pregnancies).

Then:

$$LR_{combined} = LR_{PAPP-A} \times LR_{\beta hCG} \times LR_{NT}$$

Finally:

$$Posterior\ Risk_{FTS} = Maternal\ Age\ Risk \times LR_{combined}$$

SECOND-TRIMESTER SCREENING (STS)
Markers used:

- AFP
- Total hCG
- uE3

Again, each marker → MoM → LR → combined risk:

$$LR_{combined} = LR_{AFP} \times LR_{hCG} \times LR_{uE3}$$

$$Posterior\ Risk_{STS} = Maternal\ Age\ Risk \times LR_{combined}$$

### 6. Definition of NIPT risk score

The model used for computing NIPT risk score is called the Chromosome Abnormality Detection (CAD) algorithm, built on:

- Proportional read-count normalization
- Z-score testing

#### (1) Normalized Chromosome Fraction

For each chromosome i:

$$F_i = \frac{R_i}{\sum_k R_k}$$

$R_i$ = uniquely mapped reads to chromosome i
denominator = total autosomal mapped reads

#### (2) Expected Chromosome Fraction (Euploid Reference)

From a validation set of euploid pregnancies (thousands of samples):

$$\mu_i = E(F_i)$$
$$\sigma_i = SD(F_i)$$

These depend on fetal fraction and GC correction.

#### (3) Z-score Calculation

This is the key statistic for the reports:

$$Z_i = \frac{F_i - \mu_i}{\sigma_i}$$

High $Z$ indicates excess counts, indicating possible trisomy.

### Supplementary Figures

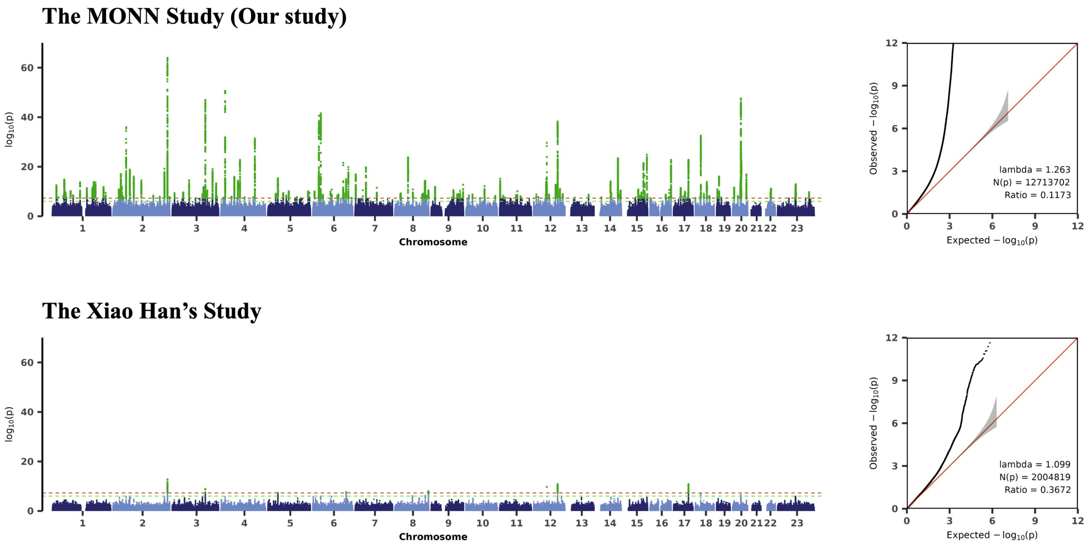

**Supplementary Fig. 1 | Comparison of genome-wide association study (GWAS) results for maternal height between the present study and the study by Xiao Han et al.**

Summary statistics from Xiao Han et al. were obtained from the original publication (PMID: 39389017).

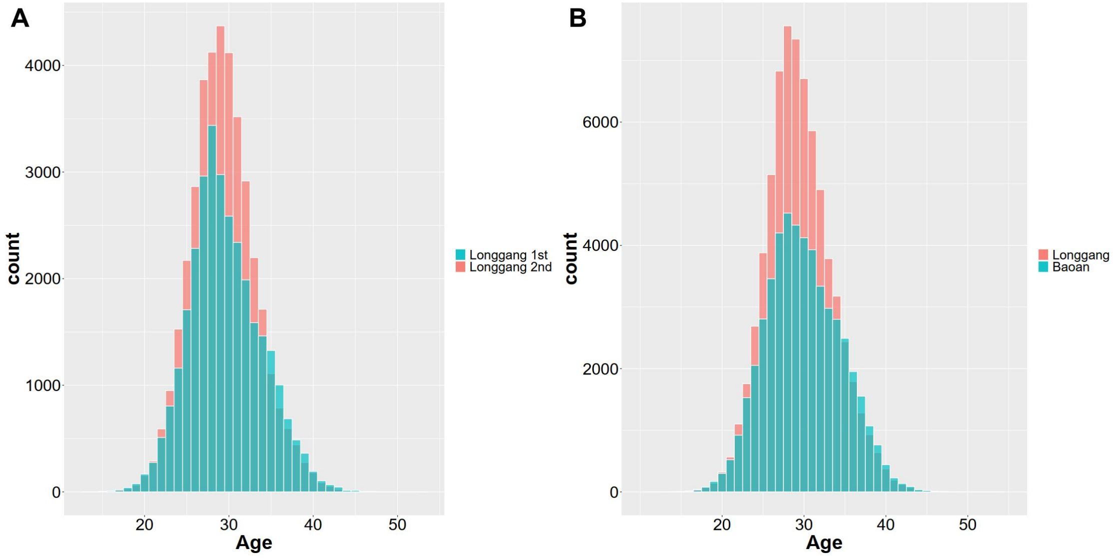

**Supplementary Fig. 2 | Age distribution of pregnant women recruited from two hospitals.**

a, Age distribution of participants in two cohorts from Longgang Hospital, collected between 2017–2019 and 2019–2022.

b, Age distribution of all pregnant women recruited from Baoan and Longgang hospitals.

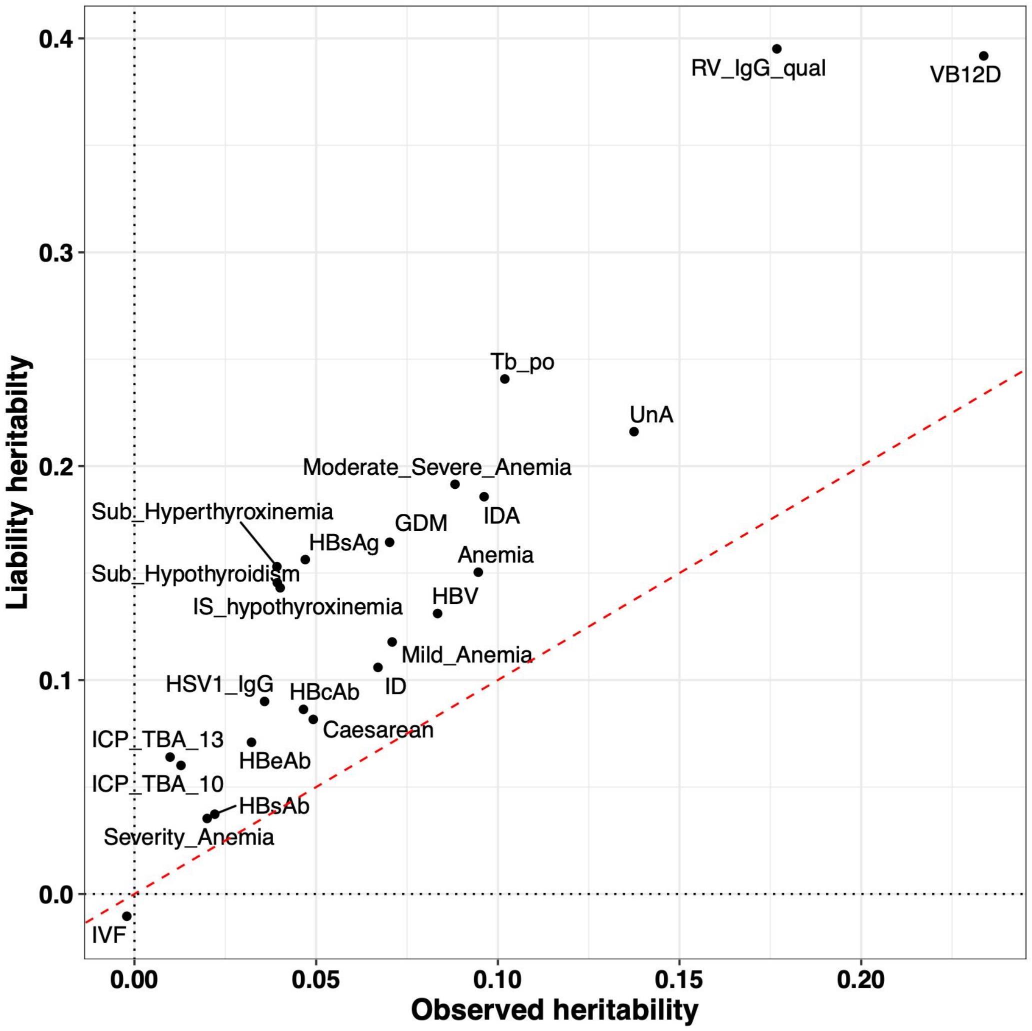

**Supplementary Fig. 3 | Comparison of SNP heritability estimates on the liability and observed scales for case–control phenotypes.**

SNP heritability was estimated using linkage disequilibrium score regression (LDSC).

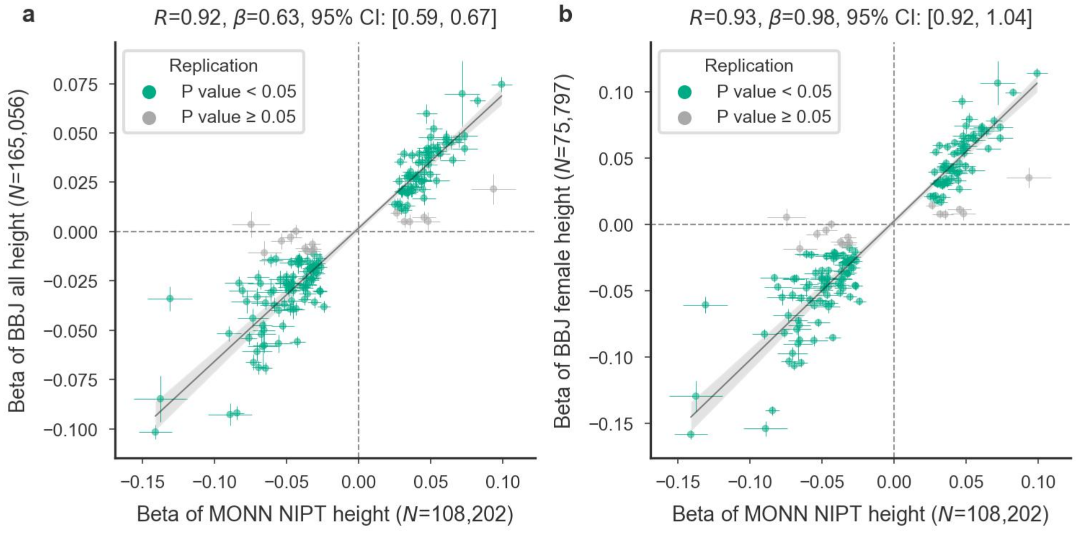

**Supplementary Fig. 4 | Comparison of genetic effect estimates for height between the MONN NIPT GWAS and BioBank Japan (BBJ).**

Effect estimates from the MONN GWAS (x axis) are plotted against those from the BBJ GWAS (y axis) for analyses including (a) all participants and (b) female participants only. $R$ denotes Pearson's correlation coefficient, and $\beta$ indicates the regression coefficient (slope of the grey line from regressing BBJ on MONN effect estimates).

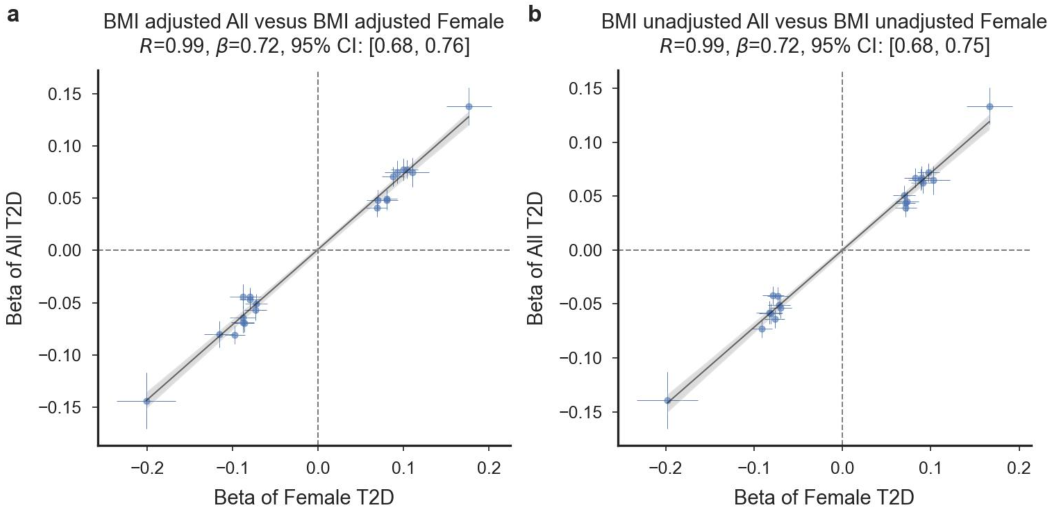

**Supplementary Fig. 5 | Comparison of genetic effect estimates between female-only and mixed-sex GWAS models for type 2 diabetes (T2D) in public datasets.**

Effect estimates from female-only GWAS are plotted against those from mixed-sex GWAS for analyses (a) adjusted for body mass index (BMI) and (b) unadjusted for BMI. Data were obtained from publicly available GWAS summary statistics (PMID: 32499647,https://blog.nus.edu.sg/agen/summary-statistics/t2d-2020/). $R$ denotes Pearson's correlation coefficient, and $\beta$ indicates the regression coefficient (slope of the grey line from regressing mixed-sex on female-only T2D effect estimates).

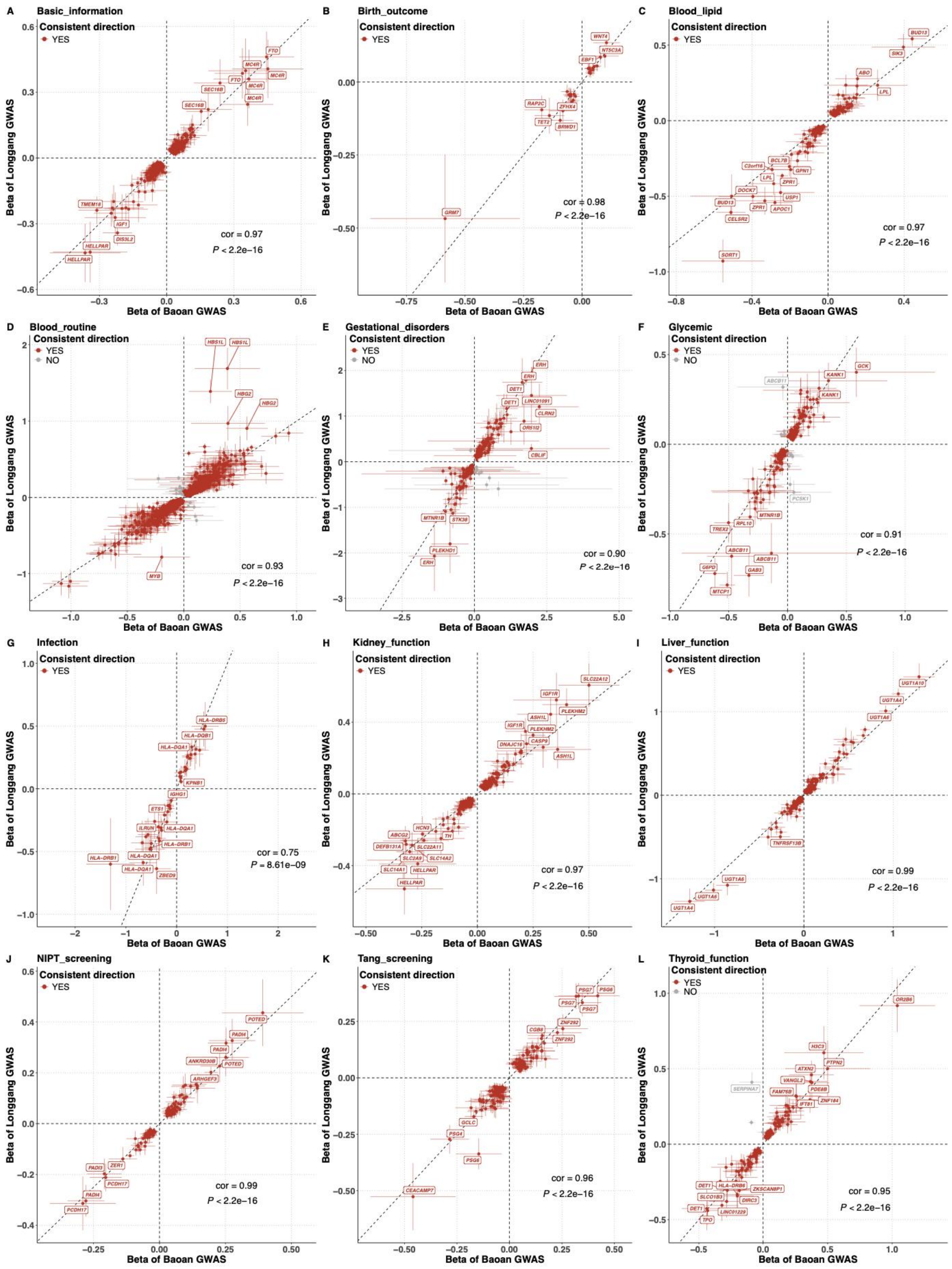

**Supplementary Fig. 6 | Concordance of genetic effect estimates between the two discovery hospitals.**

Effect sizes for independent lead variants are compared between hospitals for each phenotype. Pearson's correlation coefficients ($R$) and corresponding significance levels are shown.

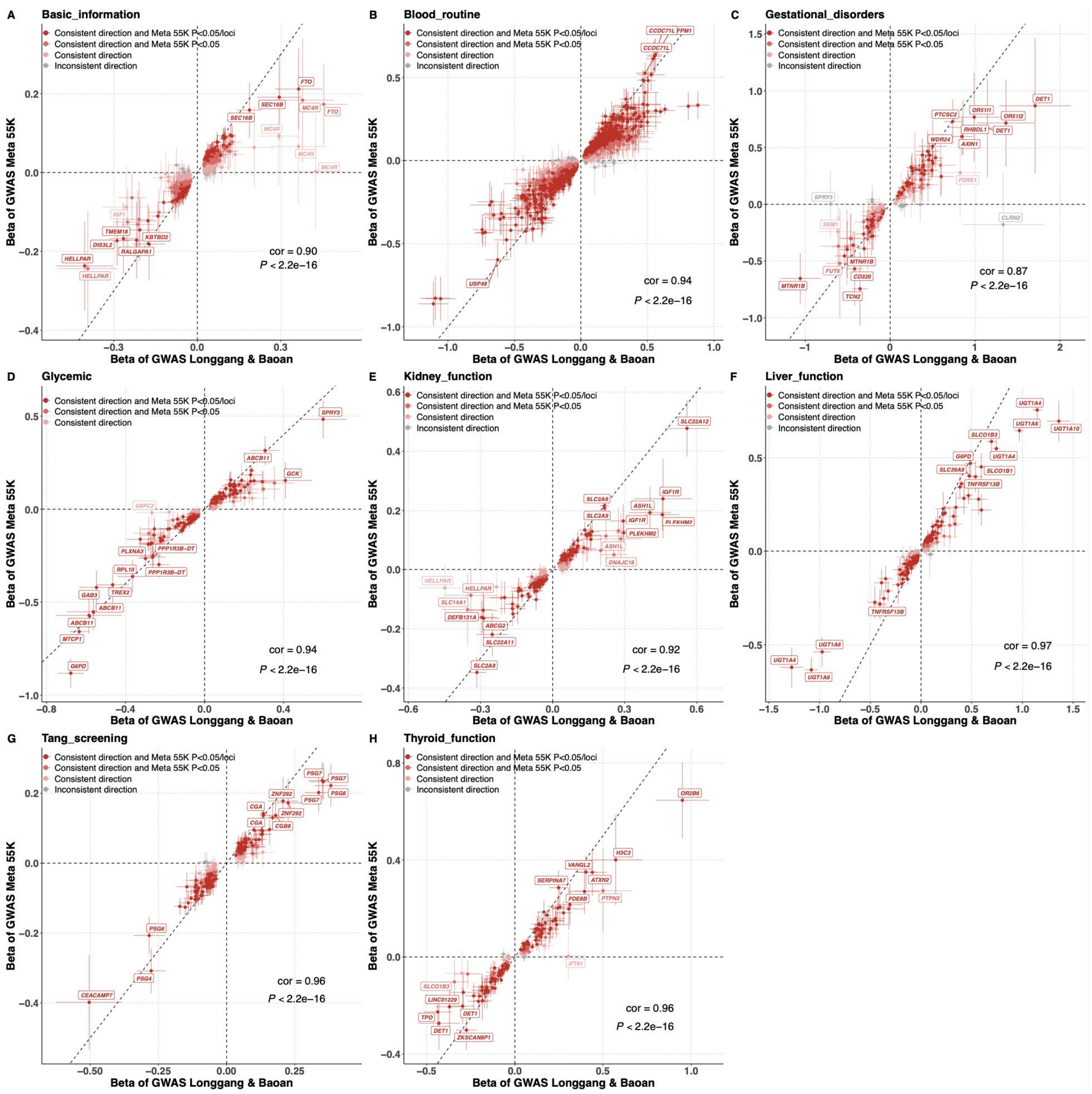

**Supplementary Fig. 7 | External replication of meta-GWAS results from the Baoan and Longgang cohorts in an independent 55K meta-analysis cohort.**

Scatter plots display effect sizes with 95% confidence intervals for phenotype-associated loci across eight phenotype categories. Replication significance was evaluated using Bonferroni correction and nominal thresholds. Pearson's correlation coefficients and corresponding *P* values are shown in the lower right corner.

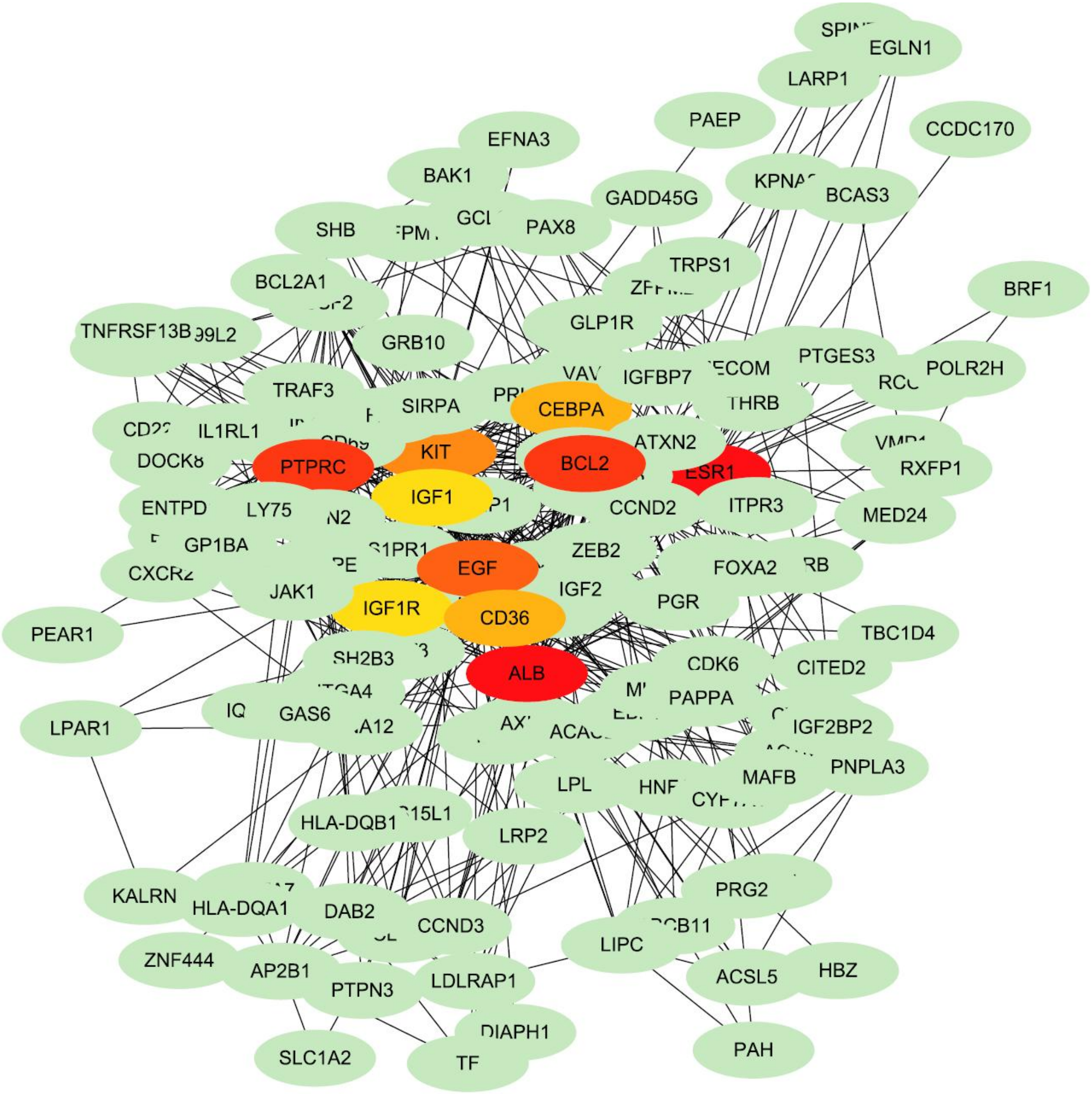

**Supplementary Fig. 8 | Protein–protein interaction (PPI) network of genes associated with multiple gestational phenotypes.**

Nodes represent genes listed in Supplementary Table 16, with yellow, orange and pink nodes indicating hub genes.

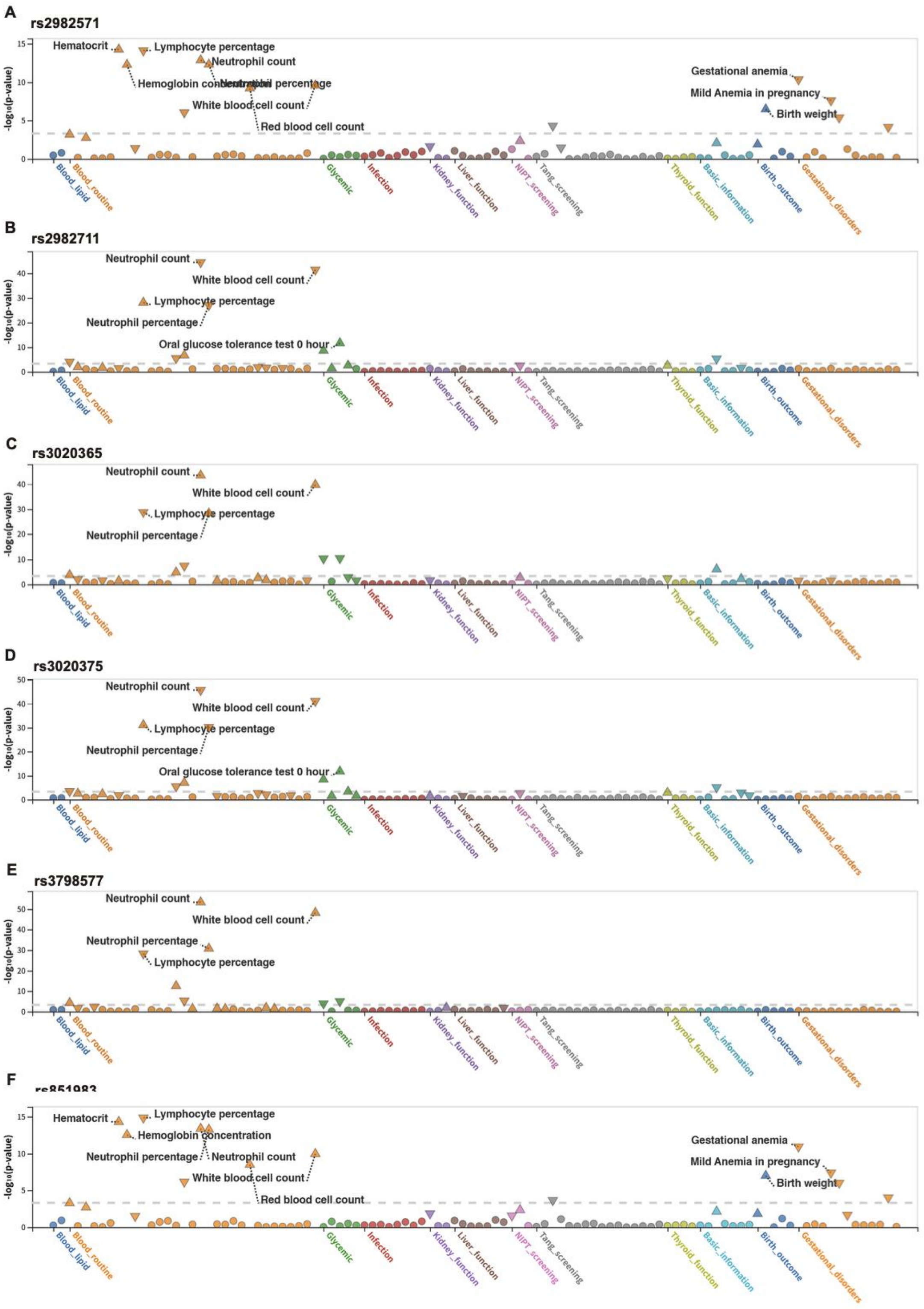

**Supplementary Fig. 9 | Phenome-wide association study (PheWAS) results for six variants at the ESR1 locus.**

Pleiotropic associations of six ESR1 variants are shown across a broad range of phenotypes.

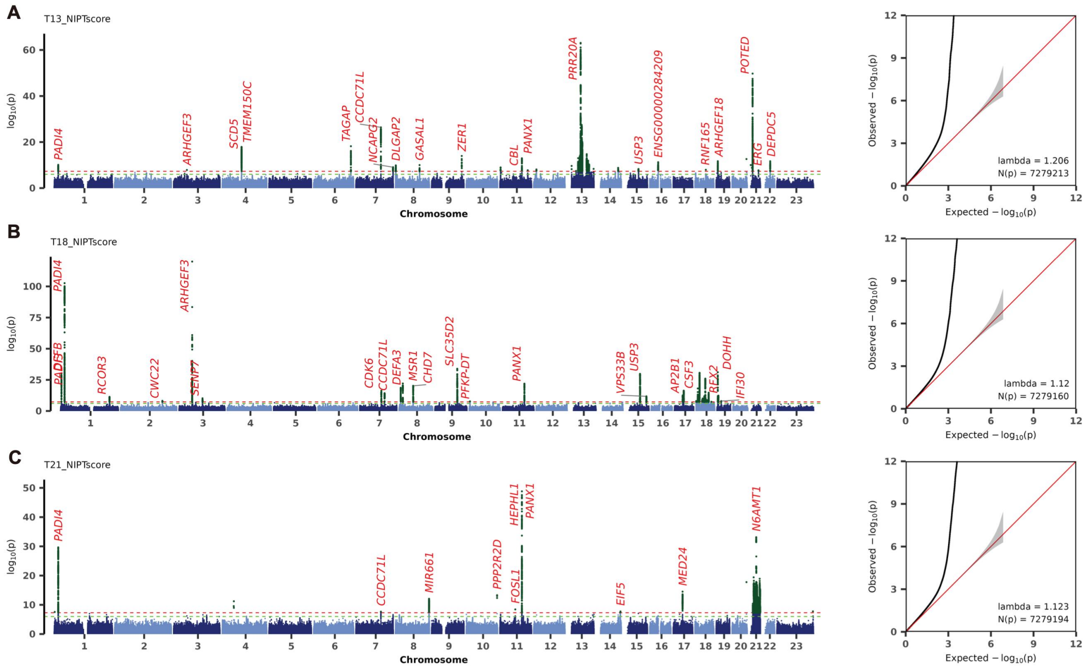

**Supplementary Fig. 10 | Manhattan and QQ plots of GWAS meta-analysis for three NIPT screening phenotypes.**

a–c, Manhattan plots show association results for trisomy 13 (T13_NIPTscore, a), trisomy 18 (T18_NIPTscore, b), and trisomy 21 (T21_NIPTscore, c). The x-axis represents chromosomal positions of SNPs, and the y-axis shows $-\log_{10}$(P-values). Genome-wide significance ($5\times10^{-8}$) and suggestive ($1\times10^{-6}$) thresholds are indicated by green solid and black dotted lines, respectively. Previously reported loci in the GWAS Catalog are colored black; novel loci are shown in red. QQ plots compare observed $-\log_{10}$(P-values) with the expected distribution under the null, with the red line representing expectation and the grey shaded area indicating standard errors.

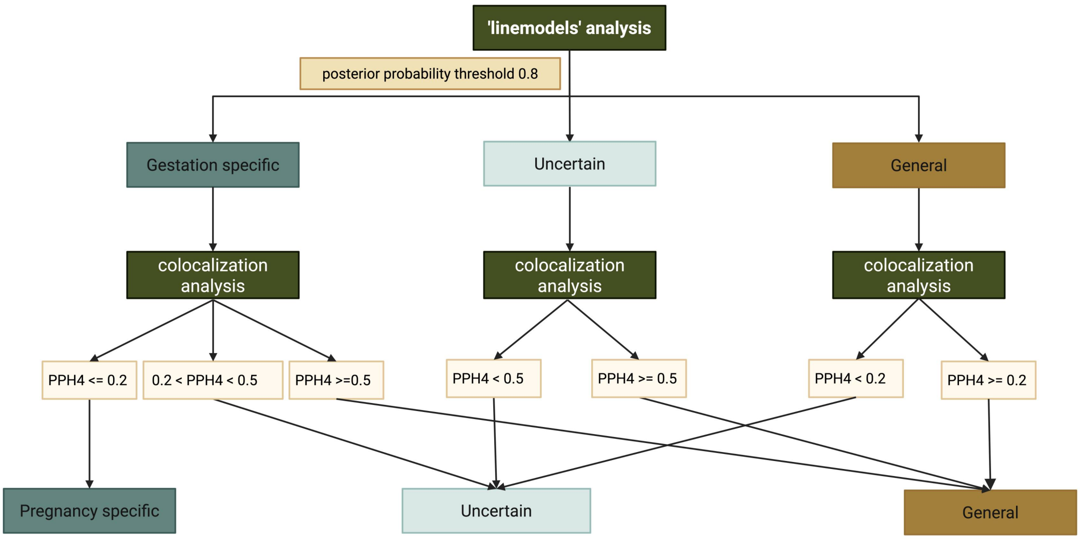

**Supplementary Fig. 11 | Schematic overview of variant classification.**

Variant effects were categorized as gestation-specific, general, or uncertain based on linemodel and colocalization analyses. A detailed description of the classification scheme is provided in the Methods section, “Detection of gestation-specific genetic effects.”

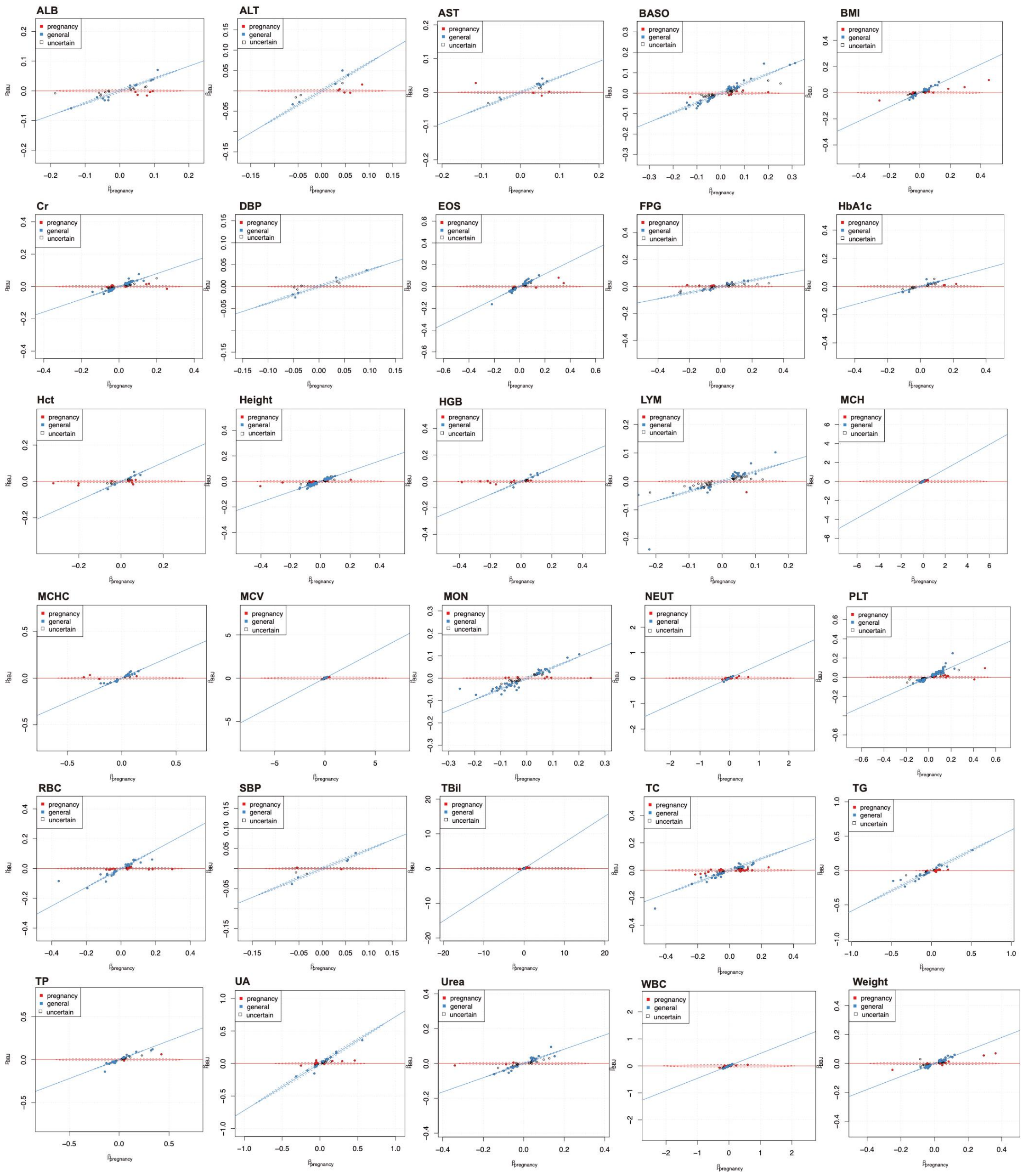

**Supplementary Fig. 12 | Line model analysis of genetic effects for 30 phenotypes during pregnancy and in female participants from BBJ.**

Scatterplots depict Bayesian classifier results for shared variants. Two clusters represent gestation-predominant effects (red) and general effects (blue), while gray SNPs have posterior probability ≤80% and are unassigned. Dotted ellipses indicate 80% probability regions for each cluster's effect size distribution.

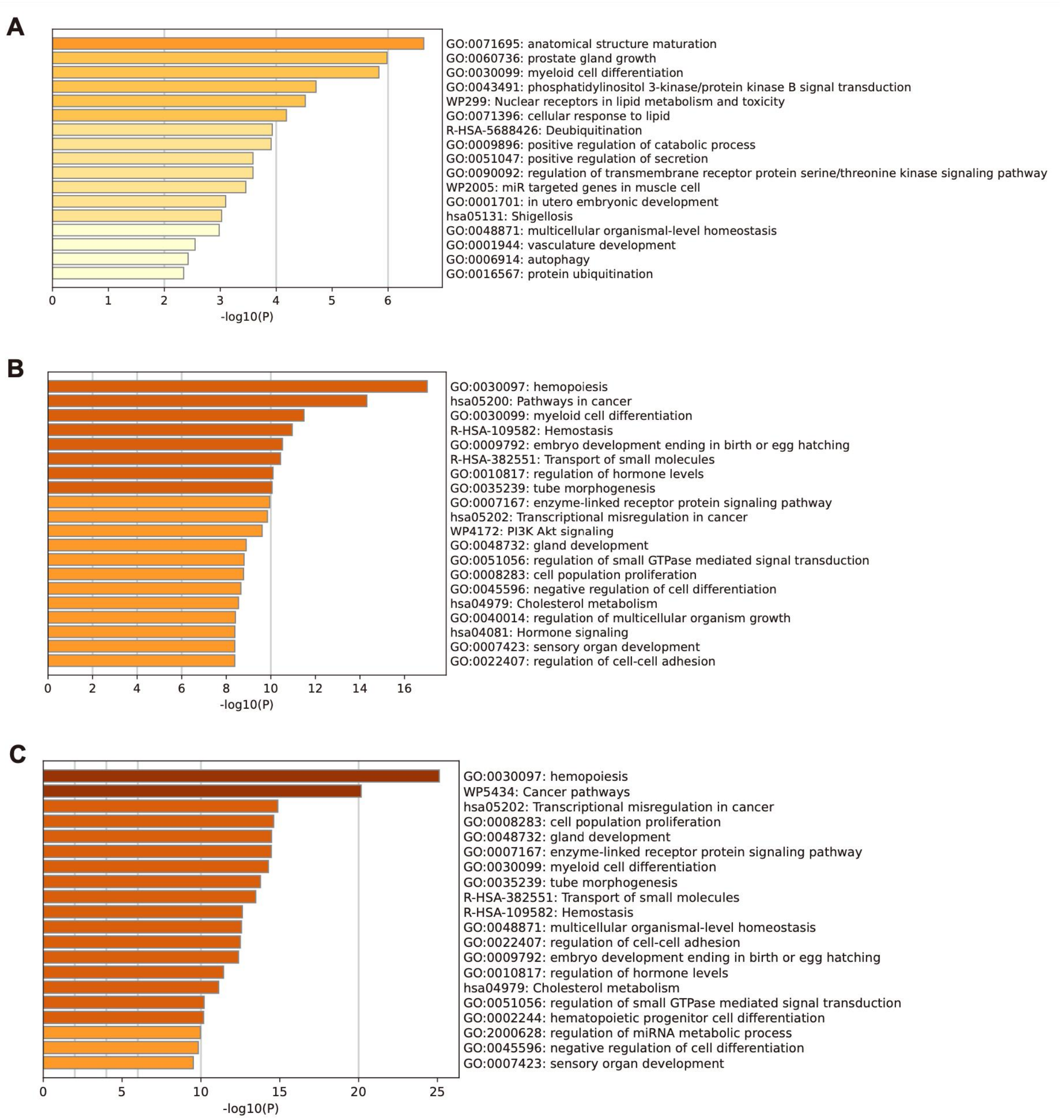

**Supplementary Fig. 13 | Enriched pathways for gestation-specific, general, and all genes.**

Pathway enrichment results are shown for gestation-specific (a), general (b), and all (c) variants. The horizontal axis represents the significance of enrichment.

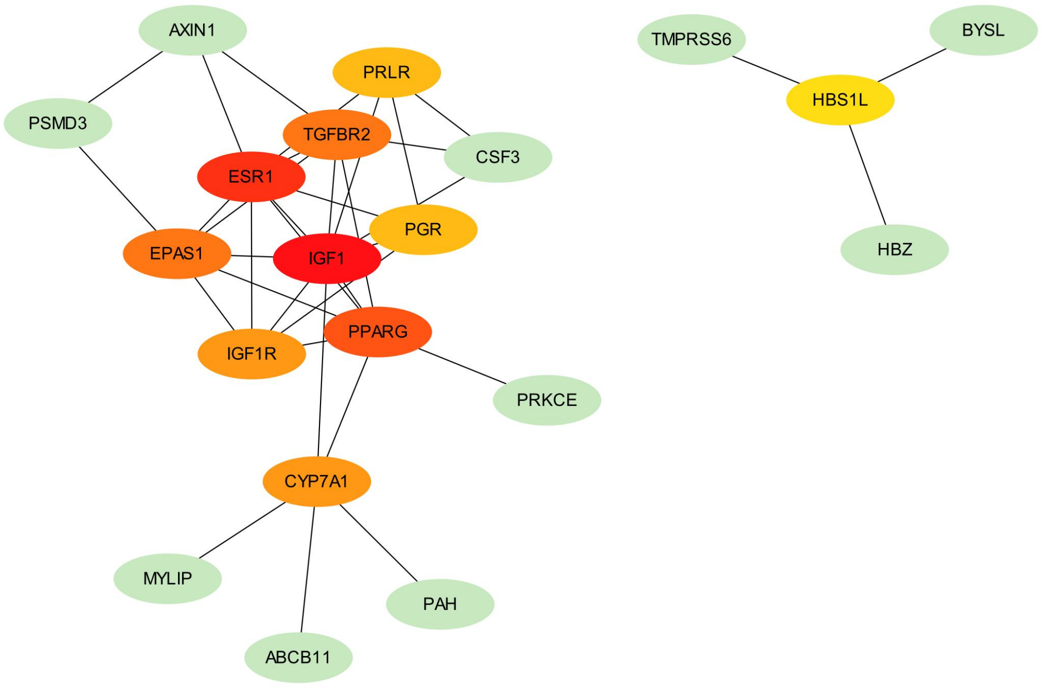

**Supplementary Fig. 14 | Protein–protein interaction (PPI) network of genes with gestation-specific variants.**

Nodes represent genes, with hub genes highlighted in red, orange, and yellow.

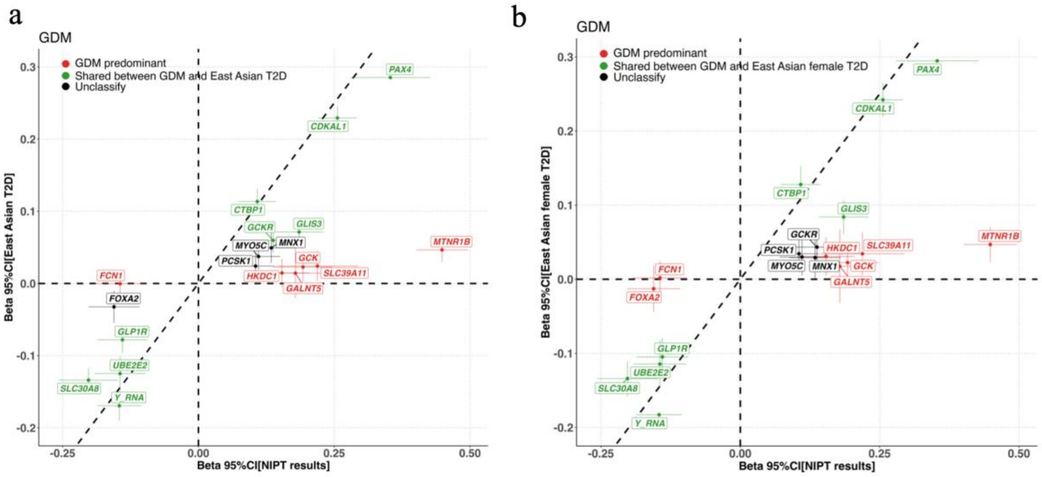

**Supplementary Fig. 15 | Comparison of gestation-specific genetic effects identified using probabilistic modeling and colocalization.**

(a) Comparison of MONN gestational diabetes mellitus (GDM) GWAS with the East Asian type 2 diabetes (T2D) GWAS including sex as a covariate.
(b) Comparison of MONN GDM GWAS with the East Asian T2D GWAS restricted to female participants.

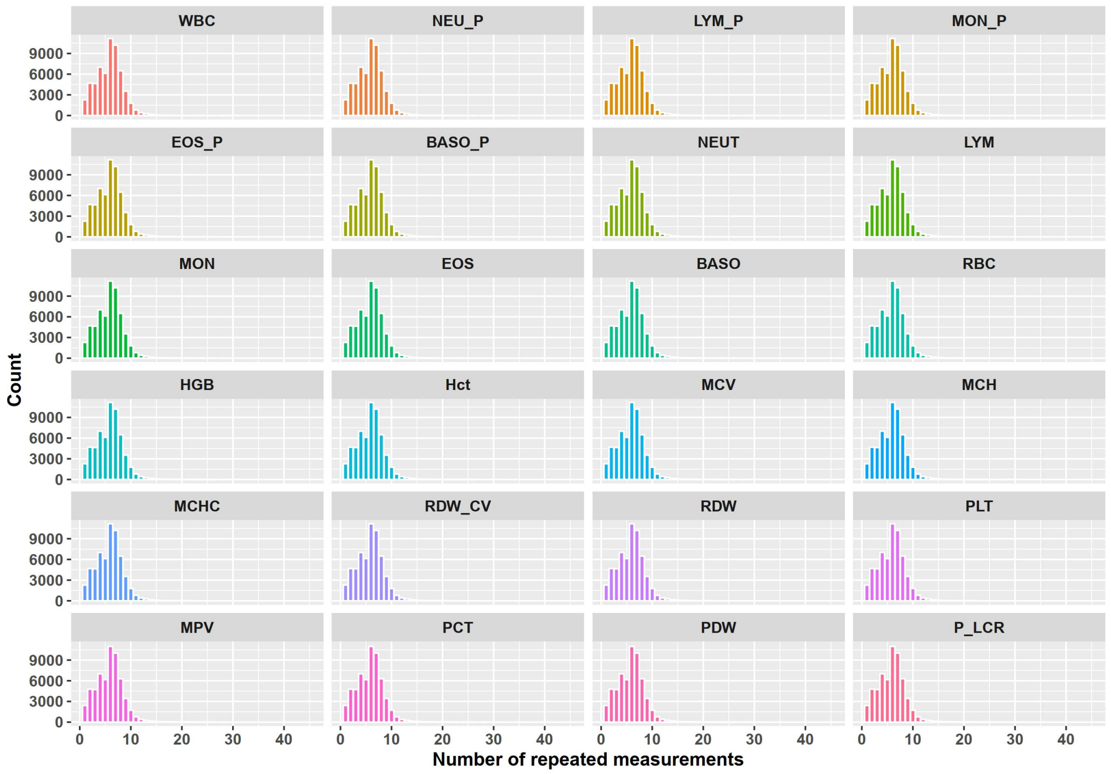

**Supplementary Fig. 16 | Distribution of repeated measurements for 24 complete blood count phenotypes in Longgang Hospital.**

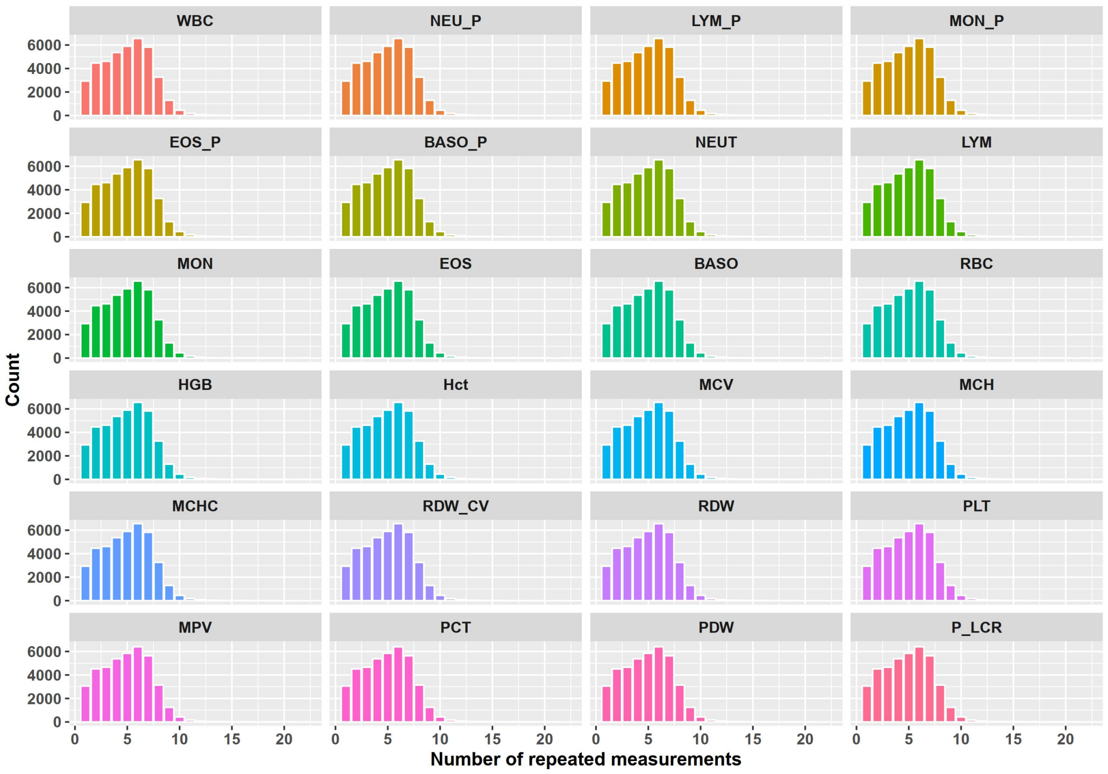

**Supplementary Fig. 17 | Distribution of repeated measurements for 24 complete blood count phenotypes in Baoan Hospital.**

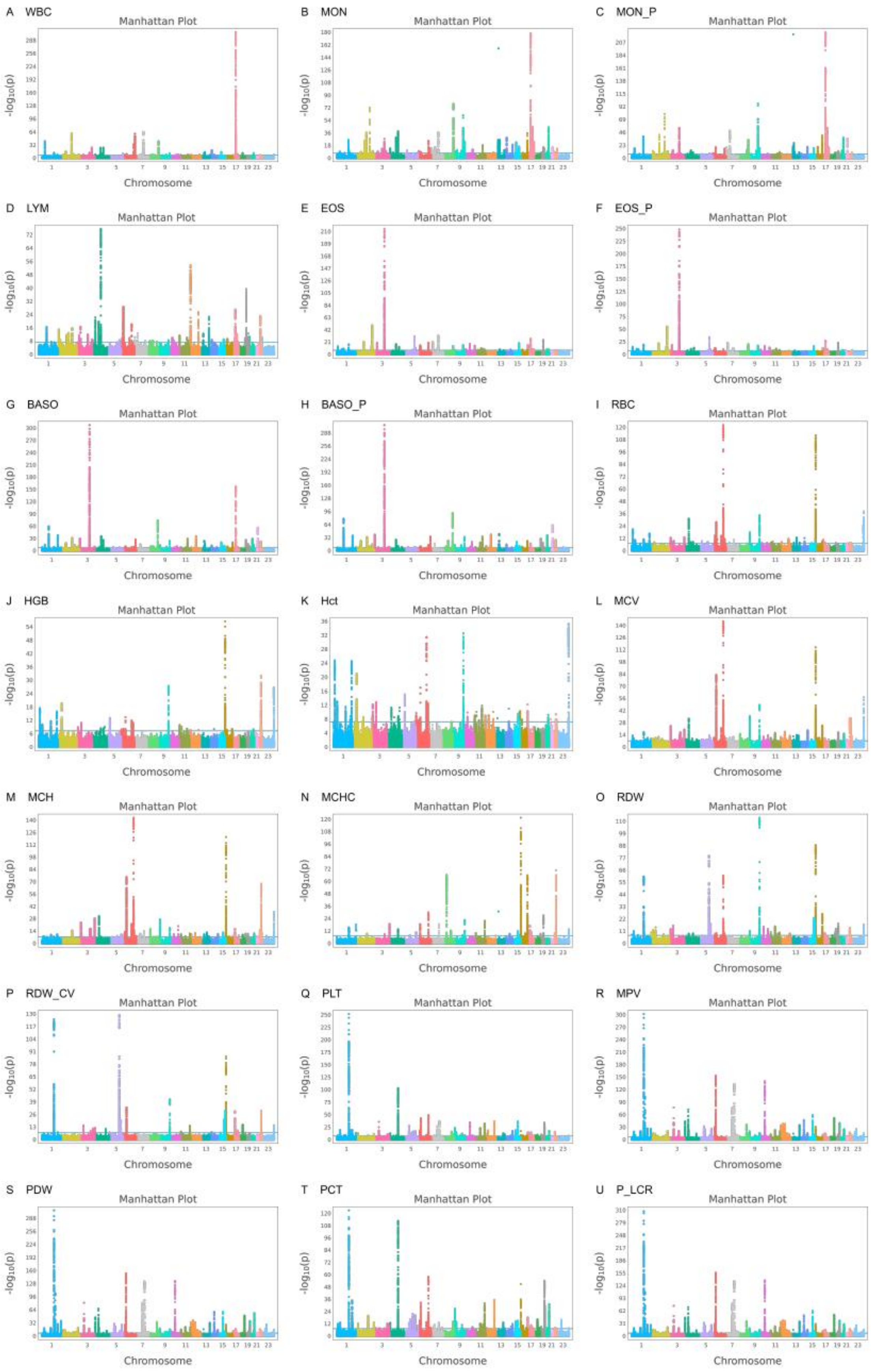
A WBC
B MON
C MON_P
D LYM
E EOS
F EOS_P
G BASO
H BASO_P
I RBC
J HGB
K Hct
L MCV
M MCH
N MCHC
O RDW
P RDW_CV
Q PLT
R MPV
S PDW
T PCT
U P_LCR
Manhattan Plot
Chromosome
-log10(p)

**Supplementary Fig. 18 | Manhattan plots of mean longitudinal complete blood count phenotypes during gestation and postpartum in Longgang Hospital.**

Twenty-four complete blood count phenotypes were analyzed using TrajGWAS across multiple measurements. Plots show the mean values for each longitudinal phenotype, with chromosomes ordered along the x axis and $-\log_{10}(P\text{-value})$ from TrajGWAS on the y axis. The blue horizontal line indicates the genome-wide significance threshold ($P = 5\times10^{-8}$). NEUT, NEU_P, and LYM_P are omitted, as their models did not converge with genetic variants.

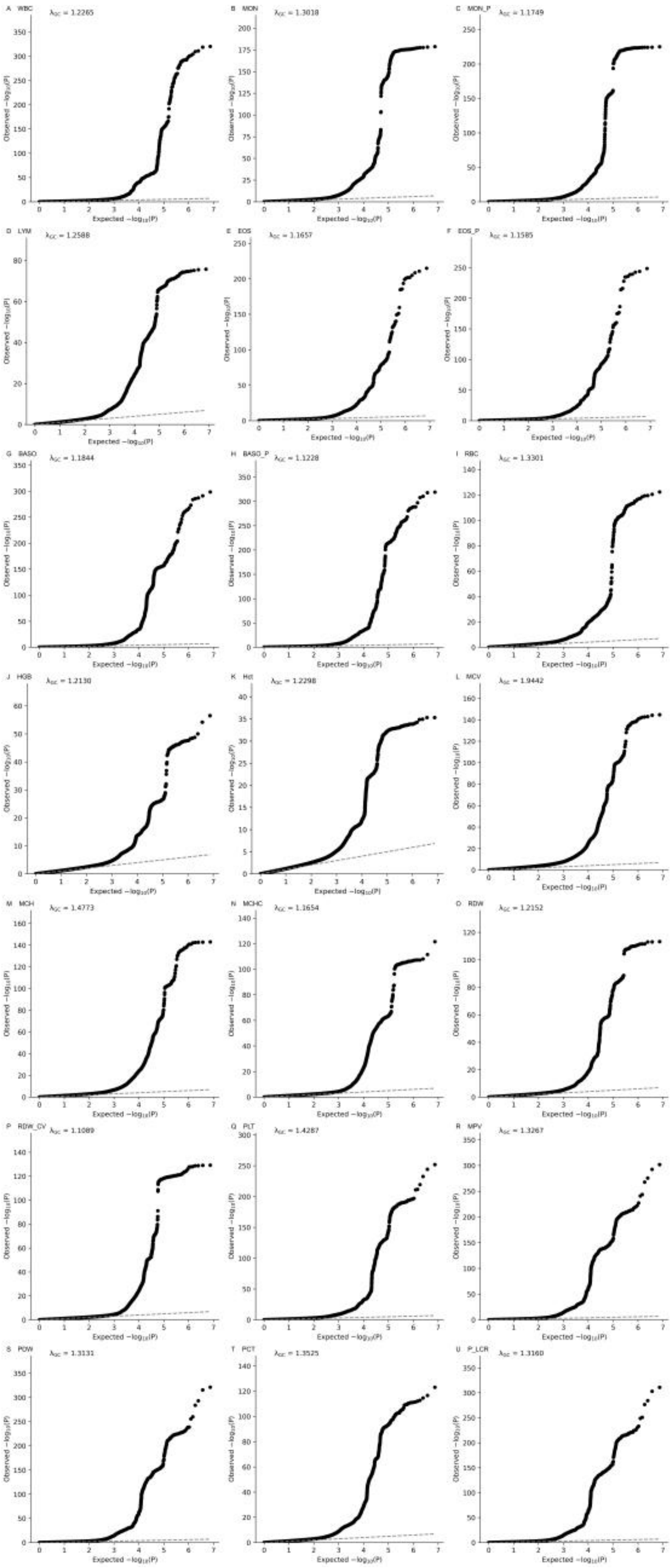
A WBC λGC = 1.2265
B MON λGC = 1.3018
C MON_P λGC = 1.1749
D LYM λGC = 1.2588
E EOS λGC = 1.1657
F EOS_P λGC = 1.1585
G BASO λGC = 1.1844
H BASO_P λGC = 1.1228
I RBC λGC = 1.3301
J HGB λGC = 1.2130
K Hct λGC = 1.2298
L MCV λGC = 1.9442
M MCH λGC = 1.4773
N MCHC λGC = 1.1654
O RDW λGC = 1.2152
P RDW_CV λGC = 1.1089
Q PLT λGC = 1.4287
R MPV λGC = 1.3267
S PDW λGC = 1.3131
T PCT λGC = 1.3525
U P_LCR λGC = 1.3160
Observed −log10(P)
Expected −log10(P)

**Supplementary Fig. 19 | QQ plots for mean longitudinal complete blood count phenotypes during gestation and postpartum in Longgang Hospital (TrajGWAS).**

Observed -$\log_{10}$(*P*-value) from TrajGWAS are plotted against expected -$\log_{10}$(*P*-value) under the null. The red dashed line represents the null distribution, and the gray shaded area indicates standard errors. NEUT, NEU_P, and LYM_P are omitted, as their models did not converge with genetic variants.

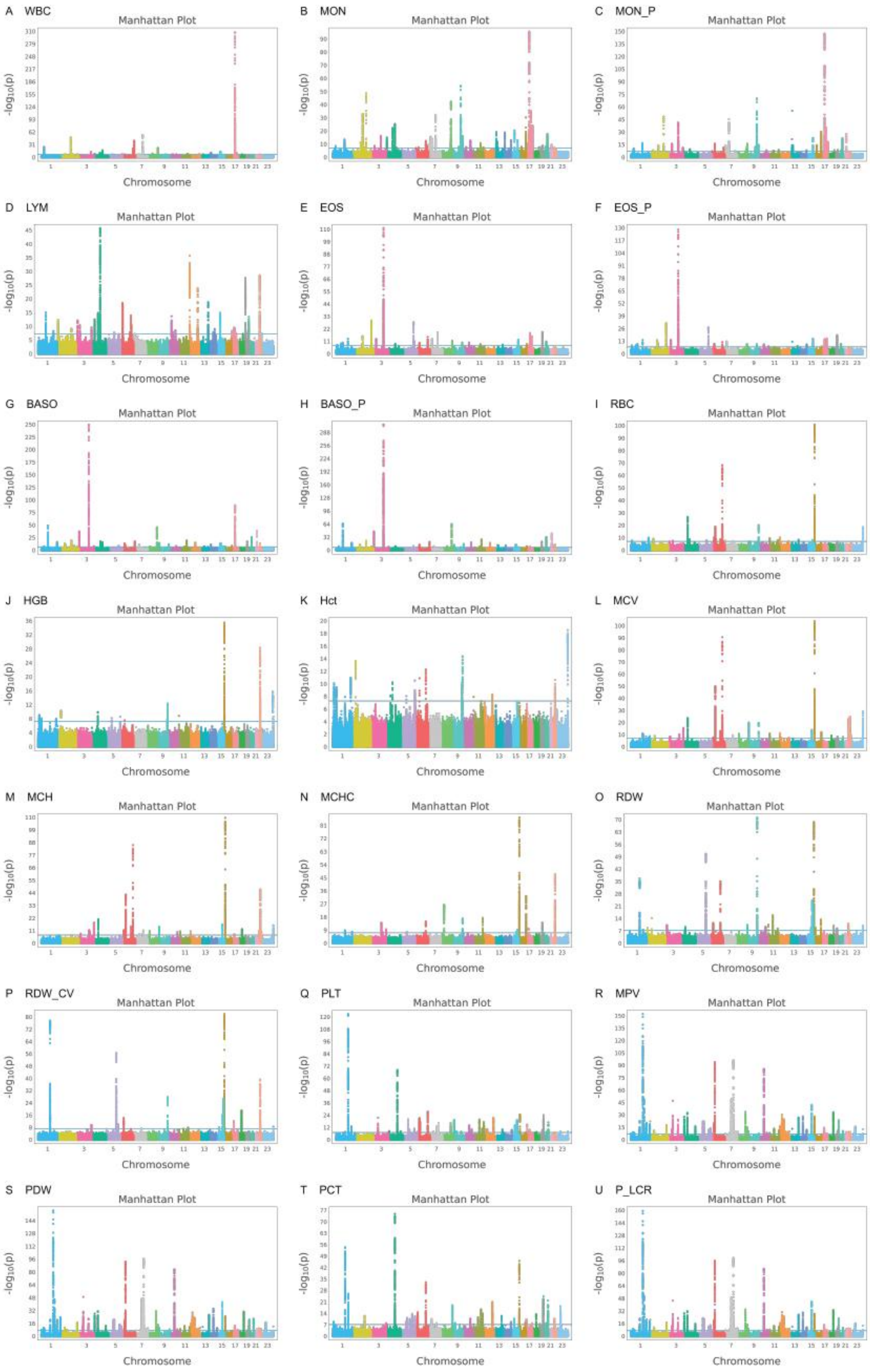
A WBC
B MON
C MON_P
D LYM
E EOS
F EOS_P
G BASO
H BASO_P
I RBC
J HGB
K Hct
L MCV
M MCH
N MCHC
O RDW
P RDW_CV
Q PLT
R MPV
S PDW
T PCT
U P_LCR
Manhattan Plot
-log10(p)
Chromosome

**Supplementary Fig. 20 | Manhattan plots of mean longitudinal complete blood count phenotypes during gestation and postpartum in Baoan Hospital.**

Twenty-four complete blood count phenotypes were analyzed using TrajGWAS across multiple measurements. Plots show the mean values for each longitudinal phenotype, with chromosomes ordered along the x axis and $-\log_{10}$($P$-value) from TrajGWAS on the y axis. The blue horizontal line indicates the genome-wide significance threshold ($P = 5\times10^{-8}$). NEUT, NEU_P, and LYM_P are omitted, as their models did not converge with genetic variants.

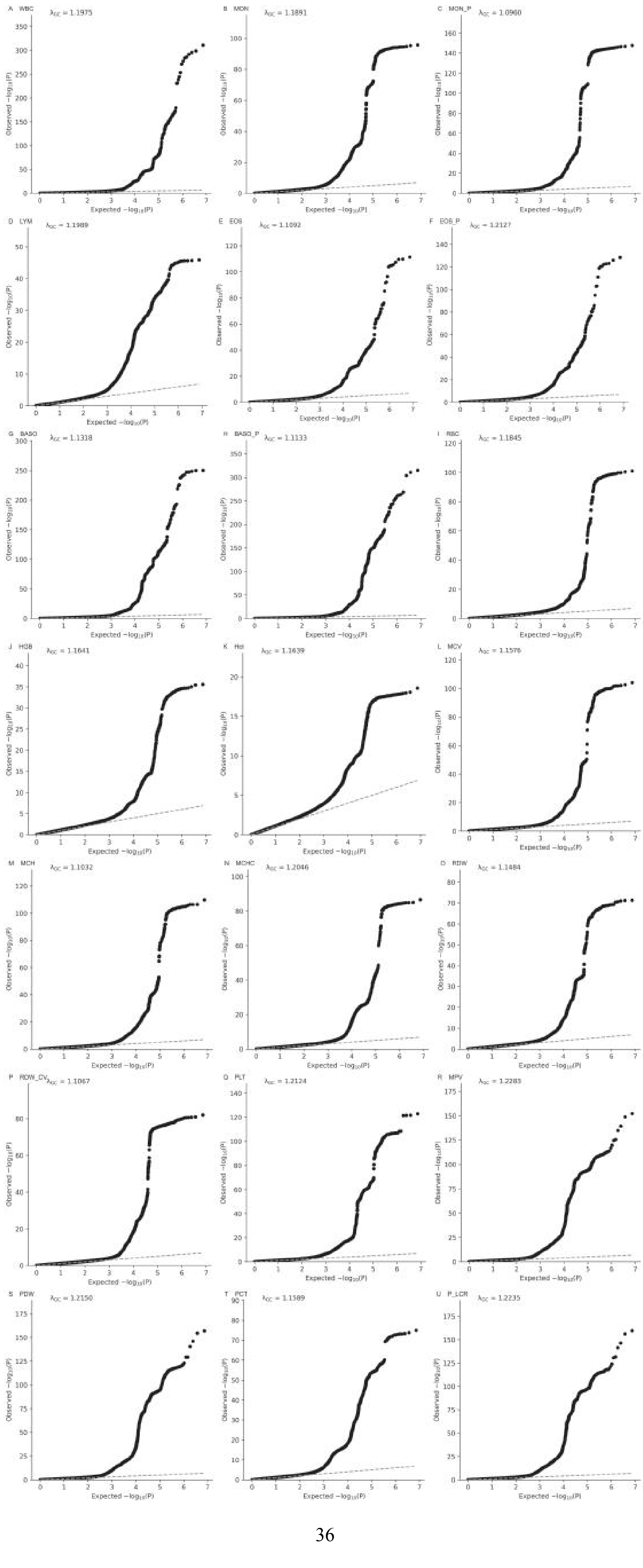
A WBC
λGC = 1.1975
B MON
λGC = 1.1891
C MON_P
λGC = 1.0960
D LYM
λGC = 1.1989
E EOS
λGC = 1.1092
F EOS_P
λGC = 1.2127
G BASO
λGC = 1.1318
H BASO_P
λGC = 1.1133
I RBC
λGC = 1.1845
J HGB
λGC = 1.1641
K Hct
λGC = 1.1639
L MCV
λGC = 1.1576
M MCH
λGC = 1.1032
N MCHC
λGC = 1.2046
O RDW
λGC = 1.1484
P RDW_CV
λGC = 1.1067
Q PLT
λGC = 1.2124
R MPV
λGC = 1.2283
S PDW
λGC = 1.2150
T PCT
λGC = 1.1589
U P_LCR
λGC = 1.2235
Observed −log10(P)
Expected −log10(P)

**Supplementary Fig. 21 | QQ plots for mean longitudinal complete blood count phenotypes during gestation and postpartum in Baoan Hospital (TrajGWAS).**

Observed $-\log_{10}(P\text{-value})$ from TrajGWAS are plotted against expected $-\log_{10}(P\text{-value})$ under the null. The red dashed line represents the null distribution, and the gray shaded area indicates standard errors. NEUT, NEU_P, and LYM_P are omitted, as their models did not converge with genetic variants.

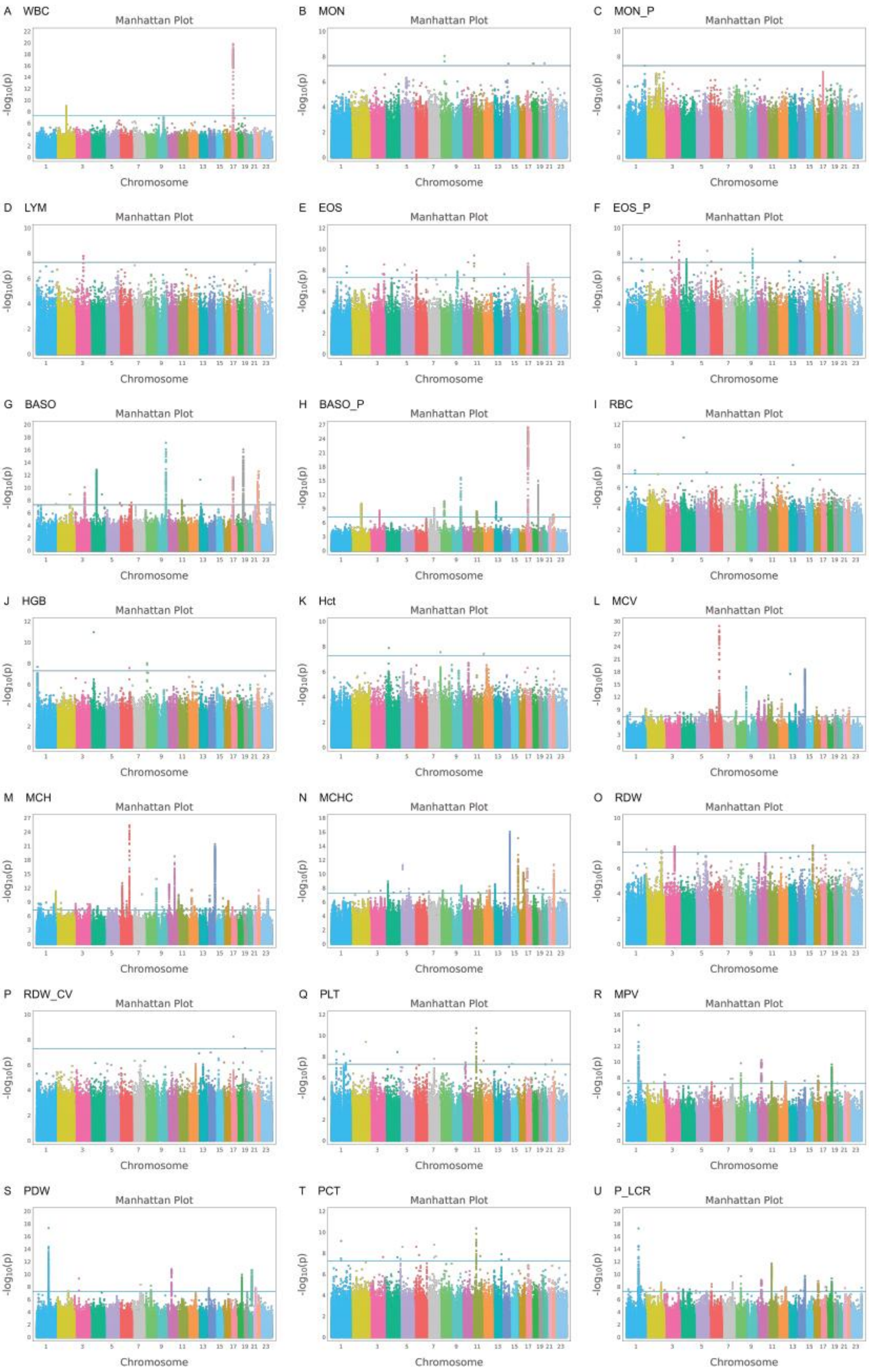
A WBC
B MON
C MON_P
D LYM
E EOS
F EOS_P
G BASO
H BASO_P
I RBC
J HGB
K Hct
L MCV
M MCH
N MCHC
O RDW
P RDW_CV
Q PLT
R MPV
S PDW
T PCT
U P_LCR
Manhattan Plot
-log10(p)
Chromosome

**Supplementary Fig. 22 | Manhattan plots of within-subject variability for longitudinal complete blood count phenotypes during pregnancy and postpartum in Longgang Hospital.**

Twenty-four complete blood count phenotypes were analyzed using TrajGWAS across multiple measurements. Plots show within-subject variability for each longitudinal phenotype, with chromosomes ordered along the x axis and $-\log_{10}$(*P*-value) from TrajGWAS on the y axis. The blue horizontal line indicates the genome-wide significance threshold ($P = 5\times10^{-8}$). NEUT, NEU_P, and LYM_P are omitted, as their models did not converge with genetic variants.

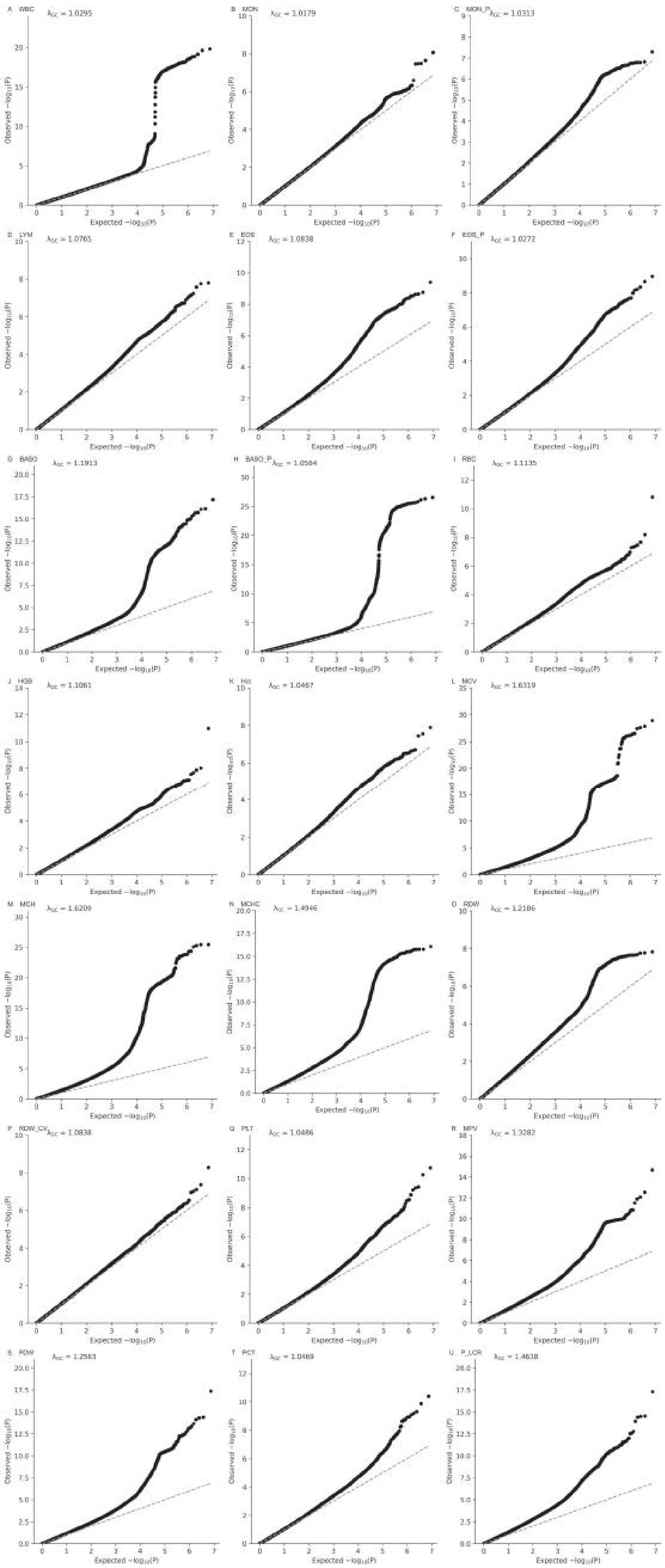
A WBC $\lambda_{GC}$ = 1.0295
B MON $\lambda_{GC}$ = 1.0179
C MON_P $\lambda_{GC}$ = 1.0313
D LYM $\lambda_{GC}$ = 1.0765
E EOS $\lambda_{GC}$ = 1.0838
F EOS_P $\lambda_{GC}$ = 1.0272
G BASO $\lambda_{GC}$ = 1.1913
H BASO_P $\lambda_{GC}$ = 1.0584
I RBC $\lambda_{GC}$ = 1.1135
J HGB $\lambda_{GC}$ = 1.1061
K Hct $\lambda_{GC}$ = 1.0467
L MCV $\lambda_{GC}$ = 1.6319
M MCH $\lambda_{GC}$ = 1.6209
N MCHC $\lambda_{GC}$ = 1.4946
O RDW $\lambda_{GC}$ = 1.2186
P RDW_CV $\lambda_{GC}$ = 1.0838
Q PLT $\lambda_{GC}$ = 1.0486
R MPV $\lambda_{GC}$ = 1.3282
S PDW $\lambda_{GC}$ = 1.2563
T PCT $\lambda_{GC}$ = 1.0469
U P_LCR $\lambda_{GC}$ = 1.4638
Observed $-\log_{10}(P)$
Expected $-\log_{10}(P)$

**Supplementary Fig. 23 | QQ plots for within-subject variability of longitudinal complete blood count phenotypes during gestation and postpartum in Longgang Hospital (TrajGWAS).**

Observed -$\log_{10}$(*P*-value) from TrajGWAS are plotted against expected -$\log_{10}$(*P*-value) under the null. The red dashed line represents the null distribution, and the gray shaded area indicates standard errors. NEUT, NEU_P, and LYM_P are omitted, as their models did not converge with genetic variants.

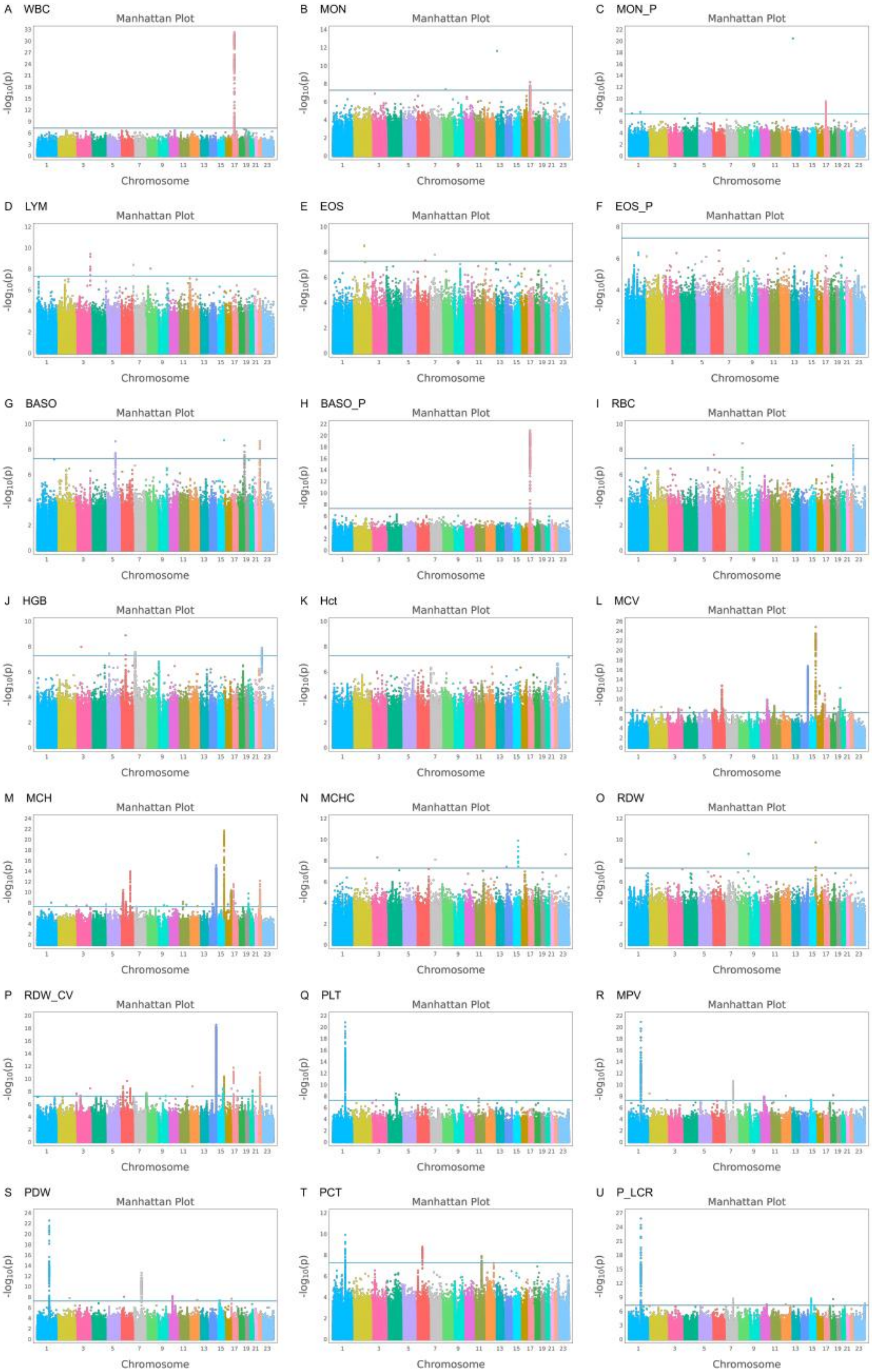
A WBC
B MON
C MON_P
D LYM
E EOS
F EOS_P
G BASO
H BASO_P
I RBC
J HGB
K Hct
L MCV
M MCH
N MCHC
O RDW
P RDW_CV
Q PLT
R MPV
S PDW
T PCT
U P_LCR
Manhattan Plot
Chromosome
-log10(p)

**Supplementary Fig. 24 | Manhattan plots of within-subject variability for longitudinal complete blood count phenotypes during pregnancy and postpartum in Baoan Hospital.**

Twenty-four complete blood count phenotypes were analyzed using TrajGWAS. Plots depict within-subject variability for each phenotype, with chromosomes ordered along the x axis and $-\log_{10}$(*P*-value) from TrajGWAS on the y axis. The blue horizontal line indicates the genome-wide significance threshold ($P = 5\times10^{-8}$). NEUT, NEU_P, and LYM_P are omitted, as their models did not converge with genetic variants.

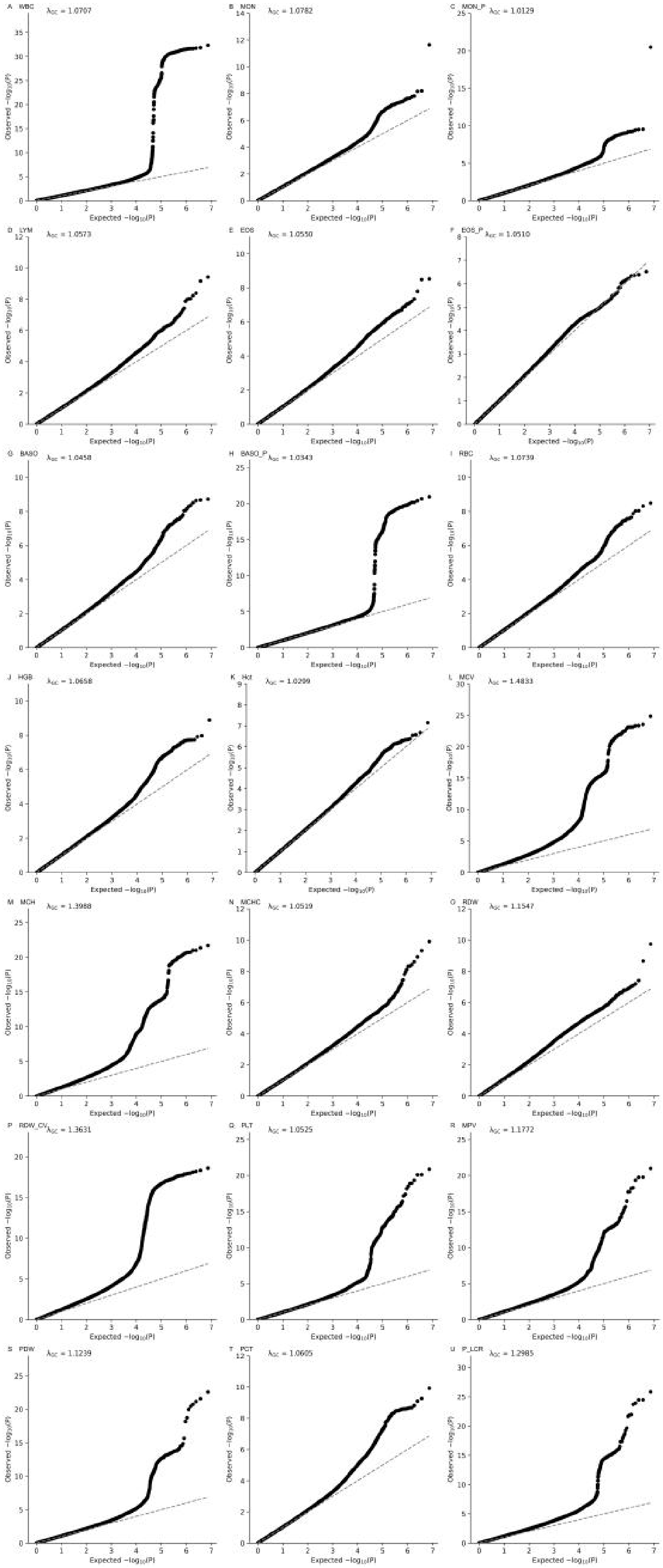
A WBC λGC = 1.0707
B MON λGC = 1.0782
C MON_P λGC = 1.0129
D LYM λGC = 1.0573
E EOS λGC = 1.0550
F EOS_P λGC = 1.0510
G BASO λGC = 1.0458
H BASO_P λGC = 1.0343
I RBC λGC = 1.0739
J HGB λGC = 1.0658
K Hct λGC = 1.0299
L MCV λGC = 1.4833
M MCH λGC = 1.3988
N MCHC λGC = 1.0519
O RDW λGC = 1.1547
P RDW_CV λGC = 1.3631
Q PLT λGC = 1.0525
R MPV λGC = 1.1772
S PDW λGC = 1.1239
T PCT λGC = 1.0605
U P_LCR λGC = 1.2985
Observed −log10(P)
Expected −log10(P)

**Supplementary Fig. 25 | QQ plots for within-subject variability of longitudinal complete blood count phenotypes during gestation and postpartum in Baoan Hospital (TrajGWAS).**

Observed -$\log_{10}$(*P*-value) from TrajGWAS are plotted against expected -$\log_{10}$(*P*-value) under the null. The red dashed line represents the null distribution, and the gray shaded area indicates standard errors. NEUT, NEU_P, and LYM_P are omitted, as their models did not converge with genetic variants.

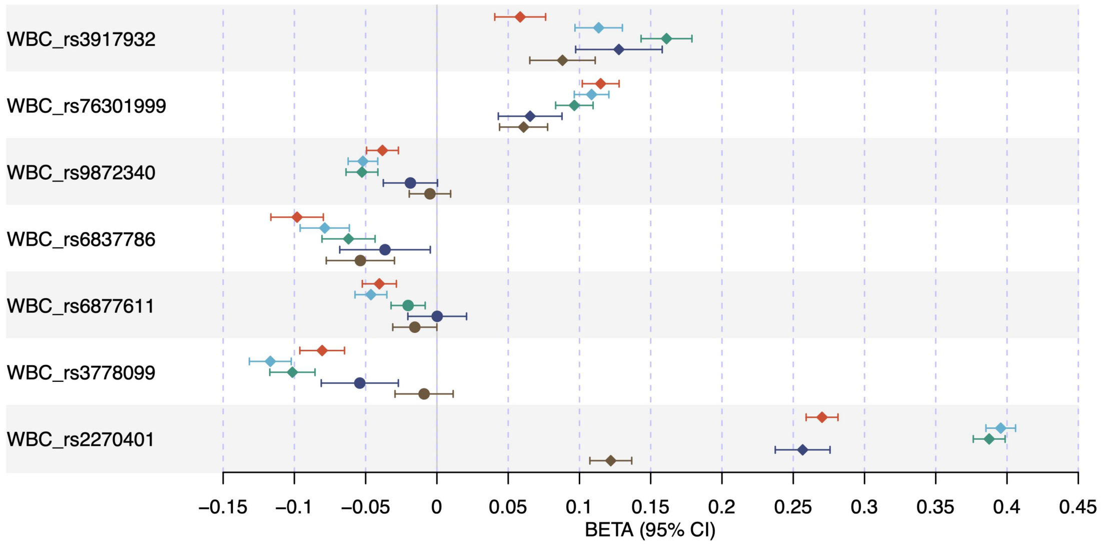

**Supplementary Fig. 26 | Genetic effects of 13 variants across five time periods.**
The forest plot shows results from localized association analyses of 13 variants with time-varying genetic effects. Colors indicate each time period, with corresponding β values and 95% confidence intervals. WBC, whole blood cell counts.

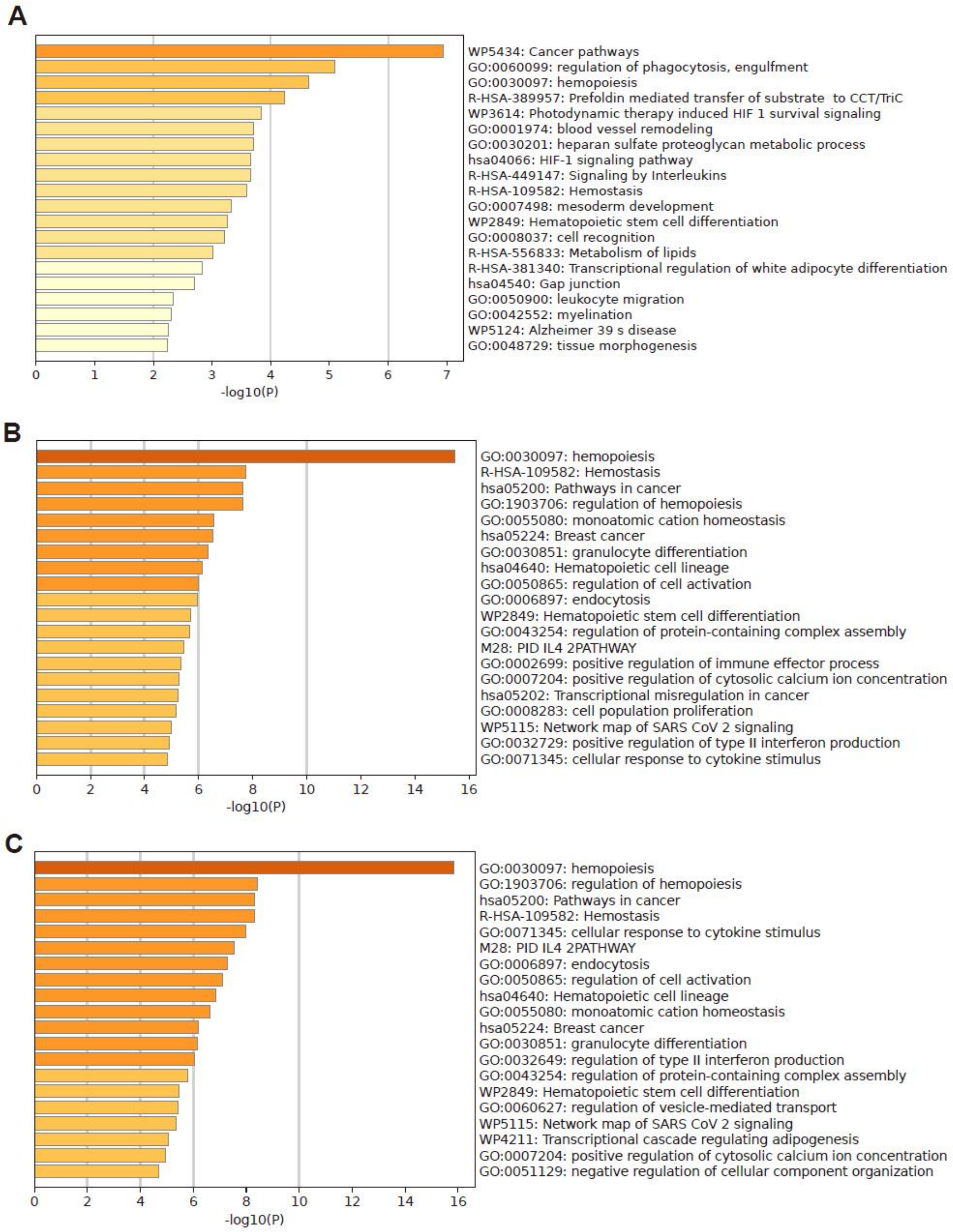

**Supplementary Fig. 27 | Pathways enriched with significant variants identified through longitudinal trajectory analysis.**

(a) Pathways enriched for variants with significant genetic and gene-by-gestational time interaction effects.
(b) Pathways enriched for variants with significant genetic effects but non-significant gene-by-gestational time interactions.

(c) Pathways enriched for all variants with significant genetic effects.

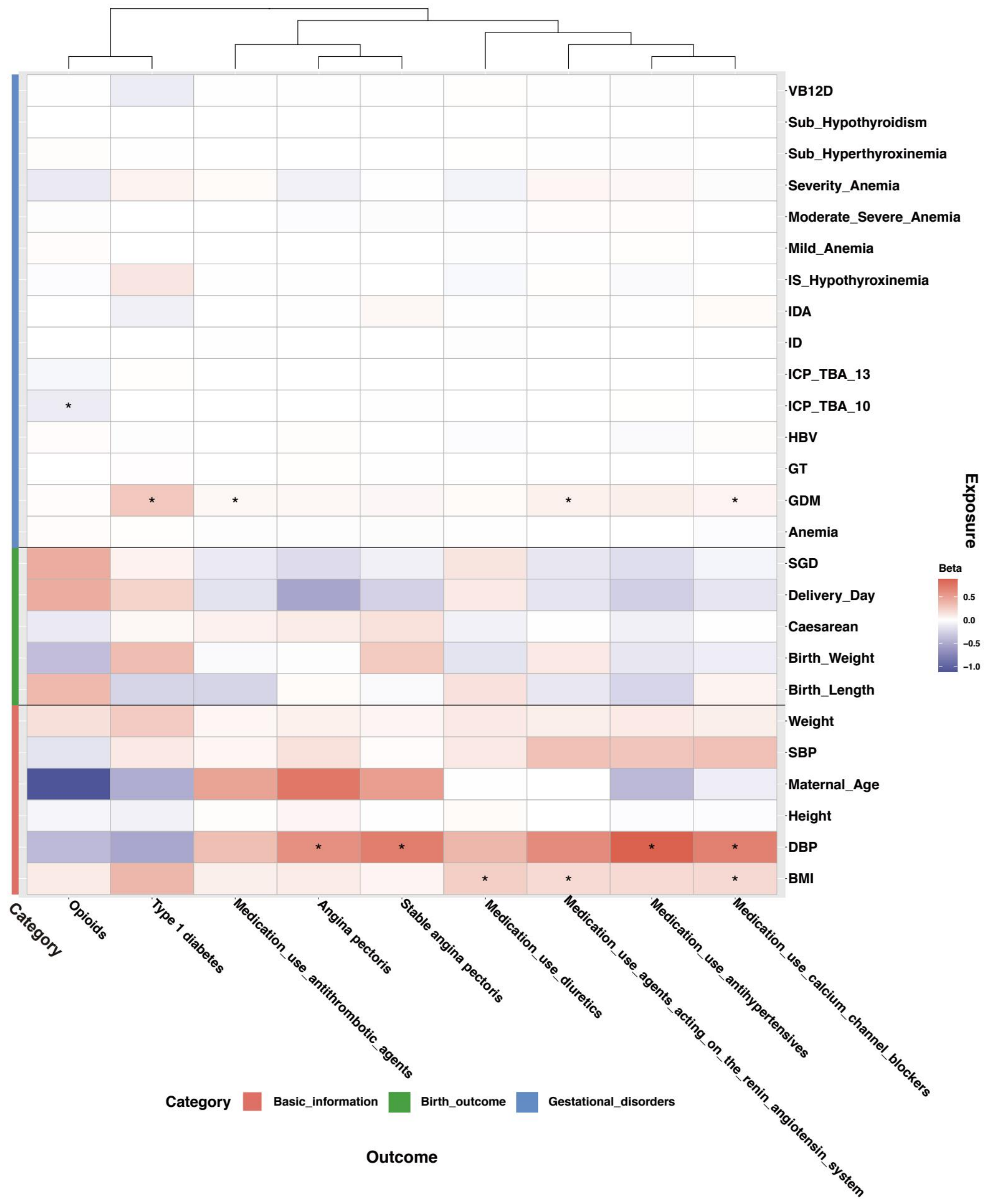

**Supplementary Fig. 28 | Heatmap of Mendelian randomization results for gestational phenotypes and BBJ female disorders.**

Gestational phenotypes (y axis) were used as exposures and BBJ female disorders (x axis) as outcomes. Associations surpassing the FDR threshold are indicated with an asterisk (*, $q < 0.05$). Beta values in the legend and heatmap correspond to inverse-variance weighted (IVW) estimates.

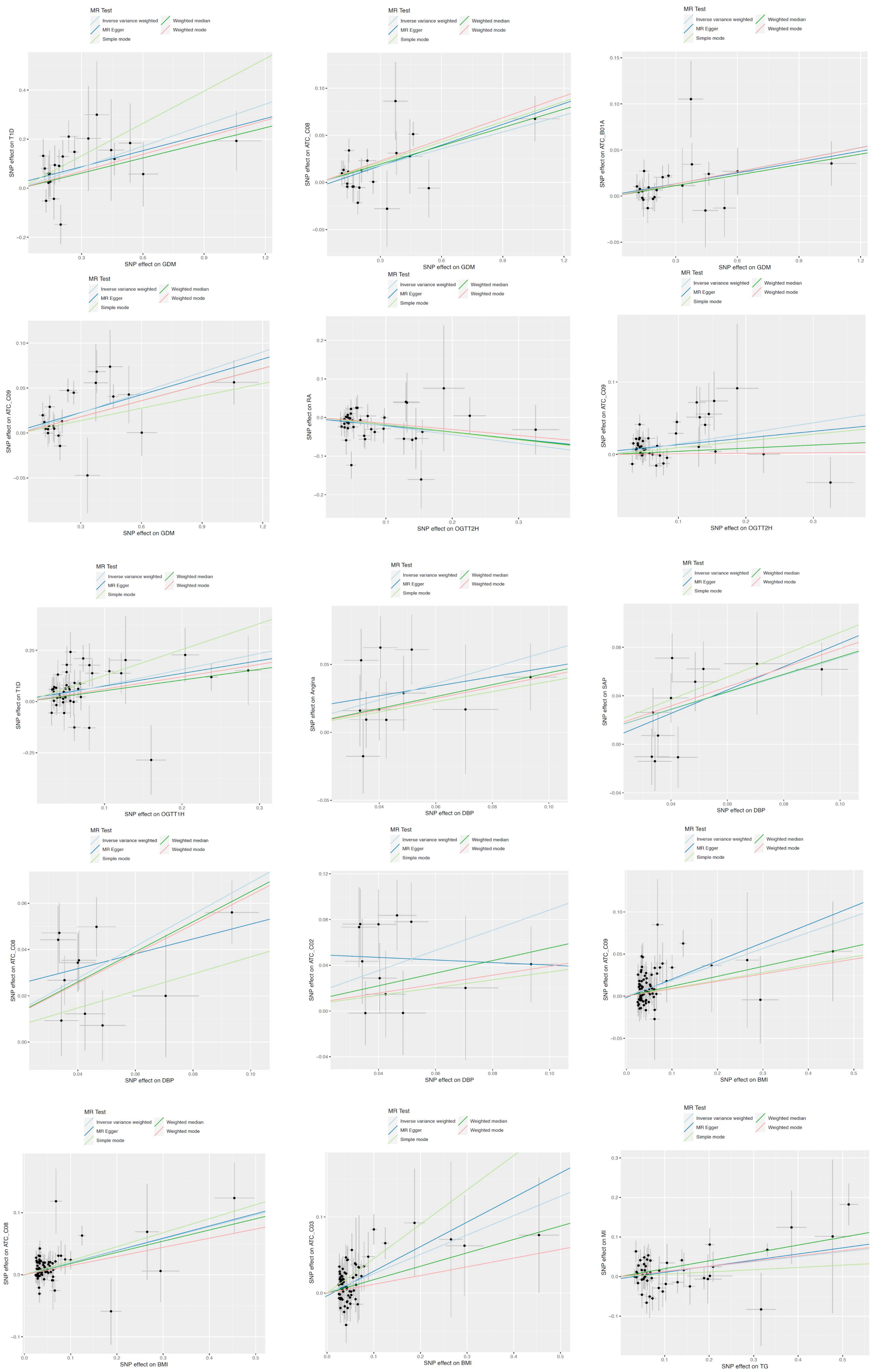

**Supplementary Fig. 29 | Scatter plots of significant Mendelian randomization associations between gestational phenotypes and BBJ female disorders and medication use.**

Significant associations (FDR-adjusted Q < 0.05) are shown for glycemic, blood pressure, body weight, and lipid-related gestational phenotypes.

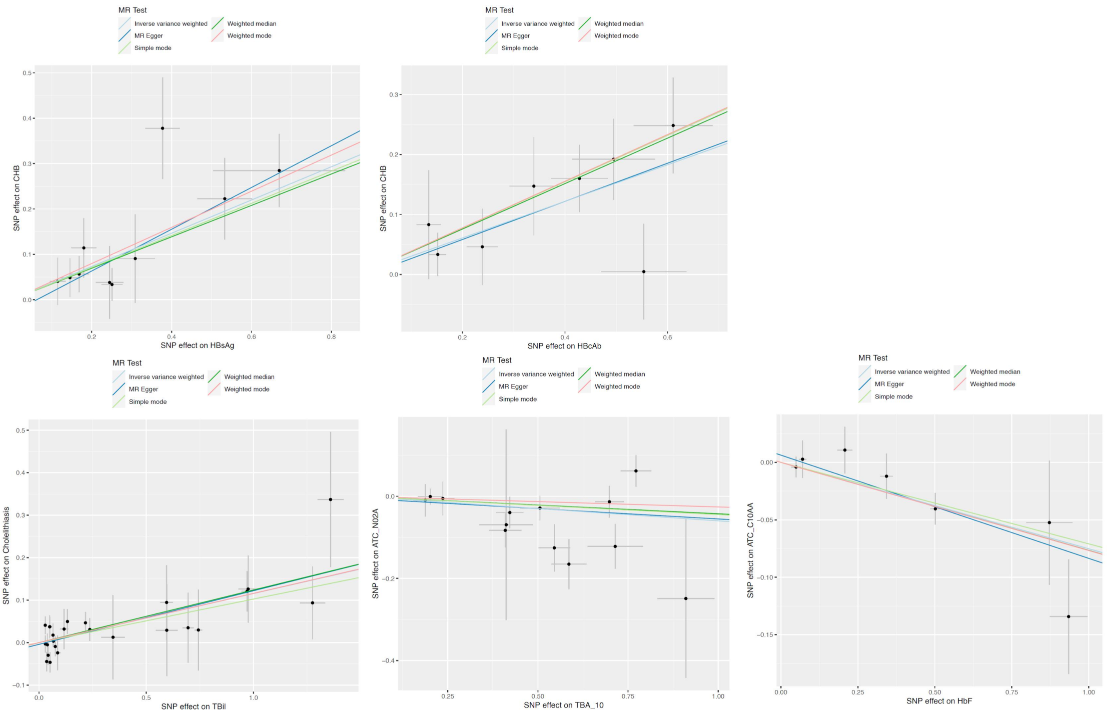

**Supplementary Fig. 30 | Scatter plots of significant Mendelian randomization associations between additional gestational phenotypes and BBJ female disorders and medication use.**

Associations shown are significant at FDR-adjusted Q < 0.05.

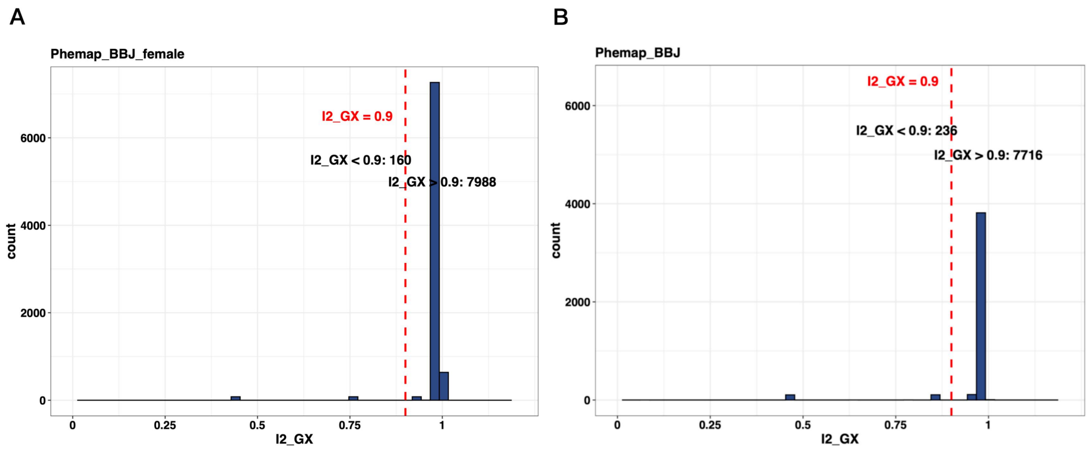

**Supplementary Fig. 31 | Distribution of I²_GX across all exposure–outcome pairs.**

(a) Mendelian randomization analyses between 111 gestational phenotypes and 80 common disorders and medication-use traits in BBJ female participants.
(b) Analyses for the same traits in the full BBJ cohort.
